# Remoscope: a label-free imaging cytometer for malaria diagnostics

**DOI:** 10.1101/2024.11.12.24317184

**Authors:** Paul M. Lebel, Ilakkiyan Jeyakumar, Michelle W.L. Khoo, Chris Charlton, Aditi Saxena, Axel Jacobsen, Emorut James, Emily Huynh, William Wu, Greg Courville, Pei-Chuan Fu, Madhura Raghavan, Robert Puccinelli, Olwoch Peter, Grant Dorsey, Phil Rosenthal, Joseph DeRisi, Rafael Gomez-Sjoberg

**Affiliations:** Chan Zuckerberg Biohub SF, 499 Illinois Street, San Francisco, CA; Infectious Disease Research Collaboration, 2C Nakasero Hill Road, Kampala, Uganda; University of Pennsylvania, Philadelphia, PA; Dept. of Biochemistry and Biophysics, University of California, San Francisco; University of California, Berkeley; Dept. of Medicine, University of California, San Francisco

**Keywords:** Malaria, machine learning, label-free imaging, diagnostics

## Abstract

Malaria diagnostic testing in high transmission settings remains a burden on healthcare systems. Here we present Remoscope, a portable automated imaging cytometer that scans fresh, unstained whole blood using a custom neural network on low-cost hardware. By screening up to two million red blood cells, Remoscope performs label-free quantitative stage-specific detection of *Plasmodium falciparum* (*Pf*) in 1-12 minutes without sample fixation, staining, or slide scanning. Flow is used to achieve high cellular throughput, with blood confined to a 4.5 µm monolayer in low-cost disposable flow cartridges. Remoscope performance was benchmarked *in vitro* by titration of cultured parasites into uninfected whole blood at concentrations of 17.1-710,000 parasites/µL. Counts generated by Remoscope demonstrated a linear response across the entire range. Considering drug susceptibility assays, the half-maximal effective concentration (EC50) of chloroquine (CQ) for the W2 strain of *Pf* was 211 nM by Remoscope, compared to 191 nM for conventional flow cytometry. Remoscope’s real-world diagnostic accuracy was evaluated in a cohort of 500 individuals in eastern Uganda, comprising 601 unique clinic visits. Parallel measurements of parasitemia were performed using Remoscope, qPCR targeting the multicopy conserved var gene acidic terminal sequence, and microscopic evaluation of Giemsa-stained thick blood smears. Remoscope’s limit of detection with respect to qPCR was 95.1 parasites/µL. At this threshold, the system had a sensitivity of 83%, specificity of 96%, Positive Predictive Value (PPV) of 91%, and a Negative Predictive Value (NPV) of 93%. Remoscope’s speed, accuracy, low cost, and ease of use address practical challenges in malaria diagnostic settings around the world. As a general imaging flow cytometer, Remoscope may also inform the development of recognition models for the diagnosis of other infectious and noninfectious blood disorders.

## Introduction

Malaria’s annual death toll of over 600,000 people disproportionately affects low-income regions with limited access to health care. Decades of attempts to control the disease have been modest, and progress has stalled in many regions, especially in sub-Saharan Africa (1). In addition to the emergence and spread of drug-resistant parasites and insecticide-resistant vectors, the recent arrival of the *Anopheles stephensi* mosquito into Africa (2, 3) has begun driving drug and diagnosis-resistant infections upwards in more densely populated urban areas (4), and with it, potential for growing disease and diagnostic burden. Although light microscopy of stained blood smears has remained the gold standard diagnostic for over a century (5–7), the dependence on skilled human labor imposes limitations on accessibility. This leaves under-resourced facilities unable to cope with the daily diagnostic burden, resulting in missed cases, delayed turn-around time, and/or poor diagnostic accuracy (8). Immunochromatographic assays identifying specific antigens (Rapid Diagnostic Tests, or RDTs) are a cost-effective modality but are not quantitative and are susceptible to HRP2 deletions in the *Pf* genome (2, 9). Additionally, HRP2-based RDTs remain positive for variable durations after treatment (10, 11), failing to discriminate live parasites from residual circulating antigen. Although molecular technologies such as PCR, LAMP, and RAMP are highly sensitive (12–17), they depend on cold chains, and their cost remains too high for routine usage in endemic countries. These issues highlight the need for novel approaches to address the ongoing diagnostic burden. Many recent efforts have leveraged machine learning combined with automated slide-scanning microscopes (18–22) to reduce the inspection burden on technicians. While these efforts retain the quantitative advantages of traditional microscopy, many systems require manual smear preparation (18–20), and with it, the corresponding variability in smear quality and dependence on reagents, which contribute to limitations in accuracy and delayed turnaround time. A fully automated commercial slide staining and scanning system was recently developed (21) but is not presently available to malaria-endemic countries, leaving the vast majority of cases unaddressed.

Here we describe Remoscope, a label-free imaging cytometer that was designed to meet the needs of malaria-endemic regions. It performs real-time morphological analysis on up to two million red blood cells in under 12 minutes, for approximately $1 USD per test (Figure 1, Supplementary Figure 1, and Supplementary Video 1). The overall dimensions are approximately 200 x 200 x 300 mm, accommodating labs with limited working space. The cost of goods of the current research prototype is $2200 USD. It uses disposable, ultra-thin flow cartridges (see Methods for fabrication details) to confine fresh, unstained whole blood in a monolayer suitable for single-cell imaging (similar to a thin smear in motion), despite their high concentration at 35-51% hematocrit (23). Operation of the system requires minimal training due to automation, which is achieved by stabilizing the focus, illumination level, and flow rate using feedback (Figure 1b). Images are processed in real time by You Only Glance Once (YOGO, manuscript in preparation), a custom convolutional neural network for cell detection and classification, allowing the system to monitor the density of cells in the chamber and display live parasitemia estimates during the run. Details of the open-source instrument control software are provided in Supplementary Figure 2, described in the Methods, and published on Github.

**Fig. 1.**
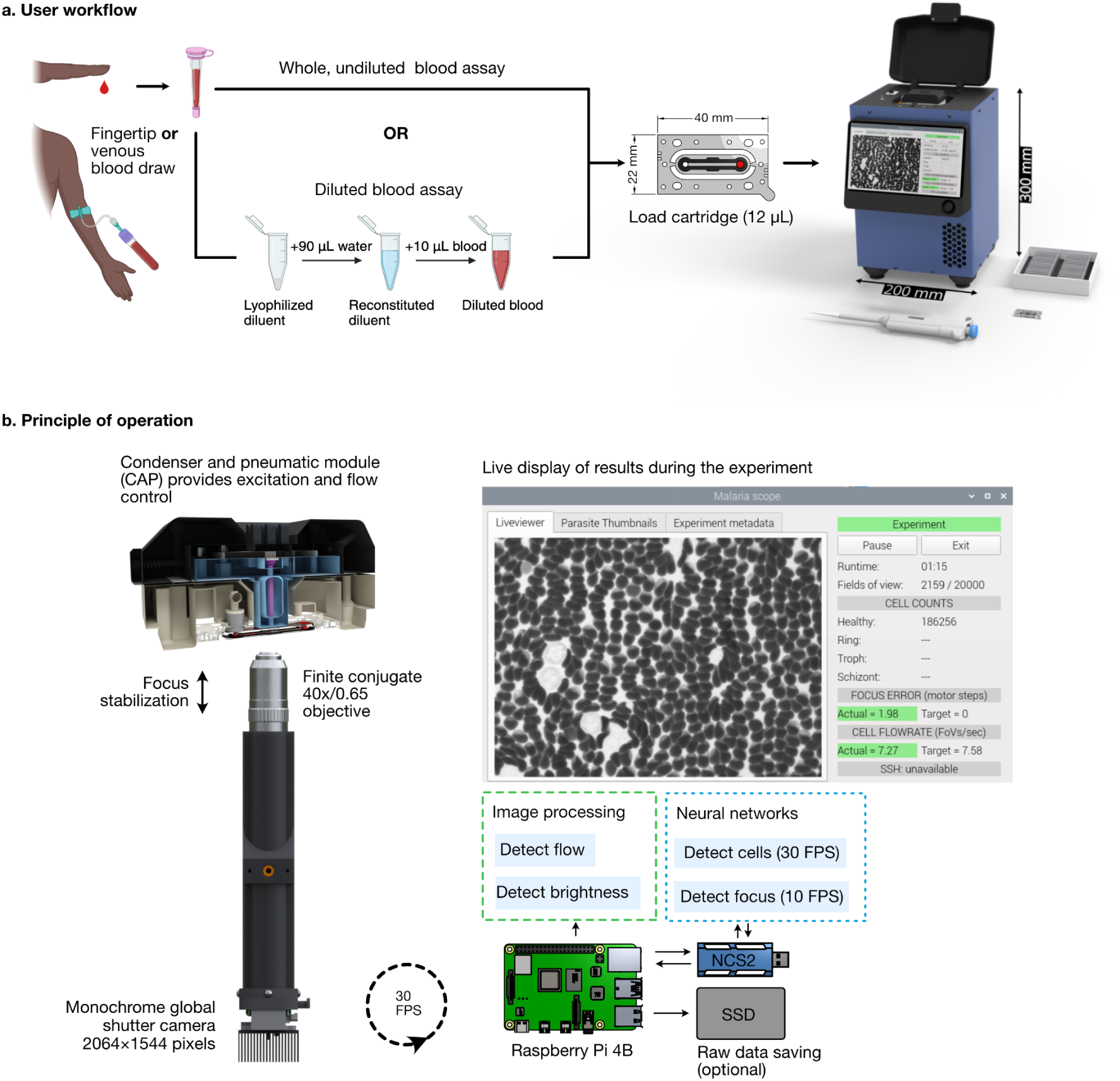
Overview of Remoscope’s workflow and principle of operation. **a**, The user workflow consists of a blood draw (either fingertip or venous, with EDTA anticoagulation), followed by an optional dilution step, chamber loading, metadata entry, run start, and monitoring for completion (1-12 minutes). **b**, High-level overview of the system processes. During continuous image acquisition, the rate of blood flow and image brightness are monitored by classical image processing on the Raspberry Pi, while focal position and cell detection are both performed using neural networks executed on the Neural Compute Stick 2 (NCS2).

In contrast with other methods, it does not require sample pre-processing aside from anticoagulation with Ethylenediaminetetraacetic acid (EDTA), enabling the screening process to begin soon after the blood draw. Additionally, real time analysis permits early termination of a run once the counting statistics meet a given threshold, allowing the system to perform many tests in rapid succession for highly parasitized blood (see Supplementary Figures 3 and 4 for runtime estimates under different conditions). However, because the clinically relevant range of parasitemia spans many orders of magnitude (13) and directly affects patient outcomes, it is helpful for a malaria test to be quantitative and to express bounds of uncertainty, which is dependent on the parasitemia and the number of cells scanned. On board statistical analysis accounts for both stochastic sampling error as well as removal of systematic bias in cell classification, enabling results that are operator independent.

In this work, we quantify Remoscope’s performance on *in vitro* cultured *Pf* as well as in a clinical setting on a cohort of individuals in Eastern Uganda. We also demonstrate its ability to distinguish drugged from un-drugged *Pf* parasites using an *in vitro* half-maximal effective concentration (EC50) assay with chloroquine (CQ).

## Results

### Working principle and methodology

Remoscope uses a simple brightfield imaging modality at 405 nm (Supplementary Figure 5) to image cells under flow, foregoing the cost and complexity of high speed XY motion control hardware required for slide scanning, as well as the sophisticated laser optics and processing electronics used in conventional flow cytometers. Since imaging throughput is achieved via flow, the only motion control systems in Remoscope are for the focus adjustment (Supplementary Figure 6) and for blood flow control, both of which use inexpensive actuators. The disposable cartridges have a 4.5 µm flow layer, which is thin enough to restrict RBCs from tumbling end-over-end, but thick enough to allow passage of the majority of white blood cells (Supplementary Figure 1 and Methods). The system controls the flow rate by exerting vacuum on the distal port with a pneumatic system (Supplementary Figure 7). The moving cells are imaged by a microscope objective with 40× magnification / 0.65 numerical aperture, and immediately processed by YOGO on low-cost computing hardware. YOGO was derived from YOLO (24) and its successors (25–27) but enhanced for inference speed by customization for detected objects consisting of few classes, which span a modest range of sizes. It was trained to recognize seven object classes: healthy RBCs, four different parasitized RBC stages (ring, trophozoite, schizont, and gametocyte), as well as White Blood Cells (WBCs) and miscellaneous objects (platelets and debris). YOGO can run at 30 frames per second on low-cost hardware in the field, enabling a morphological screening rate of about 2,775 cells per second. Intermediate results are updated and displayed to the user throughout the course of the experiment, while focus, brightness, and flow rate are stabilized using feedback. During the run, the statistics of the parasite counts are updated once per second, triggering the end of the experiment if the stop conditions are met. Raw data can optionally be saved to a Solid State Drive (SSD) during the experiment.

Remoscope uses label-free imaging at 405 nm in order to distinguish parasites without the use of fixation or staining (22). Coinciding with the optical absorption maximum of hemoglobin (Hb), this wavelength is also transmitted efficiently by conventional microscope optics. This illumination provides contrast primarily due to the redistribution of Hb by the parasite throughout development (22) (Figure 2). Early and canonical ring forms are visible as light shapes on a dark cytoplasmic background due to cytoplasmic Hb exclusion, while still lacking solid hemozoin (Hz) crystals. More developed trophozoite stages become visible with increased contrast and size, as well as formation of Hz puncta. Schizonts are delineated by the formation of nascent merozoites and a large, centralized Hz crystal (Figure 2). Gametocytes, when present, were distinguishable beyond stage III by their distention of the RBC membrane and elongated morphology. A detailed image annotation strategy is described in Supplementary Note 1: Remoscope annotation guide.

**Fig. 2.**
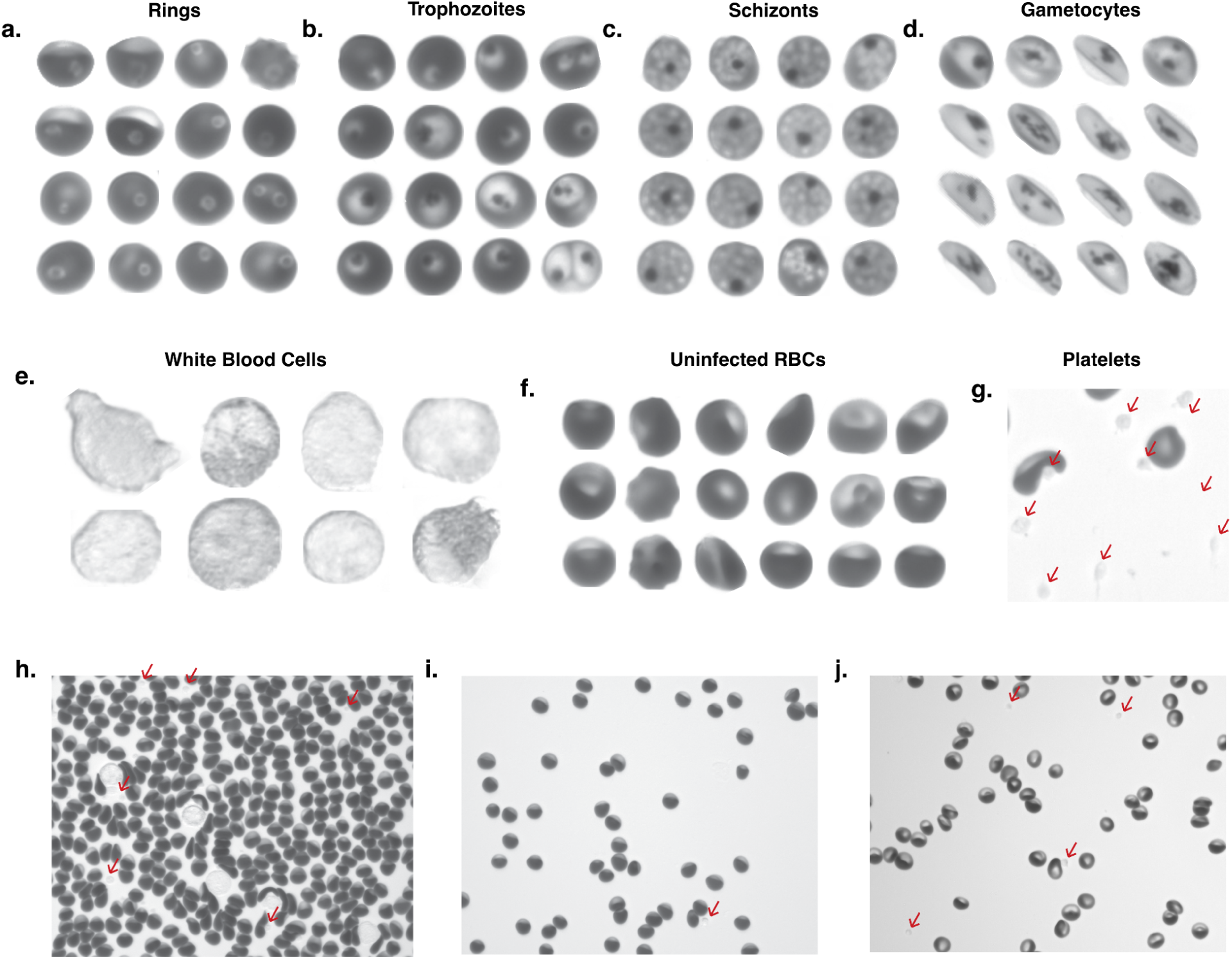
Examples of label-free blood cell images classified by Remoscope. **a**, Ring-infected RBCs of various sub-morphologies including amoeboid, dendritic, and canonical rings. **b,** Trophozoite-infected RBCs of various sub-morphologies including early trophozoites, late trophozoites, and multiply-infected RBCs (upper and lower right). **c,** Schizont-infected RBCs showing partial and full merozoite segmentation. **d,** Gametocytes, ranging from stage 3-5. Stage 1-2 gametocytes were not distinguished from trophozoites due to lack of sufficiently unique features. **e,** White blood cells **f,** Uninfected RBCs exhibiting a variety of morphologies. **g,** A region of interest with a high density of platelets, which are called out with red arrows in panels g-j. **h,** A complete field of view containing uninfected, undiluted blood under flow. RBCs are seen with shear distortion features such as rarified trailing edges. **i,** A complete field of view containing uninfected RBCs diluted to 5% hct, with normal hemoglobin content. **j,** A complete field of view containing diluted blood at 5% hematocrit, exhibiting low hemoglobin pigmentation.

Both healthy whole blood as well as lab-cultured W2 and NF54 *Pf* strains were used for development and testing. The ability to process undiluted blood in Remoscope flow cartridges was realized partway through this work, and was used for all *in vitro* experiments described here, while the clinical data was acquired with 10× diluted blood. Model training followed by intrinsic and extrinsic validation strategies were supported by experiments using various ratios of cultured *Pf* and healthy blood. Unsynchronized parasite cultures provided the bulk of initial training data, where high parasitemia samples could provide a diverse range of life cycle stages containing a high fraction of infected cells. Our annotation strategy consisted of three phases. First, a generic CellPose (28) model was used for preliminary cell-finding to locate the large majority of cells in the sample. Bounding box information from CellPose was then imported into Label Studio (community edition, labelstud.io) (29), where we applied infection-specific labels for YOGO model training and to correct for missing or incorrect bounding boxes. These annotations were used for initial YOGO training. Second, with increasing data volume and model performance, YOGO itself could be used for pre-labeling with parasite identification, accelerating the Label Studio process. Finally, image thumbnails of specific under-annotated cell class instances were manually checked and their labels corrected, and then used for re-training. This thumbnail annotation process was used for all subsequent model iterations, including the incorporation of clinical parasitemia from a Ugandan cohort. Table 1 shows a summary of fully-annotated cells by class and source. See Supplementary Figure 8 for a summary of basic shape parameters of red blood cell classes.

**Table 1.**
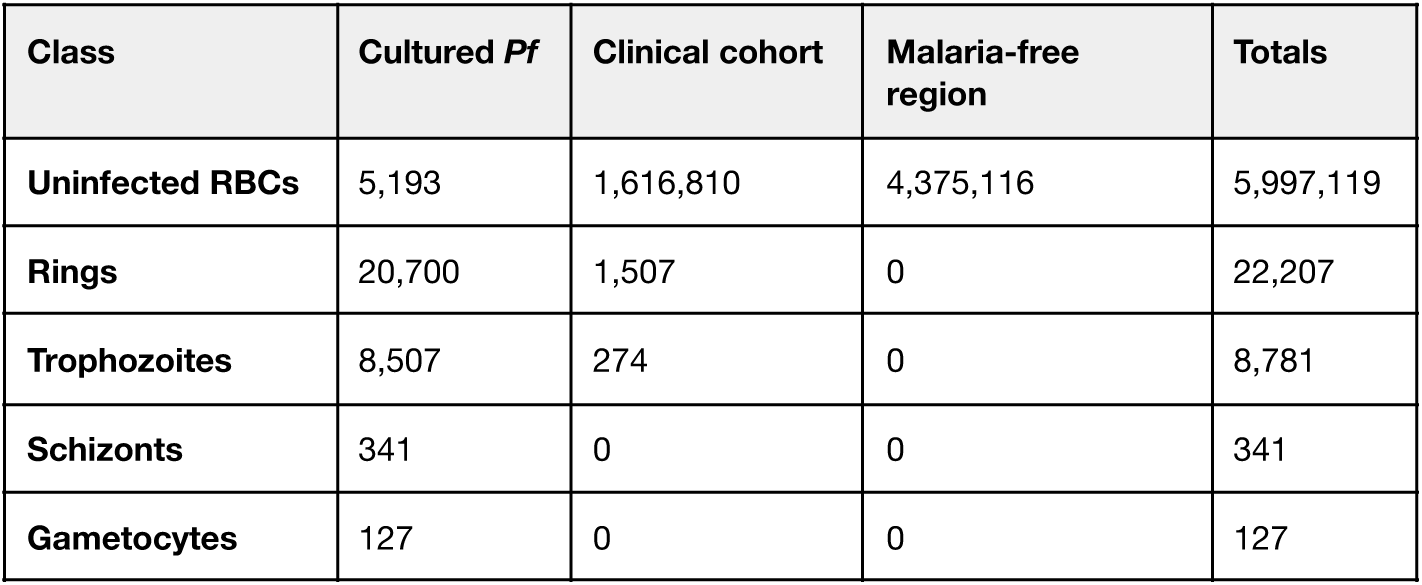
Summary of fully-annotated cell classes used for model fine-tuning. The total number of annotated cells of each category is listed for each of three separate sources: a lab-cultured W2 strain of *Pf*, a Ugandan clinical cohort, and blood donated from a malaria-free region. We define fully-annotated as either: a) every cell has been annotated by a human, b) Malaria-free donation source (all RBCs assigned *en masse* as healthy/uninfected), or c) a holdout “train” partition of images from the Ugandan cohort that were all double-negative by both qPCR and microscopy, assigned *en masse* as healthy/uninfected. All parasite class counts listed above were human-annotated either in label-studio or by the thumbnail annotation described above and in the Methods section.

| Class | Cultured *Pf* | Clinical cohort | Malaria-free region | Totals |
| --- | --- | --- | --- | --- |
| Uninfected RBCs | 5,193 | 1,616,810 | 4,375,116 | 5,997,119 |
| Rings | 20,700 | 1,507 | 0 | 22,207 |
| Trophozoites | 8,507 | 274 | 0 | 8,781 |
| Schizonts | 341 | 0 | 0 | 341 |
| Gametocytes | 127 | 0 | 0 | 127 |

### Calculation of parasitemia, dataset vetting, and uncertainty estimation

System performance was evaluated using both lab-cultured parasites as well as in a clinical Ugandan cohort with respect to qPCR and traditional Giemsa microscopy of thick blood smears. Parasitemia assessments depend on the accurate estimation of overall sample composition, which is a function of per-cell model performance (see Supplementary Figures 9 and 10 for model metrics), the total number of cells imaged, as well as other parameters such as confidence threshold and data compensation strategies. Due to the large class imbalance inherent to the problem – clinical parasitemia levels span more than five orders of magnitude (13) – the false positive rate of healthy cells scored as infected presented a challenge: even 1/100,000 healthy cells wrongly classified as infected will impact diagnostic performance. In particular, uninfected RBCs exhibit a large diversity of shapes caused by flow shear distortion, collisions with other cells or surface-bound debris, and many other physiological and biochemical factors influencing RBC shape and appearance (30, 31). With increasing flow velocity, RBC morphologies were observed to shift from canonical discocytes to morphologies characterized by a dense leading edge and a rarified trailing edge (Supplementary Figure 11), whose geometries must all be classified as healthy. To mitigate deformed but uninfected cells from causing false positives, our strategy relied on annotating cells as infected only in cases of high annotator confidence and unanimous agreement on infected morphologies (see Fig. 2 and Supplementary Note 1). Although this strategy likely resulted in a higher False Negative Rate (FNR) on a per-cell basis, mitigation of the False Positive Rate (FPR) was prioritized due to the class imbalance’s effect on sample parasitemia estimation.

Stage-specific and overall parasitemias were computed by summation of all the detected parasite classes in a given dataset, normalized by the total number of red blood cells. Remoscope’s raw recall with respect to other testing modalities was assessed systematically for both *in vitro* and *in vivo* conditions, and found to be on average lower than both Giemsa and qPCR. Raw parasitemia values could therefore be linearly transformed to compensate for known average recall and false positive rates, using parameters derived from least squares regression to variational data. We used the best fit parameters to transform Remoscope results such that they were quantitatively equivalent to qPCR results. For uncertainty estimations, we considered both stochastic and correlated sources of error. Linear transformations as well as 95% confidence interval calculations are derived in Supplementary Note 2, relative to a YOGO confusion matrix (model recall, stage-specific), or relative to Giemsa and/or qPCR (overall recall). While sampling error in the form of fundamental Poisson counting statistics determined the baseline uncertainty in all cases, imperfect knowledge of transformation parameters added some additional uncertainty to transformed results. In addition to theoretical estimates, we also provide sampling error analysis with respect to a cultured parasite titration (Figure 3), and clinical diagnostic performance metrics with respect to qPCR (Figure 4f-n).

**Fig. 3.**
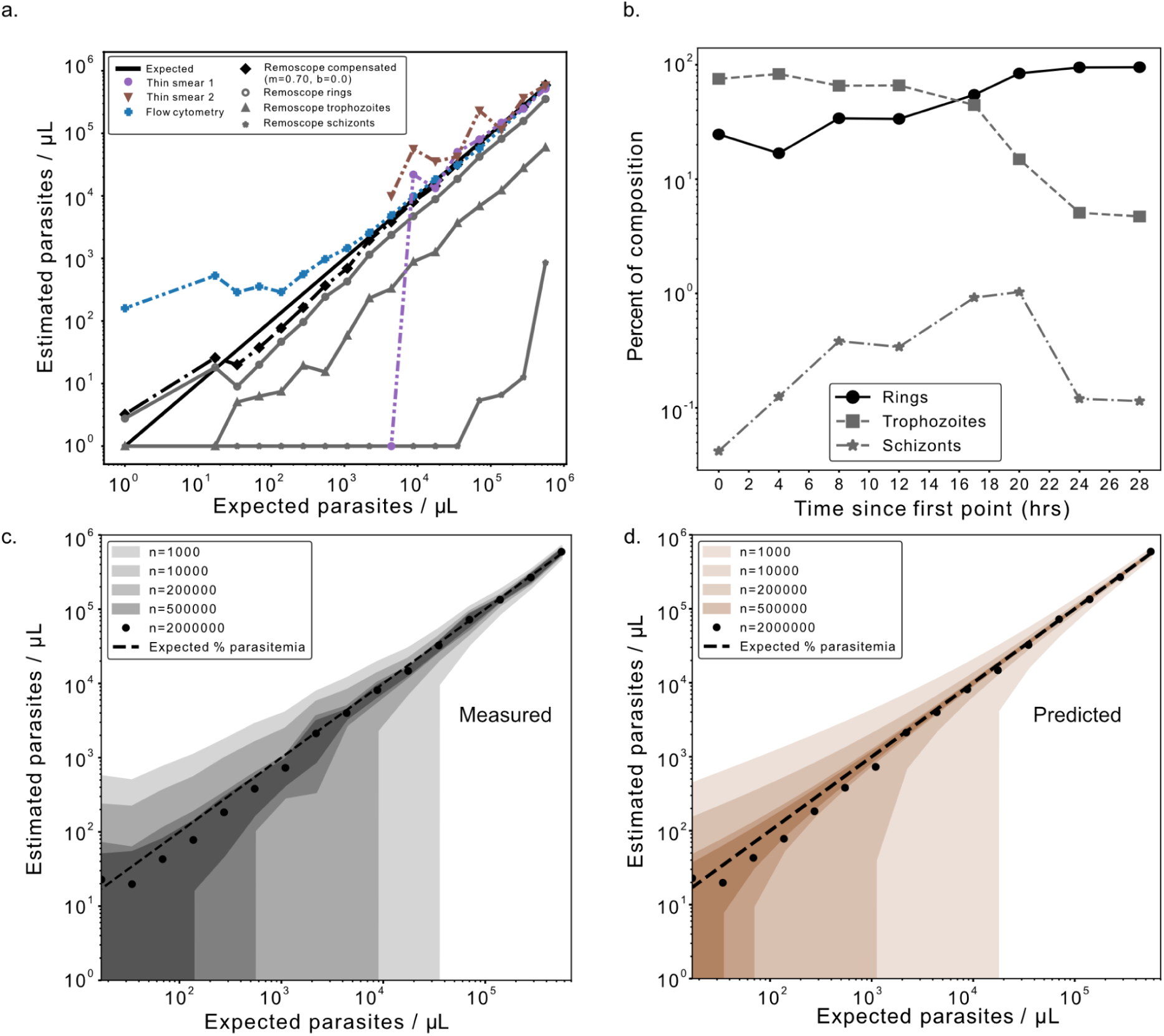
Remoscope performance on lab-cultured *Pf*. **a**. Life stage-specific parasitemia quantification over a 16-point serial dilution of parasite culture into uninfected, undiluted whole blood. Remoscope results are shown in black and gray markers. Thin smear microscopy counting ∼1,000 cells per point are shown for the first eight titration points only, and flow cytometry (2×10^6^ events per point) are plotted as blue ‘+’ signs. Remoscope data has undergone linear transformation with parameters m=0.7, b=0. **b.** Parasite life stage time course experiment over a continuous 28 hour period showing the assessed fractional compositions of each asexual life stage in the culture as a function of time. **c.** Uncertainty quantification of the titration data as a function of the number of cells analyzed. Contour regions span the overall mean +/- one standard deviation across data subsampled at the indicated number of cells. **d.** Theoretical prediction of the experimental uncertainty shown in **c**, as described in Supplementary Note 2.

**Fig. 4.**
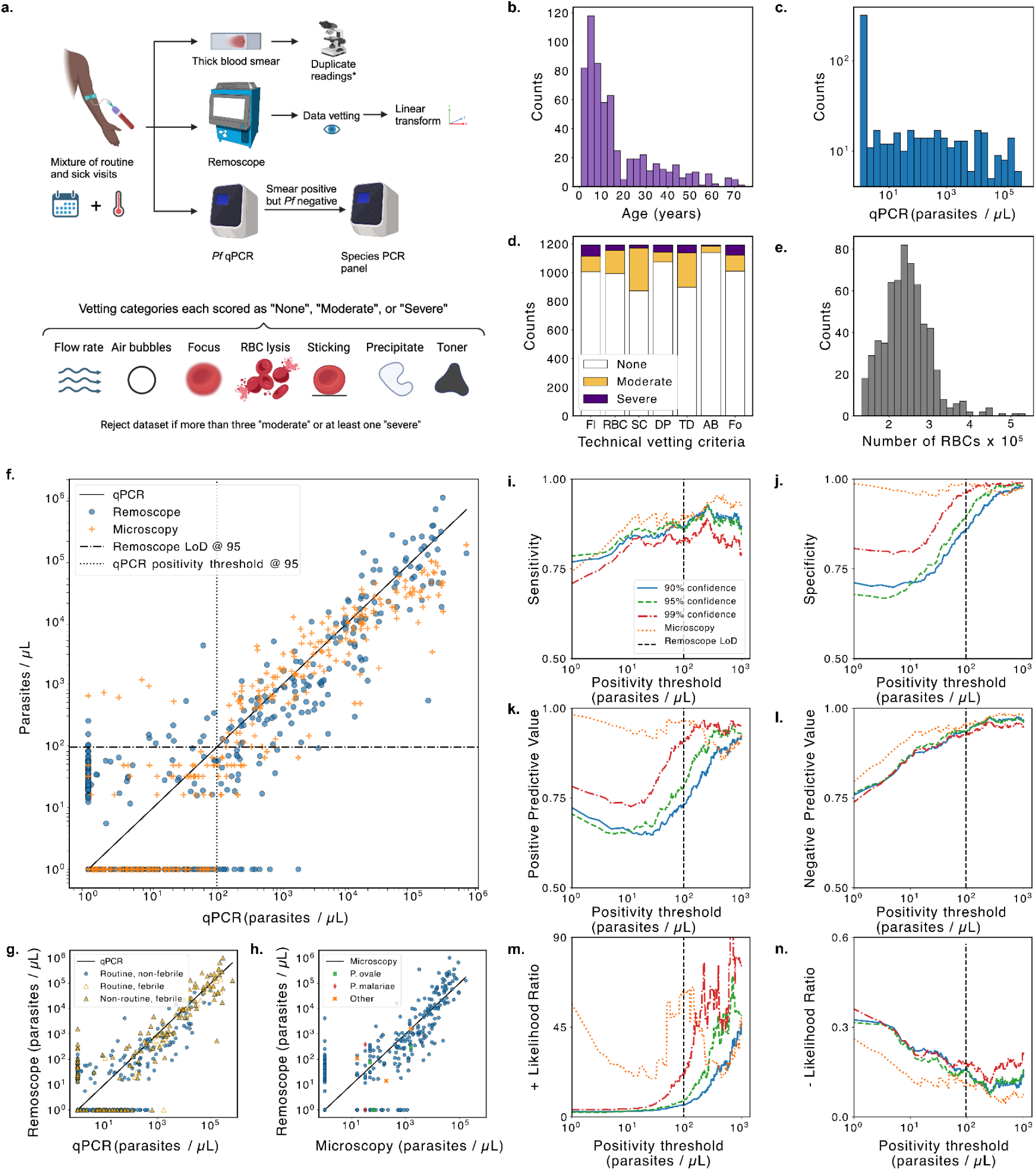
Clinical study in a Ugandan cohort using 10× diluted blood. **a**, Overall data collection and vetting strategy: venous blood samples were sent to Remoscope, qPCR, and thick blood smear microscopy with duplicate readings. Remoscope datasets underwent a quality vetting process to discard low quality datasets due to technical failures, according to seven vetting criteria. A linear transformation (slope and offset correction) was used to adjust for the average recall and false positive rates. **b,** Histogram of participant ages. **c,** Histogram of cohort parasitemias by qPCR. **d,** Stacked histogram of Remoscope dataset vetting results. “Fl” = Flow rate, “RBC” = RBC integrity, “SC” = Stuck Cells, “DP” = Diluent Precipitate, “TD” = Toner and Debris, “AB” = Air Bubbles, and “Fo” = Focus quality. **e,** Histogram of total red blood cell counts per experiment in the cohort (diluted blood), after filtering for N>132,735 cells (bottom 5% removed). **f,** Comparison of Remoscope and microscopy against qPCR. The horizontal dashed line indicates the computed Remoscope limit of detection (LOD). The vertical dotted line indicates the corresponding threshold on the qPCR axis. The two lines partition the plot into true negatives (lower left), true positives (upper right), false positives (upper left), and false negatives (lower right). **g,** The same Remoscope data as **f**, but plotted with delineation of visit type and febrility status. **h,** Direct comparison of Remoscope with microscopy. Non-*falciparum Plasmodium* species are denoted by distinct markers. ‘Other’ refers to samples testing positive by microscopy, but negative for both *Pf*-PCR as well as the species PCR panel. **i-n,** Respectively: Sensitivity, Specificity, Positive Predictive Value (PPV), Negative Predictive Value (NPV), + Likelihood Ratio, and - Likelihood ratio of the diagnostic test as a function of a common swept positivity threshold. Performance metrics were computed for three confidence thresholds: 0.9, 0.95, and 0.99. Linear transformation parameters were computed independently for each confidence threshold. In all subpanels, parasitemia values were clipped to one parasite/µL.

In this study, Remoscope used disposable flow cartridges that were fabricated in-house. We implemented quality control measures to reject defective devices and improve yield (Methods and Supplementary Figures 12 and 13). Nonetheless, failures such as poor channel alignment, debris, flow issues, lysis of red blood cells, or vacuum system leaks still occurred. Additionally, focus and flow rate control systems were also subject to occasional failures. Experiment runs were therefore vetted for various technical quality control parameters and scored in severity as either “None”, “Moderate”, or “Severe” (Figure 4a,d and Supplementary Figure 14). Runs exhibiting more than three “Moderate” issues or more than zero “Severe” issues were rejected from analysis.

### Performance assessments using cultured parasites

Cell culture of *Pf* (Methods) provided an efficient means of producing a large dataset of parasite images for initial model training and validation. With the ability to prepare parasites at parasitemia levels in excess of 10%, it was possible to image nearly 100,000 parasites in a single run, as well as vary the dominant life cycle stage. After initial data collection for annotation and model training, we conducted targeted experiments with cultured parasites in order to validate system performance.

First, we tested Remoscope’s limit of detection with a serial dilution of cultured parasites into uninfected whole blood. By spiking lab-cultured *Pf* into healthy, undiluted whole blood, we were able to vary parasitemia over a wide range, providing extrinsic validation of whether YOGO models trained on human annotations were able to accurately assess parasitemia levels (Supplementary videos 2-17). We saw a linear response over a 16-point serial dilution into whole blood from a starting parasitemia of 11.2% (∼560,000 parasites/µL equivalent), as determined by thin smear Giemsa microscopy performed by three independent readers (Figure 3). Total parasitemia and life stage breakdown decreased in concordance with dilution. An uninfected whole blood sample was included in order to assess the false positive rate in the absence of parasites. Each dilution point was additionally screened using conventional flow cytometry and thin blood smears for comparison. We acquired ∼2 million events per condition using flow cytometry and found that the sensitivity/specificity tradeoff was inferior to Remoscope’s: while aggressive gating on the fluorescence channel reduced background, the resulting recall was low. We therefore performed the same linear transformation for flow data, resulting in fit scaling parameters of m=0.7, b=0. Thin blood smears, although highly specific, were ineffective at low parasitemia due to inadequate sampling power when counting ∼1,000 RBCs per condition; therefore only the first eight titration points were counted manually as the remaining points had fewer than one parasite per 1,000 RBCs.

Compensated Remoscope measurements remained linear down to the 16th point at 0.00034% parasitemia (17.1/µL equivalent), with a root mean square ratiometric error of 0.29 over the 4.5 order of magnitude range. Model confidence thresholding was explored during analysis to vary the tradeoff between parasite recall and false positive rate (Supplementary Figure 15), resulting in an optimal value of 0.75. It can be noted that for rapid turnaround laboratory parasitemia assessments, the experiment can be set to terminate early upon achieving a target relative signal to noise ratio (Supplementary Figures 3 and 4). For example, at 1,000 parasites/µL (0.02% parasitemia) with undiluted blood, approximately three minutes of run time were required to reduce Poisson counting error below 10% relative uncertainty.

We investigated the effect of throughput (number of cells) on the empirical error bounds by subsampling the titration datasets at N=1,000, 10,000, 200,000, 500,000, and 2,000,000 cells per run (Figure 3b). We computed the standard deviation in estimated parasitemia levels between subsampled datasets, and plotted the mean +/- one standard deviation for each subsampling level. The effect of Poisson-limited counting statistics manifests as an inverse correlation between uncertainty bounds and number of cells. The basal uncertainty in the data quantifies the number of cells required in order to resolve a given parasitemia level, for both laboratory research and clinical settings. For example, a pan-analysis of 15 research studies (13) found that at 100 parasites/µL the probability of detection by microscopy was only 29.7%.

Finally, we assessed discrimination of parasite life stages by conducting a synchronized time course, using Remoscope to assess fractional compositions as a function of time. Throughout 28 hours (beginning with a trophozoite-dominated culture), the ratio of rings to trophozoites was seen to cross over, while schizonts (defined as having segmented intraerythrocytic merozoites; see Supplementary note 1 : Annotation guide) peaked thereafter. We note that while schizonts could be observed on Remoscope, recall was very low due to flow issues, presumably related to their increased stiffness and cytoadherent properties (32, 33). This phenomenon is explored further in the discussion section.

### Diagnostic accuracy in a Ugandan cohort

Testing Remoscope in a clinical setting was central to evaluating its utility as a diagnostic tool, complete with real world challenges. We deployed three Remoscopes for testing with a cohort of 500 individuals in eastern Uganda from January-September 2023. As part of the broader PRISM border cohort study (34), participants were scheduled for routine monthly blood draws regardless of symptoms (Figure 4a). Additionally, non-routine (sick) visits were conducted when participants presented at the clinic with fever. Both visit types were included in the study with a total of 407 routine (53 with fever) and 194 non-routine (all with fever) visits (Table 2 and Figure 4g). Venous blood draws were performed into 10 mL EDTA tubes, and processed within 24 hours on Remoscope using the 10× diluted blood assay with reconstituted diluent. Thick blood smears were fixed, stained with Giemsa, and read in duplicate by experienced microscopists. In the event of a disagreement, a third read was performed (as described in (34)). qPCR targeting the *Pf* multicopy conserved var gene acidic terminal sequence was performed on all samples (Figure 4a). Those with positive microscopy but negative qPCR results were additionally processed with a species PCR panel to screen for *P. ovale* and *P. malariae* (Figure 4a,h). Remoscope runs were quality vetted according to seven criteria as described above and in Supplementary Figure 14. Datasets with cell counts below 75,600 (the bottom 5 percentile) were also excluded from analysis due to inadequate statistical sampling power.

**Table 2.** Summary of participant visit types. Numbers include participant visits passing all quality control vetting criteria. Participants with SCD or testing positive for other *Plasmodium* species were also withheld from analysis.

| Visit types | Routine | Non-routine | Totals |
| --- | --- | --- | --- |
| Febrile | 53 | 194 | 247 |
| Non-febrile | 354 | 0 | 354 |
| Totals | 407 | 194 | 601 |

The cohort had an overall *Pf* prevalence of 67.9% by qPCR, 38.2% by traditional thick blood smear microscopy, and 45.4% by Remoscope (Table 3). By qPCR, the cohort’s parasitemia levels were evenly distributed across a logarithmic range spanning from less than 1 to over 600,000 parasites/µL (Figure 4c and Supplementary Video 18). One participant had sickle cell disease (SCD), five samples tested positive for *P. ovale*, and three tested positive for *P. malariae*, all of which were excluded from our diagnostic analyses, except Figure 4h. In all figures, the data were clipped along the x-axis to a value of 1 parasite/µL in order to focus on the range that is relevant to microscopy and Remoscope, as the qPCR limit of detection is approximately three orders of magnitude lower (35).

**Table 3.** Summary of diagnostic statistics by method and positivity threshold. All Remoscope statistics were computed using compensated (linearly-transformed) results and a YOGO model confidence threshold of 0.99.

| Method | Positivity threshold (parasites/μL) | Sensitivity | Specificity | PPV | NPV | PLR | NLR | F1 | MCC |
| --- | --- | --- | --- | --- | --- | --- | --- | --- | --- |
| Microscopy | 0 | 0.54 | 1.00 | 1.00 | 0.50 | inf | 0.46 | 0.70 | 0.52 |
| Microscopy | 1 | 0.74 | 0.99 | 0.98 | 0.80 | 56.61 | 0.26 | 0.85 | 0.75 |
| Microscopy | 95 | 0.90 | 0.99 | 0.97 | 0.96 | 62.96 | 0.10 | 0.93 | 0.91 |
| Microscopy | 200 | 0.92 | 0.98 | 0.94 | 0.97 | 44.64 | 0.09 | 0.93 | 0.90 |
| Microscopy | 300 | 0.93 | 0.97 | 0.91 | 0.98 | 28.10 | 0.07 | 0.92 | 0.89 |
| Microscopy | 500 | 0.93 | 0.96 | 0.87 | 0.98 | 21.78 | 0.07 | 0.90 | 0.87 |
| Remoscope | 0 | 0.55 | 0.78 | 0.85 | 0.44 | 2.46 | 0.58 | 0.67 | 0.30 |
| Remoscope | 1 | 0.71 | 0.81 | 0.78 | 0.74 | 3.67 | 0.36 | 0.74 | 0.52 |
| Remoscope | 95 | 0.83 | 0.96 | 0.91 | 0.93 | 21.63 | 0.18 | 0.87 | 0.81 |
| Remoscope | 200 | 0.87 | 0.99 | 0.96 | 0.95 | 63.85 | 0.13 | 0.91 | 0.89 |
| Remoscope | 300 | 0.85 | 0.99 | 0.96 | 0.95 | 63.87 | 0.15 | 0.90 | 0.87 |
| Remoscope | 500 | 0.83 | 0.98 | 0.93 | 0.95 | 48.54 | 0.17 | 0.88 | 0.85 |

Remoscope’s 10× diluted blood assay resulted in an average of 243,623 RBCs per run in the cohort (Figure 4e), and closely tracked the results of expert-level thick smear microscopy performed in duplicate (Figure 4f,h). In Figure 4f, we plot Remoscope and microscopy results vs. qPCR. On average, the Remoscope diluted blood assay scans a similar number of cells as viewed by a technician performing microscopy, where the number of asexual parasites per either 200 or 500 leukocytes is counted (the latter for low density samples), with an assumption of 8,000 leukocytes/µL. Thick smear microscopy therefore inspects an equivalent of 125,000-312,500 RBCs, using an average RBC concentration of 5×10^6^/µL. Fundamental counting statistics place similar bounds on the performance of Remoscope as microscopy, with microscopy benefitting from the added specificity of stain, and duplicate technician readings.

Since qPCR assays for *Pf* have a limit of detection at least three orders of magnitude lower than either microscopy or Remoscope (estimated at 22 parasites/mL (35, 36)), direct comparison of binarized diagnostic outputs with qPCR results in low sensitivity values. Furthermore, detection of sub-patent infections in a high transmission / high prevalence setting is of limited clinical value when the prevalence regardless of symptoms is 69%. We therefore considered a swept qPCR positivity threshold, varying from 1 to 1000 parasites/µL in order to quantify diagnostic performance statistics (Figure 4 i-n) and tabulated a subset in Table 3. We computed sensitivity, specificity, Positive Predictive Value (PPV), Negative Predictive Value (NPV), + Likelihood Ratio (+LR), and - Likelihood Ratio (-LR) with respect to the common swept positivity threshold.

Remoscope’s limit of detection (LoD) was computed as the mean plus three standard deviations of all qPCR negative samples, resulting in a LoD of 95.1 parasites/µL. By this metric, microscopy had no false positive results. While this suggests an LoD of 0, the method cannot detect arbitrarily small parasitemia, and the smallest detectable level is then limited by the probability of detection. Using the method outlined by Armbruster and Pry (37), we computed the Limit of Blank (LoB) for each method, and subsequently the fraction of clinical data points exceeding each method’s LoB as a function of qPCR value (Supplementary Figure 16).

The YOGO model confidence threshold also influenced diagnostic performance, and we computed the metrics for three values: 0.9, 0.95, and 0.99. For example, Remoscope’s limit of detection continued to improve with increasing model confidence threshold up until a value of 0.99 (Supplementary Figure 15) due to the reduction of false positives and improvement of specificity. The optimal selection of model confidence and positivity threshold is influenced by what is important for clinical decision making: here, we opted to use a value of 0.99 model confidence threshold in order to prioritize specificity and limit of detection. In Table 3 and Figure 4 f-h, a model confidence threshold of 0.99 was employed throughout.

We also computed integrated performance metrics such as the F1-score and Matthews Correlation Coefficient (MCC) (38) as a function of positivity and model confidence thresholds (Supplementary Figure 17). We achieved a maximal MCC value of 0.9 at a model confidence threshold of 0.98 and positivity threshold of 300 parasites/µL.

Remoscope exhibited similar sensitivity and NPV as expert thick smear microscopy across a wide range of positivity thresholds (Table 4 and Figure 4i,l). While microscopy remains superior in specificity and PPV, there is robust evidence that Remoscope’s false positive rate is more than tenfold lower with undiluted blood (Figure 3 and Supplementary Figure 18).

**Table 4.** Agreement level between microscopy and Remoscope. A breakdown of the agreement between Remoscope and microscopy for binarized diagnostic results. For Remoscope, a positivity threshold equal to its limit of detection was used (95.1 parasites/µL), whereas for microscopy a threshold of 0 parasites/µL was used, in concordance with existing clinical practices.

| Binarized result summary | Microscopy negative | Microscopy positive |
| --- | --- | --- |
| Remoscope negative | 368 | 66 |
| Remoscope positive | 10 | 160 |

Fever status of participants is stratified in Figure 4g and Supplementary Figure 19), showing bias toward higher parasitemia with fever. In Figure 4h, we plot Remoscope vs. microscopy and include other *Plasmodium* species, noting that the YOGO model, untrained on these species, exhibited a partial sensitivity to the parasites. See Supplementary Figure 20 for example images of non-*falciparum* parasites.

### Determination of half-maximal effective drug concentration (EC50) with Remoscope

An important laboratory application is the determination of *in vitro* drug efficacy in cultured parasites, where the ability to quickly and easily quantify stage-specific parasitemia levels is crucial. For life stage assessment, thin blood smears represent the gold standard, while overall parasitemia assessment is commonly achieved with fluorimetry, or for higher sensitivity assays, flow cytometry (39).

We performed a titration of chloroquine (CQ) into *Pf* cultures, and tested each concentration by Remoscope and flow cytometry after a 72 hour incubation with the drug (Figure 5). Sigmoidal fits to the Remoscope and flow cytometry “all-stages” data points yield similar EC50 values of 211 nM and 191 nM, respectively. We found that Remoscope’s stage-specific assessments of parasitemia and exhibition of images of cell morphology provided insight into the drug’s efficacy and mechanism of action. CQ is known to inhibit the parasite’s heme polymerase, preventing formation of hemozoin crystals, and promoting accumulation of toxic hematin (40, 41). We observed morphological changes associated with drug treatment (Figure 5b). For example, there was a notable lack of hemozoin crystal formation in the presence of inhibitory concentrations of CQ. Ring stages were also seen to collapse, forming solid puncta in place of the canonical annuli (42). YOGO was not trained on images of drugged parasites. Nonetheless, we observed a loss of parasite detection with increasing drug concentration. With replication disrupted by the drug, the overall number of parasites in the sample was lower. Also, drugged parasites lacked the ability to form hemozoin crystals, and exhibited altered appearances. In Figure 5, we showed example images from the ring and trophozoite categories under low and high drug concentrations, and in Supplementary Figure 21, we show the effect on model confidence score.

**Fig. 5.**
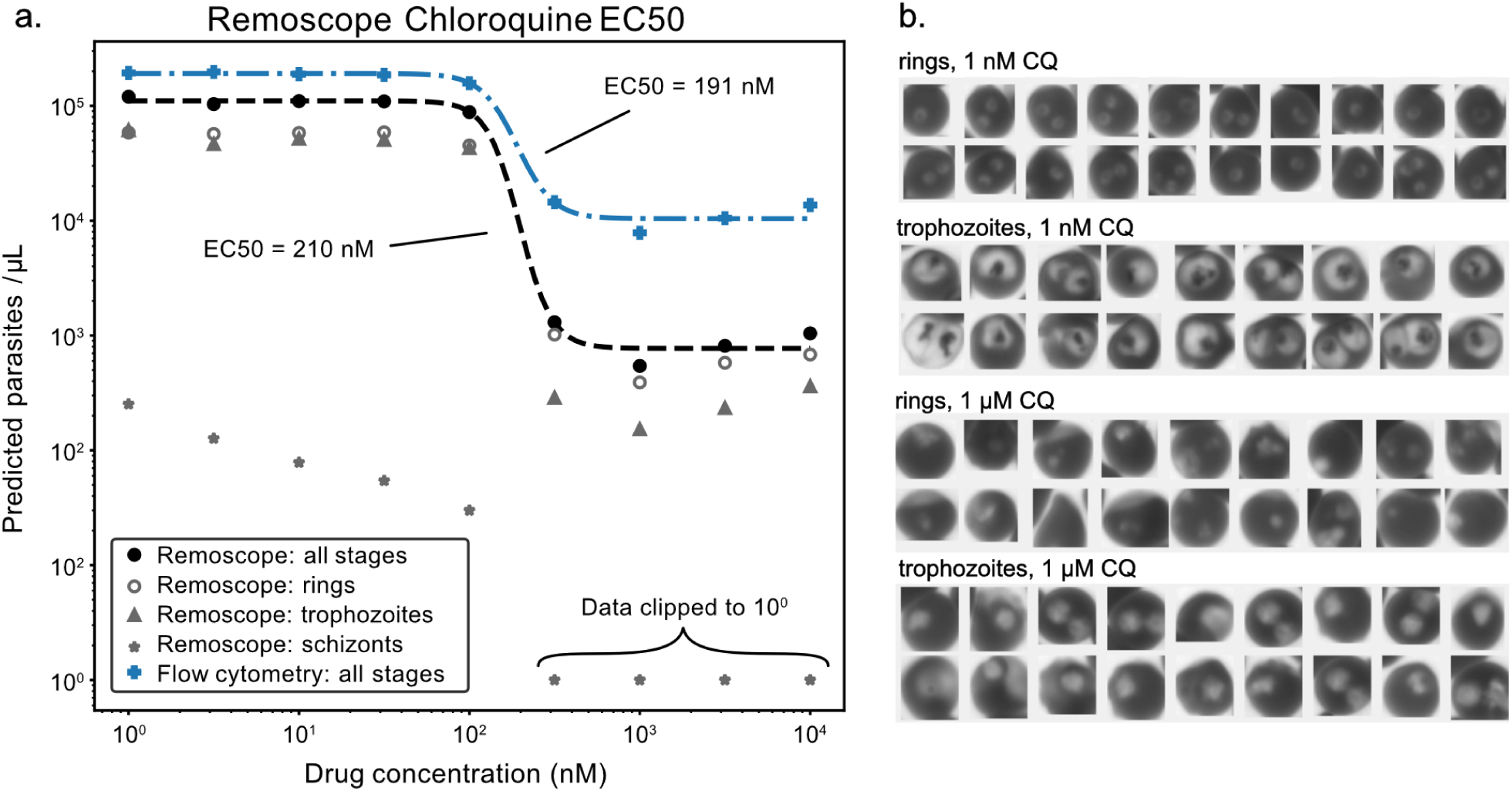
EC50 assessment of chloroquine (CQ) effectiveness. **a,** Stage-specific parasitemia assessments as a function of CQ concentration after a 72 hour drug exposure under cell culture conditions. Remoscope overall parasitemia is shown in filled black circles, while rings, trophozoites, and schizonts are shown in gray rings, triangles, and stars, respectively. Schizont counts above 100 nM drug concentration were zero, but set to 1 for display on the logarithmic scale. Flow cytometer data of the same samples is shown for comparison. Dashed lines represent sigmoidal curve fits to both all-stages datasets. **b,** Montages containing example rings and trophozoites from the 1nM and 1µM conditions, respectively.

### Extension to generalized blood morphological phenotyping

In this work, we trained a convolutional neural network (CNN) to identify subtle image intensity patterns due to the presence of *Pf* parasites at different life stages. It follows that other major RBC morphologies such as sickled cells (SCD), echinocytes (variety of causes), stomatocytes (present in liver disease), target cells (can result from thalassemias, alcoholic liver disease, and SCD), spherocytes (asplenism), microcytes (occur in various anemias), macrocytes, hypo/hyperchromia, agglutination, and more (31), could also be classified through appropriate model training. Supplementary video 19 and Supplementary Figure 22 show the blood from a Ugandan participant with SCD, displaying sickled cells, poikilocytes, target cells, and other non-discocytes. Supplementary video 20 demonstrates the phenomenon of hemagglutination, occurring when spiking parasites cultured in RBCs of one donor blood type into healthy whole blood from a second, non type-matched donor. Hemagglutination can occur clinically in cold agglutinin disease (43), or in hemagglutination assays. Supplementary Video 21 demonstrates abnormal RBC morphologies observed in a Ugandan participant who was also positive for *Pf*. Target cells, echinocytes, and stomatocytes were observed. To our knowledge, no other system can perform real time fully-automated morphological analysis on millions of blood cells without sample preparation and/or manual inspection of smears. Furthermore, blood smear inspections often contain spurious results due to the presence of lipid contamination, pH changes, poor technique, or other reagent problems during smear preparation (31), whereas Remoscope processes whole blood without introducing morphogenic agents.

## Discussion

In this proof of concept study, we have introduced the Remoscope platform and demonstrated its utility as a diagnostic instrument for malaria in a clinic in eastern Uganda, comparing performance to thick smears and qPCR. The cost of the instrument and disposable flow cartridges are within the means of endemic countries: material cost of the prototype instrument is on the order of $2,200 USD (a mass-manufactured version will cost significantly less), and while the prototype flow cartridges costed ∼$3 USD (including labor) for this study, a ∼$1 USD commercially-made flow chamber is now ready for use at the time of this manuscript’s submission, and scaling beyond research quanitities will further reduce the cost per test. We evaluated Remoscope’s use in laboratory research applications, by showing that parasitemia quantification is linear across a 16-point dilution series. We also showed that treatment of *Pf* parasite cultures with CQ imparted sufficient morphological changes to evade detection by a custom trained CNN, enabling EC50 drug evaluations *in vitro*.

In comparison with traditional Giemsa microscopy (using qPCR as the gold-standard), Remoscope exhibited similar sensitivity and PPV, but trailed in specificity and NPV. However, it should be noted that the manual smear readout in this study was expert-level and performed in duplication with research-grade reagents, resulting in performance unlikely to be achieved in most lower-tier health facilities. In fact, it has been shown that even at 100 parasites/µL the average probability of detection by microscopists in clinical settings globally is less than 30% (13). In comparison with these data, Remoscope’s diluted blood assay already exceeds the average standard of care. Examination of Remoscope false positives revealed the primary causes to be deformed RBCs and debris stuck to the flow cartridge surface. The lyophilized reagent used in the blood diluent suffered from an intermittent degree of insolubility upon reconstitution after transport and storage on site. Despite vetting efforts (Figure 4a), it is likely that inconsistent diluent reconstitution affected RBC morphologies, given their sensitivity to the local biochemical environment (30, 31). With undiluted blood, a tenfold improvement to the limit of detection is anticipated, along with concomitant improvements to all other performance metrics. These benefits of using undiluted blood have been validated *in vitro* with a parasite titration, in which we were able to detect concentrations as low as 17.1 parasites/µL, as well as demonstrate an average false positive rate of 3 parasites/µL based on a small cohort of healthy donors (Supplementary Figure 18). Future clinical studies will aim to validate these advantages in performance and workflow.

An additional consideration is the rigidity and cytoadherence properties of *Pf* schizonts (44, 45), which renders them more likely to be trapped inside our prototype flow cartridge upstream of the imaging window, failing detection. Our life stage analysis (Figure 3) showed a substantial decrease in mature schizont detection efficiency. Fortunately, since the vast majority of clinical *Pf* samples contain primarily rings as more mature stages are not circulating, this phenomenon is not expected to impact clinical diagnostic performance. We also observe adherence of WBCs to the flow cartridge due to its 4.5 µm channel height, straddling the requirement that RBCs lay flat for imaging while not clogging larger cell types. In some cases, the diagnosis was hampered by high WBC counts and rejected during the vetting process. Future flow cartridge designs will employ hydrodynamic flow focusing (46) to confine blood to a freely-flowing monolayer while permitting unbiased flow and counting of all cell types including late stages of *Pf*, while also enabling a wide array of other low-cost blood-counting applications beyond the detection of malaria.

In this work, only models to detect *Pf* were trained as it is the species responsible for the large majority of malaria cases and deaths worldwide (1) and is amenable to lab culture. Expanding into other species is a logical continuation of this work, as there are regions where other *Plasmodium* species such as *P. vivax* are dominant (1), and identification of species information is crucial to informing treatment as well as epidemiological tracking in certain geographical regions. In our clinical cohort, we observed a small number of *P. ovale* and *P. malariae* cases (Supplementary Figure 20). Our model achieved partial cross-species detection, despite no training examples of non-*Pf* parasites (Figure 4h). Future work will involve clinical data collection and model training on additional *Plasmodium* species by launching clinical studies in other regions.

The image data from this study will be made freely available as a resource to the community. To our knowledge, this resource will represent the largest collection of label-free live parasite images ever produced, consisting of 14.4 million images and 399 million blood cells from healthy donors, infected patients, and cultured parasites. This collection of images provides a rich set of data for machine learning and evolving AI approaches. In addition to further development for malarial diagnostics, future efforts will expand the application space of high-throughput, low-cost scanning of blood morphology to other bloodborne pathogens (such as Trypanosomiasis or Babesiosis), hereditary blood disorders, complete blood counts, as well as morphological fingerprinting of RBC ensembles using more generalized machine learning frameworks. These expansions in throughput and model complexity will be well-supported by a favorable trend in portable edge computing hardware performance (47), without increasing cost.

## Methods

### Ethics statement

All research was conducted in accordance with local regulatory requirements and approvals. Blood was drawn at UCSF from healthy adult volunteers providing written consent, for use in sustaining malaria cultures and imaging of healthy blood under UCSF IRB#10-02381.

Approval for the Ugandan cohort study was obtained from the Makerere University School of Medicine Research and Ethics Committee (REF 2019-134), the Uganda National Council of Science and Technology (HS 2700), the London School of Hygiene & Tropical Medicine Ethics Committee (17777), and the University of California, San Francisco Committee on Human Research (257790). Written informed consent was obtained for all participants prior to enrollment into the study. Funding was provided by the National Institutes of Health as part of the International Centers of Excellence in Malaria Research (ICMER) program (U19AI089674).

### Remoscope

#### Fabrication

All instruments were assembled in-house by CZ Biohub SF research staff. Here we present an overview of the instrument design.

#### Structural overview

The mechanical support structure of the instrument is assembled from custom machined aluminum plates forming a mounting scaffold for other elements of the system, including: a pneumatic module for controlling vacuum, a microscope tube with coarse focus mechanism, a motorized microscope fine focus mechanism, top plate surface with protective lid, a Condenser And Pneumatic (CAP) interface for coupling light and vacuum into the flow cartridge, an onboard computer (Raspberry Pi 4b) and hardware accelerator (Intel Neural Compute Stick 2), a user touch screen (Canakit RSP-DISPLAY), custom electronics, forced air cooling, and internal wiring. The entire system is protected by a tight-fitting, custom sheet metal enclosure. Sorbothane feet are used to passively isolate the system from table vibrations.

#### Flow cartridge design

Flow cartridges are 22×40 mm in size and constructed from laser-cut acrylic, a standard No 1.5 glass coverslip, with laser printer toner used to define the flow channel path and thickness, which was 1 mm wide and 4.5 µm in height. For more information, see the CAD model above and the fabrication details below.

#### Optical design

The near-UV brightfield microscope comprises a 405 nm LED light source coupled to a plastic homogenizer rod (see CAP module below), a finite conjugate image-forming objective, and a monochrome machine vision camera. We previously found that the excitation wavelength of 405 nm works well for parasite classification due to a lower dependency on focal error and the fact that it coincides with the maximum absorption peak of hemoglobin (22). Crucially, the LED source provides adequate illumination intensity in order to use sufficiently short exposure times required for capturing images of fast-moving cells without inducing motion blur. A monochrome, global shutter machine vision camera is used to capture images of the moving cells at a fixed rate of 30 frames per second.

#### Onboard computation

Hardware control, process automation, and some image-based computations are performed by a Raspberry Pi 4B (8GB) single board computer coupled to a custom printed circuit board (PCB). An Intel Neural Compute stick 2 (NCS2) is used for all real-time inferencing tasks, including running YOGO, the deep-learning model that classifies cells. A portable SSD for saving raw images is connected to the Raspberry Pi via a USB hub.

#### Sample mounting stage and focus mechanism

The fine focus mechanism employs a spring-coupled flexural mechanism to deflect the flow cartridge holder (stage) with respect to the objective. The mechanism employs a displacement dividing system, where the motion of a $35 linear stepper motor (Portescap 20DAM10D1B-K) is divided by a 40:1 ratio of spring constants, resulting in a ∼0.3 µm minimum displacement when driven from a half-stepping motor controller. The linear stepper motor’s range of motion was set by a single-ended limit switch and limited to a total motion of 900 steps, resulting in a nominal range of 286 µm of fine focus travel. A spring clip is used to retain the flow cartridge in place and maintain lateral alignment of the imaging window with respect to the objective during the experiment.

#### Pneumatic control module

Vacuum-driven flow of the blood cells is achieved by connecting the flow cartridge’s exit port to a servo-controlled pneumatic piston (SMC Pneumatics NCMB044-0100). The linear position of the piston is adjusted by a servo motor (Pololu HD-1810MG) coupled to a linkage mechanism converting the servo’s rotational motion into linear motion of the piston. Imaging-based closed-loop feedback control is used to adjust the vacuum pressure to stabilize the velocity of the moving cells. An absolute pressure sensor (Honeywell MPRLS0025PA00001A) connected to the vacuum line is used to monitor vacuum pressure and detect leaks. The pneumatic module is enclosed in a 3D-printed box and mounted to the instrument’s base plate.

#### Condenser And Pneumatic interface (CAP) module

Due to space constraints on top of the flow cartridges, the excitation LED and pneumatic seal are integrated into a single compact module. Within the assembly, a UV-resistant acrylic light guide (Amazon B07W6ZRJB6) couples light from the LED down to the sample, simultaneously homogenizing the spatial and angular emission profiles. A monolithic 3D-printed structure mounts these elements as well as a pneumatic connector which interfaces an internal conduit with external polyethylene tubing going to the vacuum piston. The internal conduit routes directly above the exit port of the flow cartridge, where it meets a custom-cut silicone gasket that mates with the exit port on the flow cartridge. A set of four rare earth magnets (K&J magnetics DH16-N42) provide force to snap the module in place as well as exert the necessary pressure to form a seal on the flow cartridge’s exit port. LED wiring and pneumatic tubing route from the CAP module to the instrument’s top plate along a polymer ribbon, keeping them tidy and strain-relieved.

#### Electronics design

A full CAD model with schematics, PCB layout, fabrication files, and bill of materials for all custom electronics in the system are provided under an open source license (Supplementary Figures 23, 24, and https://github.com/czbiohub-sf/remo-pcbs). Custom printed circuit boards (PCBs) serve as an interface between the Raspberry Pi 4b (RPi) and all the active hardware elements in the system. A main control PCB connects to the 40-pin GPIO header on the RPi, which provides communications as well as power to the RPi. The main PCB also houses driver circuits for the focus motor, pneumatic servo, and excitation LED, as well as routes signals from the pressure sensor, focus limit switch, and safety interlock switch to the RPi, while also providing power for two positive displacement fans that cool the instrument’s interior. A secondary ‘daughter’ PCB distributes power and signals to various devices including the pneumatic servo, upper fan, limit switch, and focus motor. To navigate global supply constraints, we also housed both the 405 nm UV excitation LED and pressure sensor elements on custom PCBs. A combination of off-the-shelf and custom-built cabling solutions serve to connect both the main PCB and daughter PCB to the various hardware components around the system.

#### Software design

Custom, Python-based software with a Graphical User Interface controls operation of the instrument, including overall experiment workflow and real-time control of system hardware. A finite state machine controls progress through metadata entry, calibration steps, experiment progress, and handles errors. During the experiment, a function called at regular intervals acquires images and allocates time-consuming tasks such as YOGO object detection inferencing, flow rate estimation, and raw data saving to multithreaded queues. During the experiment, closed loop feedback is used to stabilize focus, flow rate, and brightness of the excitation LED, while additional variables such as the density of cells are monitored in real time. Focus stabilization is done using a custom CNN that estimates the focal error magnitude and direction without the need to pause data acquisition, which we refer to as Single Shot Autofocus (SSAF) since it does not require a focus sweep (48, 49). If user intervention is required to correct an error, the experiment is paused and the user informed of the issue. After termination of the run, a summary report including parasite counts and thumbnail images is generated and saved to disk. Every image of the blood is saved to the SSD as a lossless PNG file, alongside experiment metadata, per-image metadata, and the summary report.

#### Onboard image processing

In order for Remoscope to be able to process live images at close to 30 frames per second raw images need to be cropped to ¼ height. The target flow rate was set such that each ¼ cropped image would contain, on average, a new set of unique cells, with the full raw image from the camera containing on average four images of each unique cell. Therefore, the operation of ¼ cropping nominally did not reduce the cellular throughput of the system. Data for the publication were processed offline to make use of the most up to date trained models and analysis tools, using the full raw images. Parasitemia and error estimates are computed based on the estimated number of unique cells in the dataset, using the computed flow rate from pairwise sequential images.

#### Disposable flow cartridge fabrication

Disposable flow cartridges are fabricated in-house in sheets of 32 devices by a custom lamination process. Briefly, channel layer patterns are printed on silicone release liners (Slick Paper Silicone Release Paper) using a standard laser printer and transferred to laser-cut acrylic plastic sheets by a heated lamination step (the heat and pressure transfers the printer toner to the acrylic surface). Cleaned No 1.5 glass coverslips are subsequently aligned to the resulting toner-patterned acrylic sheets using a custom jig, and tacked in place with Loctite 420 cyanoacrylate adhesive. The tacked assembly is then laminated, serving to compress and bond the laser printer toner to the glass and plastic layers, resulting in a 4-6 µm flow channel height in the imaging area. Dithering of the toner in the imaging area is used to fine-tune the channel height in the imaging area, as grayscale ink values are compressed slightly more by the lamination. The devices are then sealed and strengthened by a second application of Loctite 420 and allowed to cure. Throughout the process, care is taken to avoid particulate contamination inside the channel, in the absence of a cleanroom environment, by using a filtered dry air gun on internally-facing layers before bonding steps. Corona treatment of the flow cartridges is used to enhance hydrophilicity and reduce cell adherence, permitting continuous flow of undiluted whole blood.

#### Flow cartridge quality control and tracking system

Devices are quality-control checked using two imaging processes. First, the sheets are scanned using an office document scanner and images analyzed to confirm XY alignment of the toner features to the laser-cut patterns. Next, the sheets are imaged using a custom interference fringe detection jig in order to assess the flow layer flatness, lamination quality, and glue wicking. The jig comprises fluorescent tube lighting, a diffuser, a large-area beam splitting mirror, and a Raspberry Pi HQ camera (Supplementary Fig 12). Interference fringe images are used to visualize the flow channel thickness profiles and to detect devices with either insufficient bonding, glue leakage, or delamination. Devices are subjected to corona treatment (Electro-technic products BD-20) for 10 seconds per chamber, at a distance of 1 cm, in order to increase channel surface hydrophilicity. Flow cartridges failing either QC modality are rejected and passing devices are packaged into cardboard freezer boxes.

### Model Training

#### Image annotation

Prior to extrinsic validation, annotation criteria were determined by manual inspection of thousands of raw Remoscope images of healthy blood and cultured parasites of different life cycle stages. A full annotation guide is provided (Supplementary Note 1), outlining the image features used to identify each object class, as well as the entire annotated dataset used for YOGO training (Supplementary data 1).

Image annotations were conducted in roughly three phases as YOGO performance improved. First, Cellpose (28) was used to produce coarse-grained annotations consisting of RBC bounding boxes without knowledge of infection status. Cellpose bounding boxes were imported into Label Studio (community edition), where manual annotation was used to correct missing or incorrect bounding boxes as well as begin the process of labeling infected cells (ring, trophozoite, schizont, gametocyte), white blood cells (WBCs), and miscellaneous objects (large platelets and debris). Training data in standard YOLO format was exported from Label Studio and used for YOGO training, which was performed in parallel with the annotation effort to monitor performance increases with dataset size. The second phase began once YOGO bounding box accuracy exceeded Cellpose output, as well as recapitulating some detection of other categories, allowing for more efficient assignment of labels to data. From this point on, YOGO was exclusively used for pre-labeling (to increase efficiency), and human annotation was performed only for error correction.

Thumbnail classification was used to augment the dataset more efficiently as follows: cropped “thumbnail” images of YOGO-labeled cells were exported to disk into sorted directories. Human annotation was conducted by moving incorrectly sorted images into correct directories. Upon completion, a python script was used to parse the sorted directory and retrain the network. This thumbnail method allowed more targeted export criteria to be used (such as model confidence thresholding or class composition) to only display relevant cells for annotation.

Further refinements were conducted by validating YOGO performance on intermediate titration experiments in order to identify sources of false positives (for example debris, platelets, or flow-distorted blood cells). Images from the identified failure points were annotated and added to the training data. The same procedure was later applied to healthy (defined as having < 1 parasites/µL by qPCR and 0 parasites/µL by microscopy) and infected (the 20 highest parasitemia participants by qPCR) subsets of clinical data from the Ugandan cohort: 1% of the 20,000 images acquired during a full experiment were set aside for model training and not used for testing or performance evaluation in the results presented above.

#### YOGO training

YOGO was trained on NVIDIA A100 GPUs at an on-site high performance computing cluster. Parallelized training was facilitated using the Pytorch Distributed Data Parallel module(50). Weights and Biases (51) was used for run tracking and hyperparameter search. An individual training run was complete once the model was trained on a set number of epochs. The Single Shot Autofocus CNN was trained using the same infrastructure and tools as YOGO.

#### Vetting process for data quality control

Datasets for the clinical study in Uganda underwent a manual vetting process to remove low-quality data. All videos from the study were exported to a directory, and manually inspected without knowledge of parasitemia or other participant metadata. The following categories were scored independently on a three-point scale (‘normal’, ‘moderate’, and ‘unusable’): Flow issues, RBC abnormalities, cell sticking, diluent insolubility, debris on surface, air bubbles, or focus issues. If one or more categories were marked as ‘unusable’, or if three or more were marked ‘moderate’, the dataset was not used. Otherwise, the data was used. Supplementary Figure 14 shows a graphical representation of the vetting status results for all clinical datasets.

#### Parasite culture

*Pf* strains W2 and NF54 were cultured in either T-25, T-75 or T-150 flasks depending on the scale of experimentation. Cultures were grown in RPMI_c_ supplemented with 0.5% Albumax II (Gibco Life Sciences 11021029), 2g/L sodium bicarbonate, 0.1mM hypoxanthine, 25mM HEPES (pH 7.4), and 50 µg/L gentamicin. Cells were maintained at 2% hematocrit in a gas and temperature-controlled environment set to 37°C, 5% O_2_, and 5% CO_2_. To avoid parasite overgrowth and death, cultures were split and passaged daily to maintain a parasitemia between 1-5%.

#### Blood smear preparation and counting

500 µL of culture were centrifuged, and 10 µL of the pellet was used to create a thin smear on a glass specimen slide. To visualize iRBCs, slides were dried, fixed in methanol, and stained for 15 minutes using 2% Giemsa. The slide was then observed at 100X magnification on an upright light microscope.

#### Enrichment of specific life-stages

The *Pf* life cycle under culture conditions contained a mixture of life cycle stages, depending on the degree of synchronization. To aid in the annotation effort, specific stages were enriched in order to increase the density of image data.

##### Sorbitol synchronization – enrichment of ring stage parasites

50mL of culture at >10% rings was moved into a 50mL falcon tube and centrifuged for 5 minutes at 1500 rpm with low brake. All but 5mL of media and RBCs were aspirated, then mixed gently with an equal volume of warm 5% sorbitol. The tube was shaken slowly to mix every 5 minutes for 15 minutes and then centrifuged again for 5 minutes at 1500 rpm with low brake. The media and sorbitol were aspirated completely, and the pellet was resuspended in the starting volume of RPMI_c_.

##### MACS purification – enrichment of trophozoite and schizont stage parasites

MACS purification was carried out as described previously (52). Cultures were first synchronized at ring-stage and then MACS purified once at a >10% parasitemia in the late-trophozoite stage. 50mL cultures were centrifuged at 1500 rpm for 5 minutes with low brake, and the supernatant was aspirated till 20mL remained. 4 LD columns (Miltenyi Biotec 130-042-901) were set up on the magnetic stand, washed twice with RPMI_c_ and then had 5mL of resuspended culture allowed to drip through. When around 1mL remained in the column, an additional 5mL of RPMI_c_ was added to wash the columns. The columns were then taken off the magnetic rack and placed into a 15mL conical tube, where they were eluted with 5mL RPMI_c_. The collected eluate was then used to expand the culture or for immediate use in experiments.

#### Titration and flow cytometric analysis of parasitemia

Parasitemia assessment was performed on a serial 2-fold dilution of in vitro *Pf* culture into type-matched fresh, healthy, whole blood. Identical aliquots of culture run on Remoscope were also assessed by flow cytometry. Cells were diluted to 0.5% parasitemia and fixed in 2% Paraformaldehyde for 1 hour at room temperature in a 96 well U-bottom cell culture plate. The fixed cells were then stained overnight in 50nM YOYO-1 and run using a high throughput system attachment on a BD FACSCelesta flow cytometer. Each sample was run in duplicate, with a collection threshold of 2 million events. Gating and parasitemia calculations were then carried out using FlowJo.

#### Gametocyte induction strategy

Freshly thawed strains of NF54 were utilized to ensure maximal capacity of gametocytogenesis, and blood less than one week old was used to minimize lysis of cells during the induction assay. Cultures were grown in RPMI_C_ and tightly synchronized using sorbitol and MACS purification. On day -3, a T-150 was set up with 2% trophozoites at 5% hematocrit at a total volume of 25mL. On day -2, hematocrit was lowered 1:2 by adding 25mL of fresh media. On day -1, only half the media was changed out with fresh RPMI_C_ and the culture was split to ∼2% trophozoites. From this day forward, daily media changes were carried out with no addition of new blood. The culture was smeared and inspected each day, until Stage IV or V gametocytes were observed.

#### Time course acquisition of life stages

To capture a continuum of stages across a generation of the *Plasmodium falciparum* life cycle, a time course experiment was conducted on a high parasitemia culture. The culture was twice synchronized by sorbitol treatment, and the first time point was collected at the early trophozoite stage. Samples were prepared by spinning down culture at 2% hematocrit, resuspending the pellet in an equal volume of diluent to bring the hematocrit to ∼50%. Images were acquired in full length runs every four hours for 28 hours.

#### Chloroquine dose response assay

Cultures were initiated at 0.5% hematocrit and 0.8% rings, and 190 µL was dispensed into rows of a 96-well U-bottom culture plate. A √10 dilution series beginning at 200 µM and ending at 20 nM was made in 1X PBS: 10 µL from each stock well was transferred into the culture wells for a 20-fold dilution in concentration. Cultures were incubated at 37℃ for 72 hours, after which parasitemia at each dose was quantified on both Remoscope and flow cytometry. To image on Remoscope, replicate wells were pooled to obtain at minimum 30uL of 50% hematocrit culture in diluent. For flow cytometry, cells were fixed in 2% paraformaldehyde for 1 hour at room temperature, stained with 50nM YOYO-1 overnight, and then run on a BD FACSCelesta with a collection threshold of 2 million events..

#### Clinical diagnostic testing methods

The clinical cohort described in this work was a subset of the PRISM border cohort study. All thick smear microscopy and qPCR was performed exactly as described in (34).

## Data availability

All data supporting the findings of this study will be made freely available before publication of the final version of this manuscript.

## Code availability

All software developed for this project is open source and publicly available here:

● Instrument control software: https://github.com/czbiohub-sf/ulc-malaria-scope
● YOGO (manuscript in preparation): https://github.com/czbiohub-sf/yogo
● Data analysis utilities: https://github.com/czbiohub-sf/lfm-data-utilities
● Manuscript figure preparation: https://github.com/czbiohub-sf/remo-fig-gen
● Single shot autofocus: https://github.com/czbiohub-sf/ulc-malaria-autofocus

## Acknowledgements

The authors would like to express their gratitude to many members of the Infectious Disease Research Collaboration who helped facilitate the project. We would also like to acknowledge Dr. Georgios Batsios for assisting with lyophilization of the diluent.

## Author contributions

P.M.L led the project; C.C. and P.M.L designed and built the optics and hardware with assembly help from P.C.F, W.W and J.D; E.H designed the custom electronics; M.W.L.K, A.J, and I.J developed instrument control and data analysis software with P.M.L; C.C and P.M.L developed the flow cartridges; P.M.L, C.C, A.J, E.H and P.C.F developed the flow cartridge QC system; W.W fabricated and performed QC on the flow cartridges; image annotation was performed by A.S, P.M.L, and A.J; parasite culture, titrations, life stage experiments, and drug studies were performed by A.S; YOGO was developed and deployed by A.J; J.E collected clinical cohort data and assisted with data pre-processing by G.D; R.G.S and J.D provided guidance and advisory support for the project. P.M.L wrote the paper with assistance from M.W.L.K, I.J, A.J., and A.S.

## Supplementary Information

### Supplementary Videos

Supplementary videos can be downloaded here:

**Supplementary Video 1 | Overview of the Remoscope workflow.** A video demonstrating the workflow from fingertip blood draw, sample loading, metadata entry, cartridge loading, experiment progress, and summary report.

**Supplementary Videos 2-17 | Titration data, points 1-16.** The first 30 seconds of each titration dataset from Figure 3 are shown. The first titration point (highest parasitemia, Supplementary Video 2) was diluted twofold in diluent buffer in order to aid with flowing high concentrations of late-stage parasites.

**Supplementary Video 18 | High clinical parasitemia.** Blood from a participant from the Ugandan cohort with a parasitemia of 654,045 parasites/µL by qPCR. A white blood cell containing phagocytosed hemozoin fragments is visible crossing the field of view.

**Supplementary Video 19 | Sickle Cell Disease.** Blood from a participant from the Ugandan cohort with sickle cell disease is shown. Various RBC morphologies are present including sickled cells, poikilocytes, target cells, among other non-discocytes.

**Supplementary Video 20 | Agglutination.** A video showing *Pf* parasites cultured in red blood cells from one donor, diluted into healthy whole blood from a second, non typed-matched donor. Plasma from the second donor is seen to agglutinate RBCs from the first donor, forming large cross-linked rafts of cells. In conditions such as cold agglutination (31), a similar effect is caused by autoantibodies.

**Supplementary Video 21 | Abnormal RBCs in a Ugandan cohort participant.** Target cells, echinocytes, stomatocytes, and other abnormal RBC morphologies in a participant with normal genotype (not SCD), who also exhibited a parasitemia of 6730.4 parasites/µL (by aPCR).

### Supplementary Figures

**Supplementary Fig. 1.**
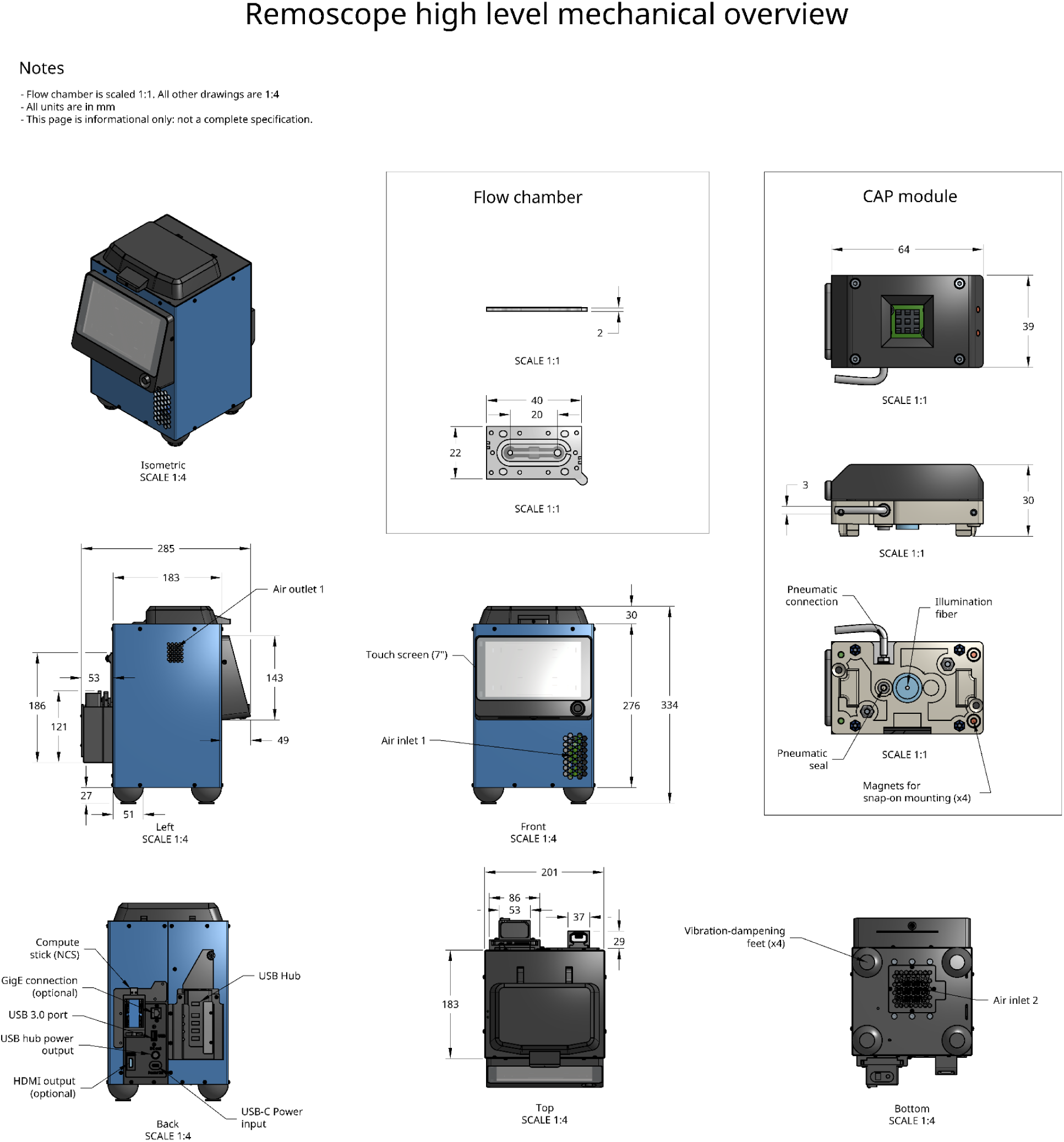
Overall mechanical design of the system, user-facing elements only. High-level overview of the system including overall dimensions of key components including the instrument exterior dimensions, the CAP module, and the flow cartridge.

**Supplementary Fig. 2.**
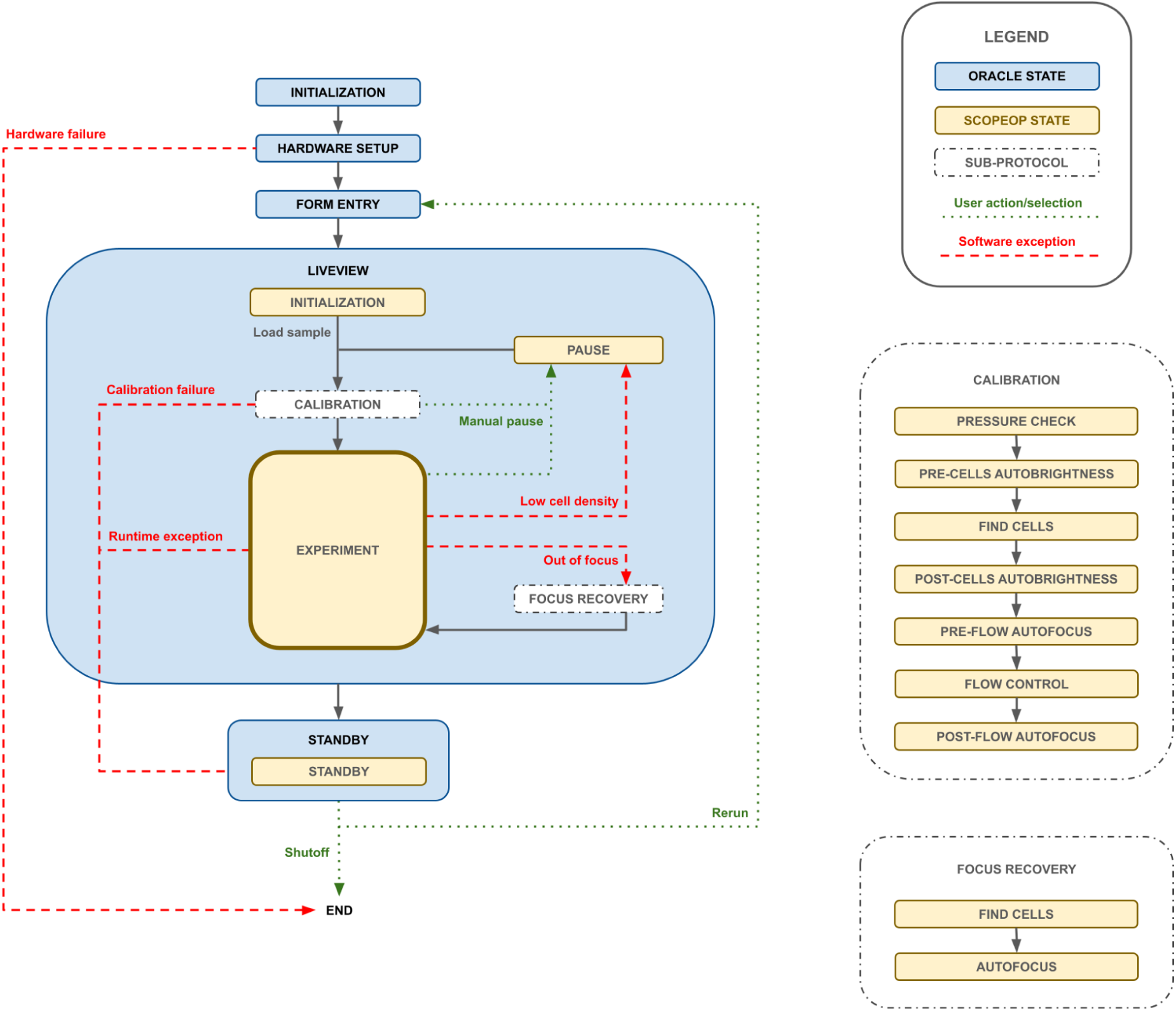
Remoscope software state machine flowchart. Two state machines were used to coordinate the sequence of events in an experiment. “Oracle” is a state machine class used to control the high level flow from standby, setup, form entry, run sample, and run completion states. “ScopeOp” (owned by Oracle), was used to control real-time dynamics of the hardware. All hardware I/O was managed by ScopeOp. In the diagram, Oracle states are denoted by blue boxes with bold text labels, and ScopeOp states are denoted by yellow boxes with regular text labels. Software-triggered state transitions are indicated by red dotted arrows, and user-triggered transitions are indicated by black solid arrows.

**Supplementary Fig. 3.**
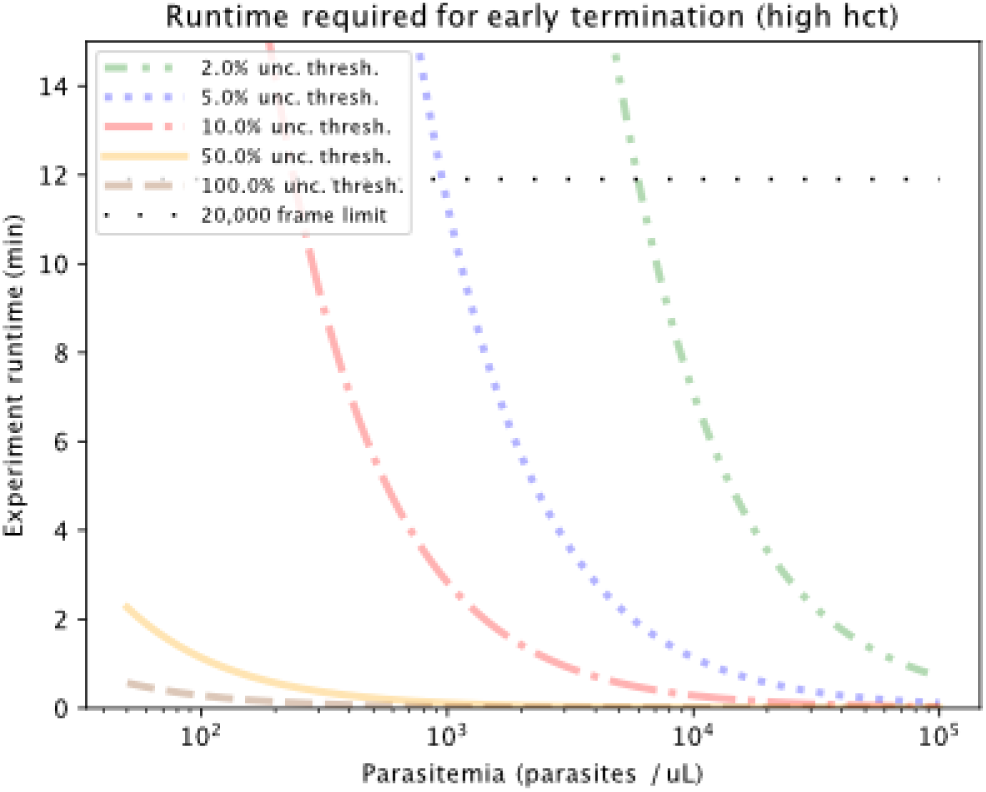
Remoscope early termination estimates for the undiluted blood assay (high hematocrit, (hct)). Based on Poisson counting statistics only, the experiment run duration was computed using the average rate of cellular throughput for various target relative uncertainty thresholds, as a function of parasitemia. These estimates do not include sources of correlated error.

**Supplementary Fig. 4.**
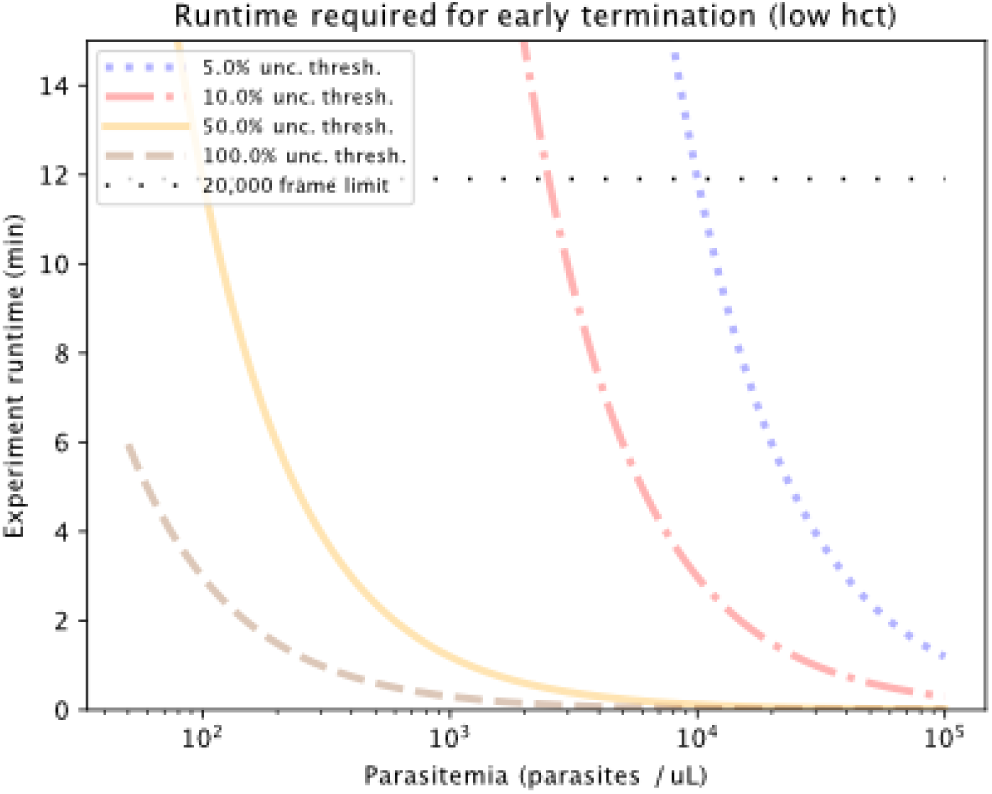
Remoscope early termination estimates for the diluted blood assay (low hct). Based on Poisson counting statistics only, the experiment run duration was computed using the average rate of cellular throughput for various target relative uncertainty thresholds, as a function of parasitemia. These estimates do not include sources of correlated error.

**Supplementary Fig. 5.**
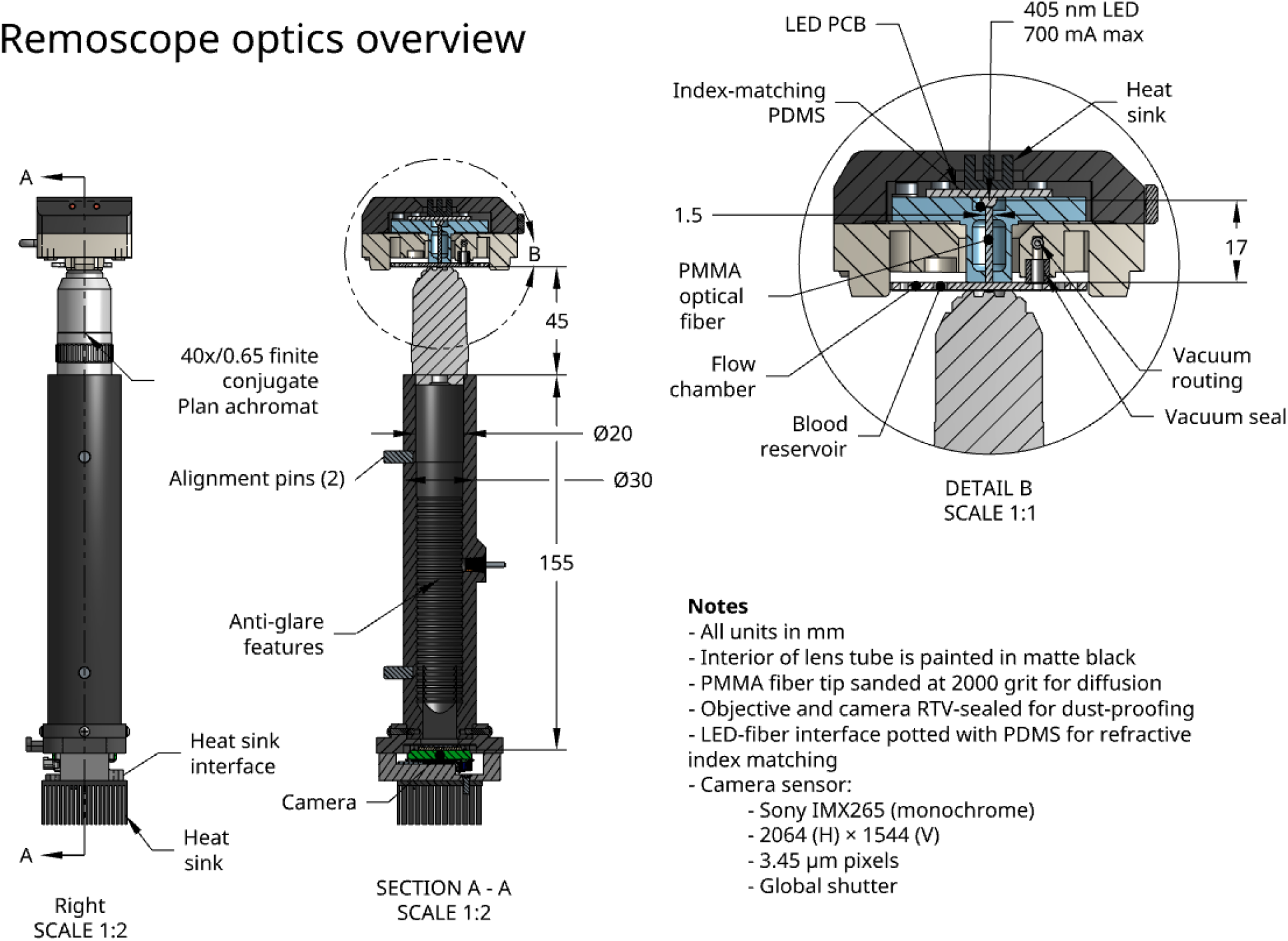
Remoscope optics. Remoscope optics consist of a simple finite conjugate brightfield microscope with 405 nm excitation, provided by a near UV LED that is coupled into a PMMA optical fiber. Polydimethylsiloxane (PDMS) was used as a potting agent to improve coupling into the fiber, providing a 30% increase in efficiency. The Condenser And Pneumatic module (CAP) houses the excitation as well as the vacuum interface to the flow cartridge. Imaging is performed using a Plan Achromat 40× objective with a numerical aperture of 0.65. Images were acquired using an AVT Alvium 1800 U-319 machine vision camera with a monochrome global shutter Sony IMX265 sensor. A silicone RTV compound was used to seal all objective and camera thread interfaces in order to prevent entry of dust into the optics.

**Supplementary Fig. 6.**
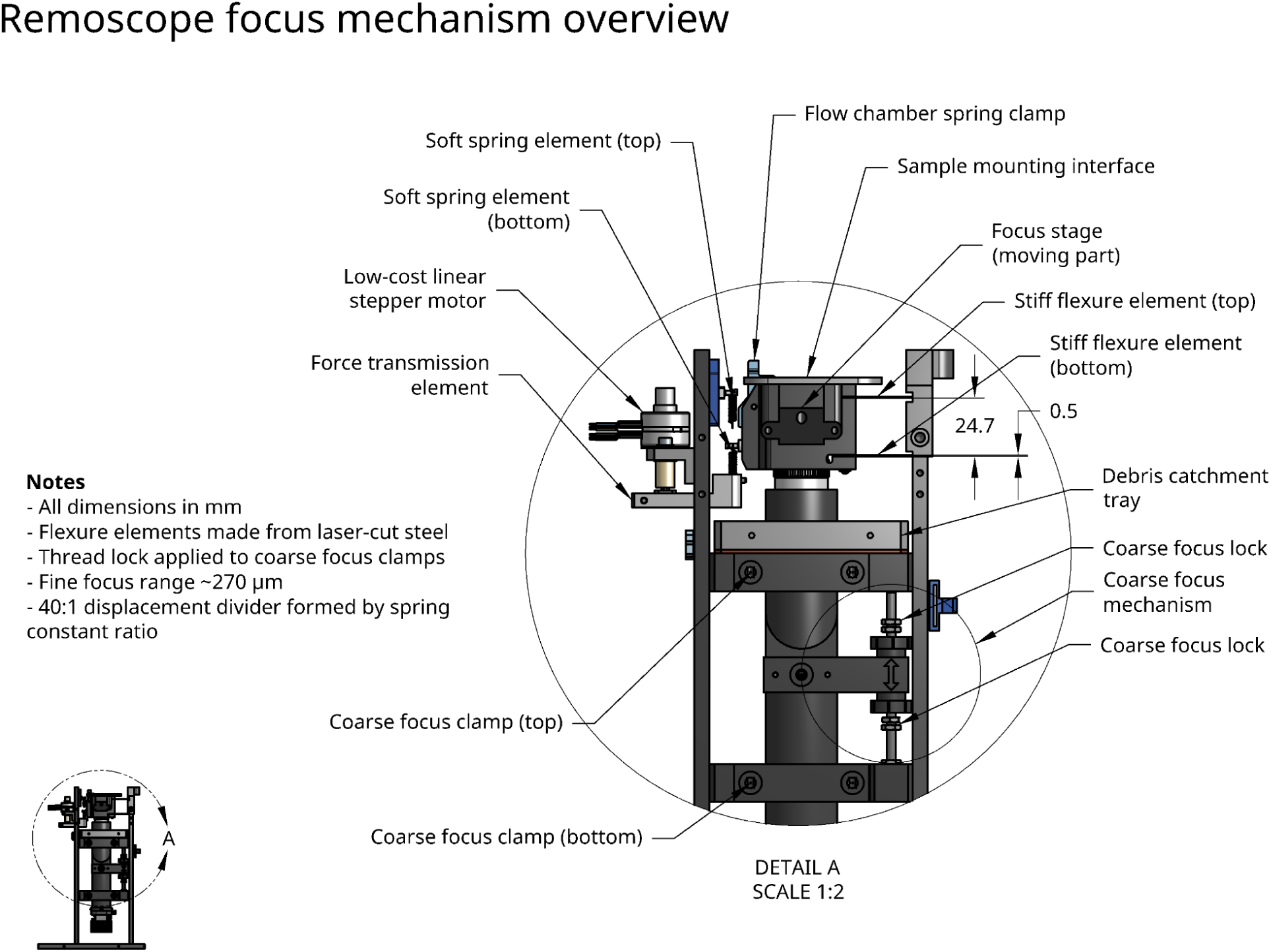
Design of the focus stage. Remoscope has both coarse and fine focus mechanisms. The coarse mechanism moves the optics sub-assembly by rotating translating thumbscrews vertically along a threaded rod, setting and locking the sliding range of motion. Adjustments on the order of ∼5-10 microns can be made via careful rotation of the lower thumb screw, allowing coarse finding of the sample focus during assembly of the instrument (not for routine use). The fine focus mechanism consists of a 40:1 “displacement divider” analogous to a voltage divider: the ratio of spring constants (soft/stiff elements) defines a relative motion amplitude of the sample stage with respect to the linear stepper motor. The total displacement range is ∼270 µm with a step size of ∼0.3 µm.

**Supplementary Fig. 7.**
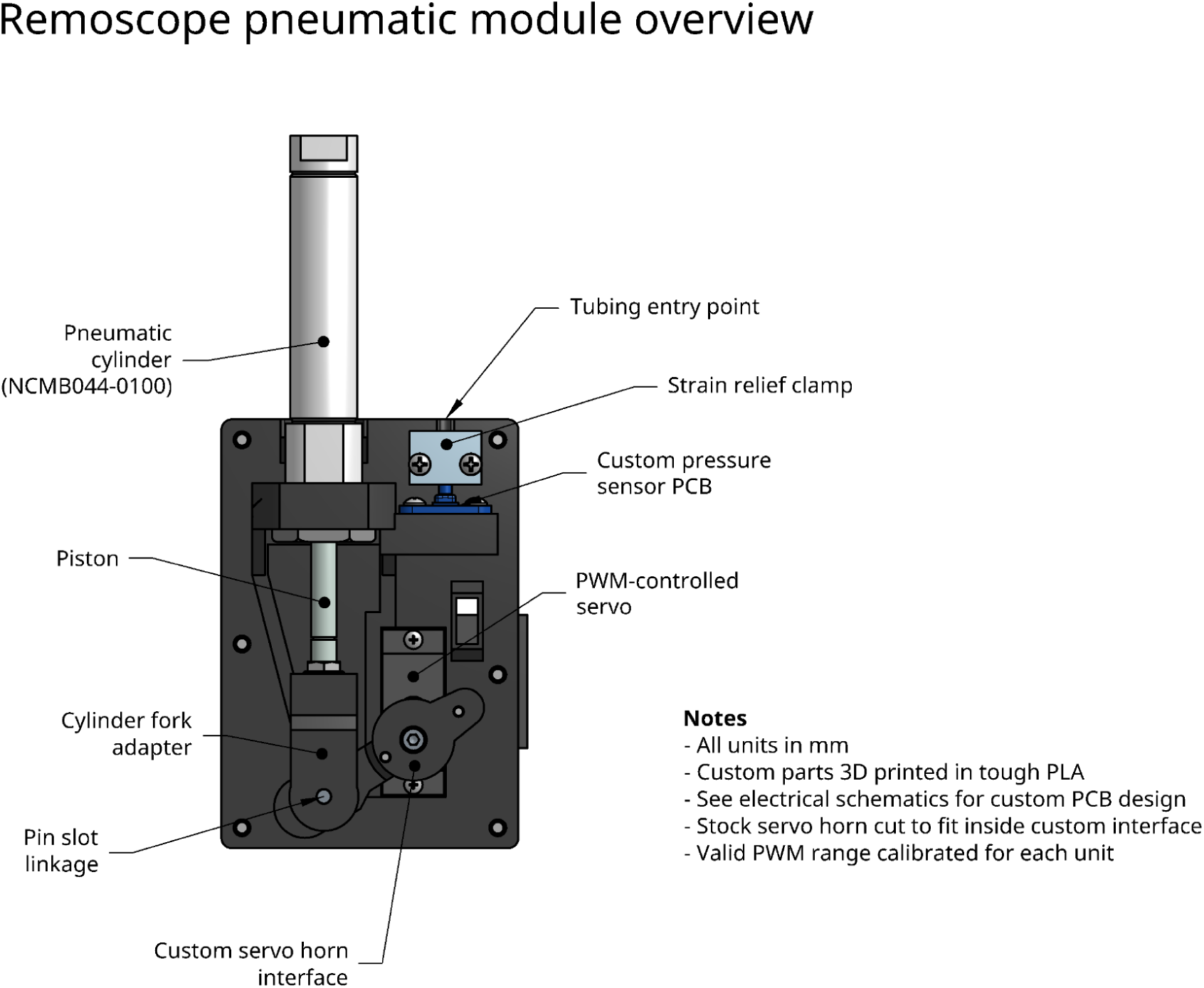
Design of the pneumatic module. The pneumatic module uses a PWM-controlled servo motor to adjust the piston of a commercial pneumatic cylinder, via a pin slot linkage formed by a 3D-printed interface to the servo horn and a pin mounted to the cylinder fork adapter. Polyethylene tubing is connected to the top of the cylinder, with a tee junction to split off the connection to the pressure sensor. The fluted, absolute pressure sensor is mounted on a custom PCB. Each module has a valid PWM range that is calibrated on-scope using an automated script in the ulc_mm_scope software package.

**Supplementary Fig. 8.**
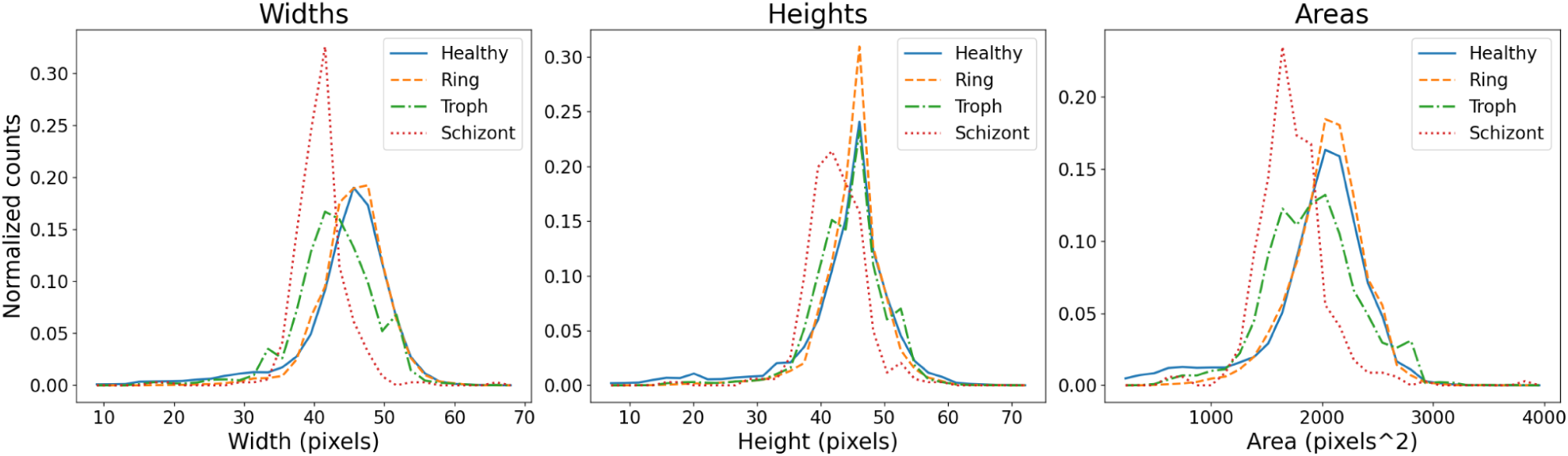
Cell size histograms. Histograms of bounding box dimensions of fully-annotated cells by class type from the training data, containing both cultured and clinical samples. Each histogram is normalized to the total count for its cell type.

**Supplementary Fig. 9.**
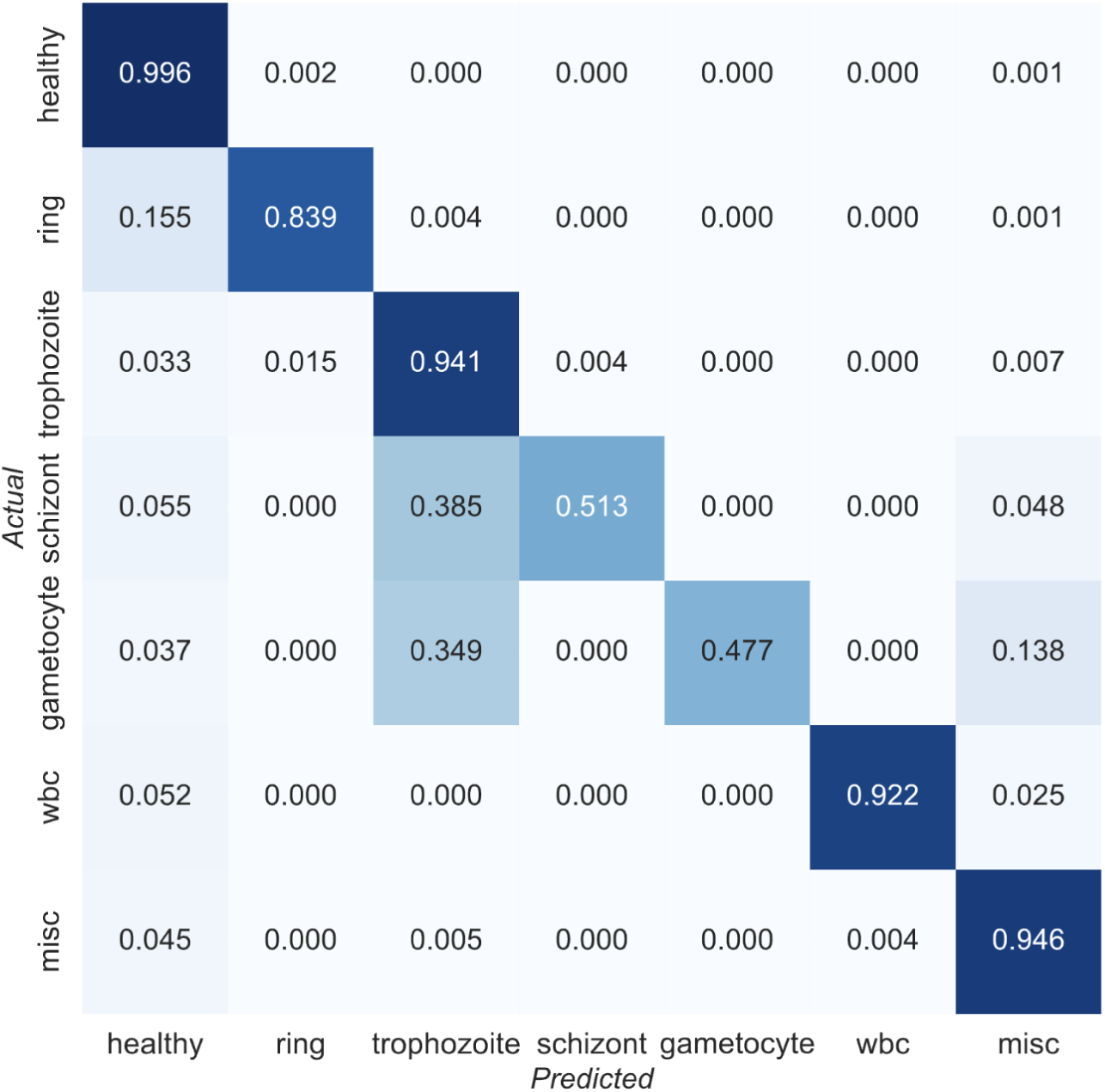
YOGO confusion matrix. The row-normalized confusion matrix was generated by comparing YOGO predictions to human-annotated Remoscope images on a test partition of the data (not seen during training). ‘Actual’ denotes human-annotated labels and ‘Predicted’ denotes YOGO model predictions.

**Supplementary Fig. 10.**
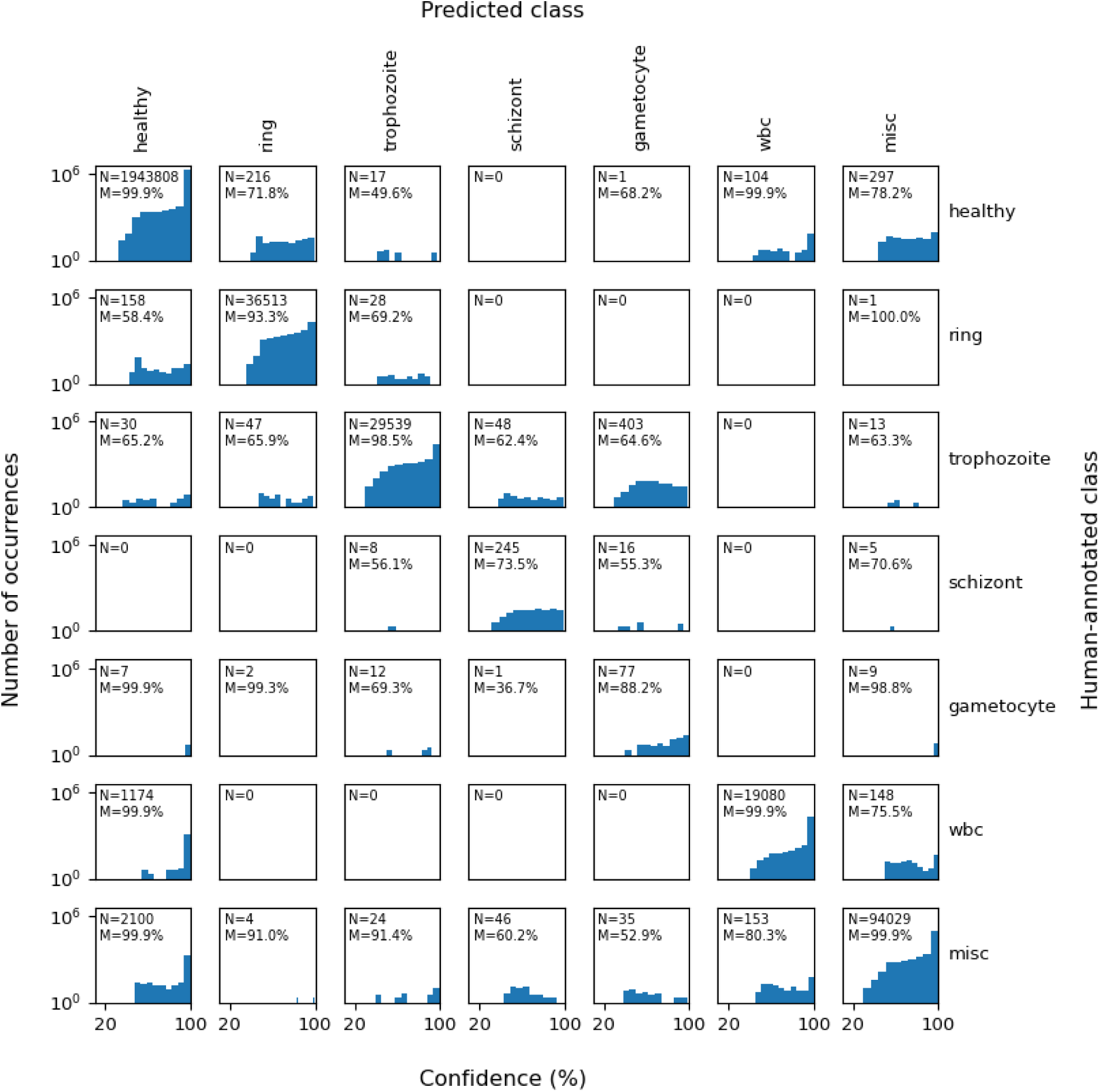
Matrix of YOGO confidence scores. Each sub-panel is a histogram of model confidence scores for a subset of test data with the given ‘Actual class’ (human annotated label) and ‘Predicted class’ (model predicted label), with one-to-one correspondence with the confusion matrix.

**Supplementary Fig. 11.**
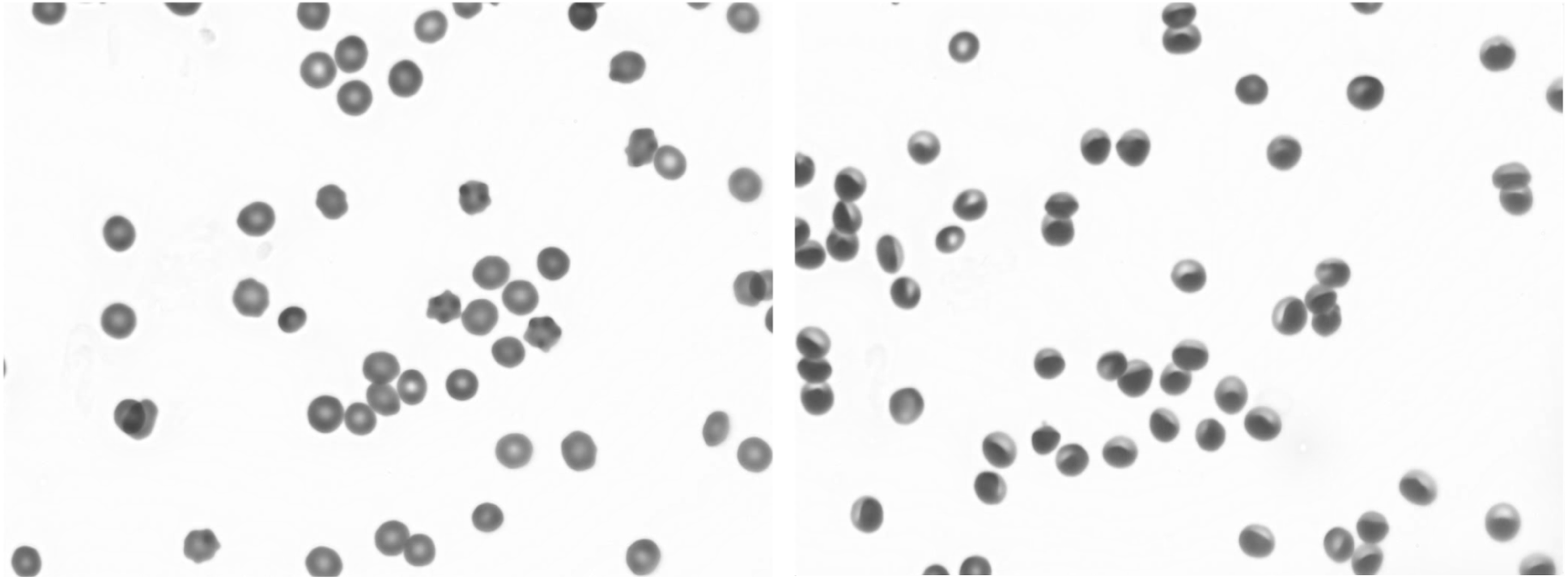
Effect of flow rate on RBC shape. Diluted blood from a Ugandan cohort participant shown in both stopped (left) and under moderate flow (right). In the stopped condition, RBCs are seen to be primarily discocytes with some type I echinocytes. Under flow, the shear forces cause the cells’ leading edges to enrich and their trailing edges to rarify.

**Supplementary Fig. 12.**
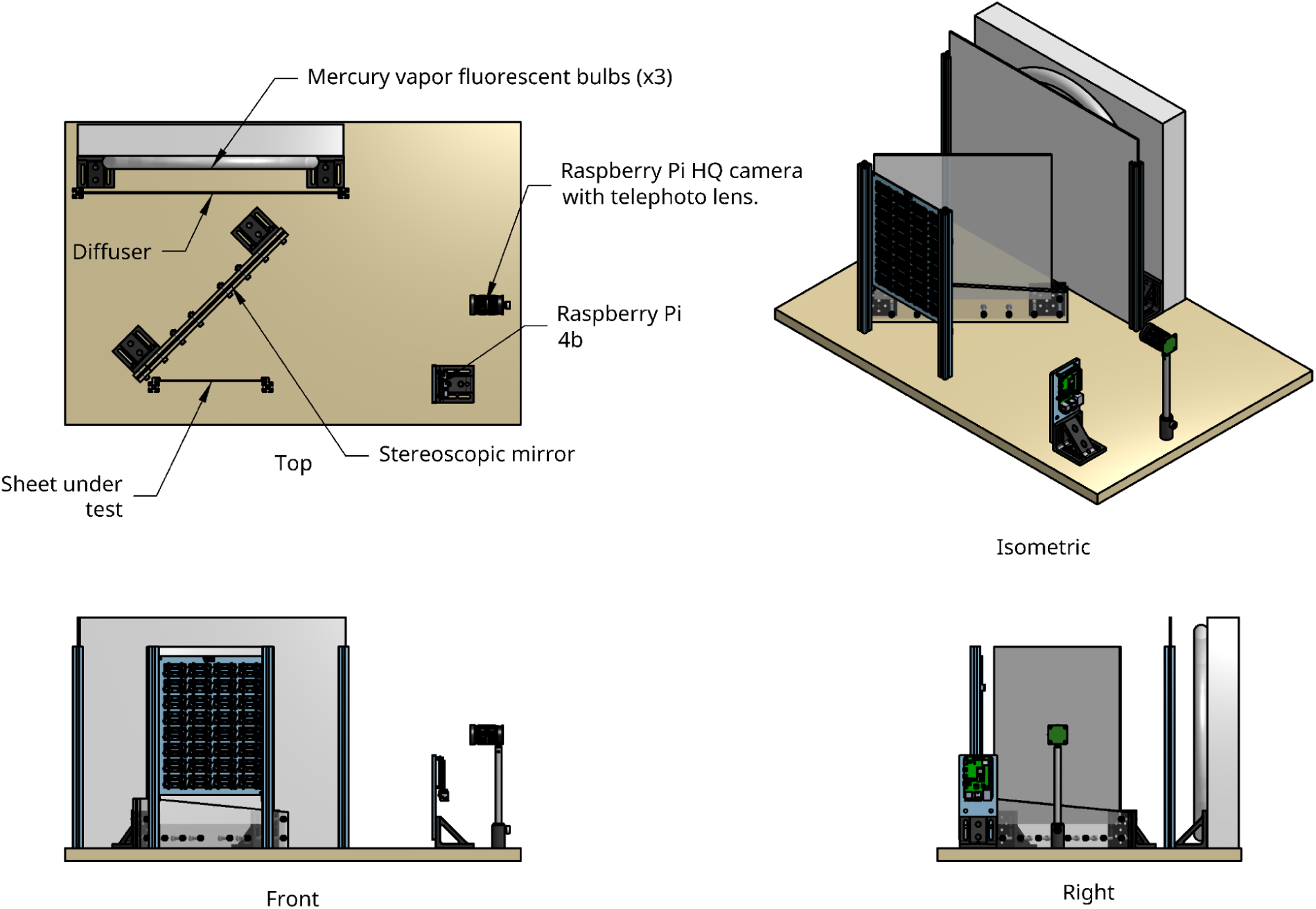
Design of the flow cartridge quality control interference fringe imager. The imager was constructed by focusing a Raspberry Pi HQ camera with telephoto lens option onto the reflected image of a flow cartridge sheet. The central mirror (purchased from www.stereoscopicmirror.com) was a 14 x 14 inch square, partially-silvered mirror used to transmit fluorescent bulb illumination onto the sheet under test, which was then imaged via reflection onto the camera. The setup permitted uniform illumination of the sheet by three concentric mercury vapor bulbs, which exhibit peaked spectra conducive to forming interference fringes by virtue of the etalon effect within the thin flow cartridge. Interference fringe patterns were used to assess the flatness of the flow layer, the approximate thickness of the flow layer (presence of strong fringes indicated the chamber was appropriately thin), the wicking of the glue used for bonding, or leaking of glue into the flow channel. Results were assessed in a semi-manual fashion using custom software allowing an operator to score flow cartridges’ defects and automatically entering the results into a database.

**Supplementary Fig. 13.**
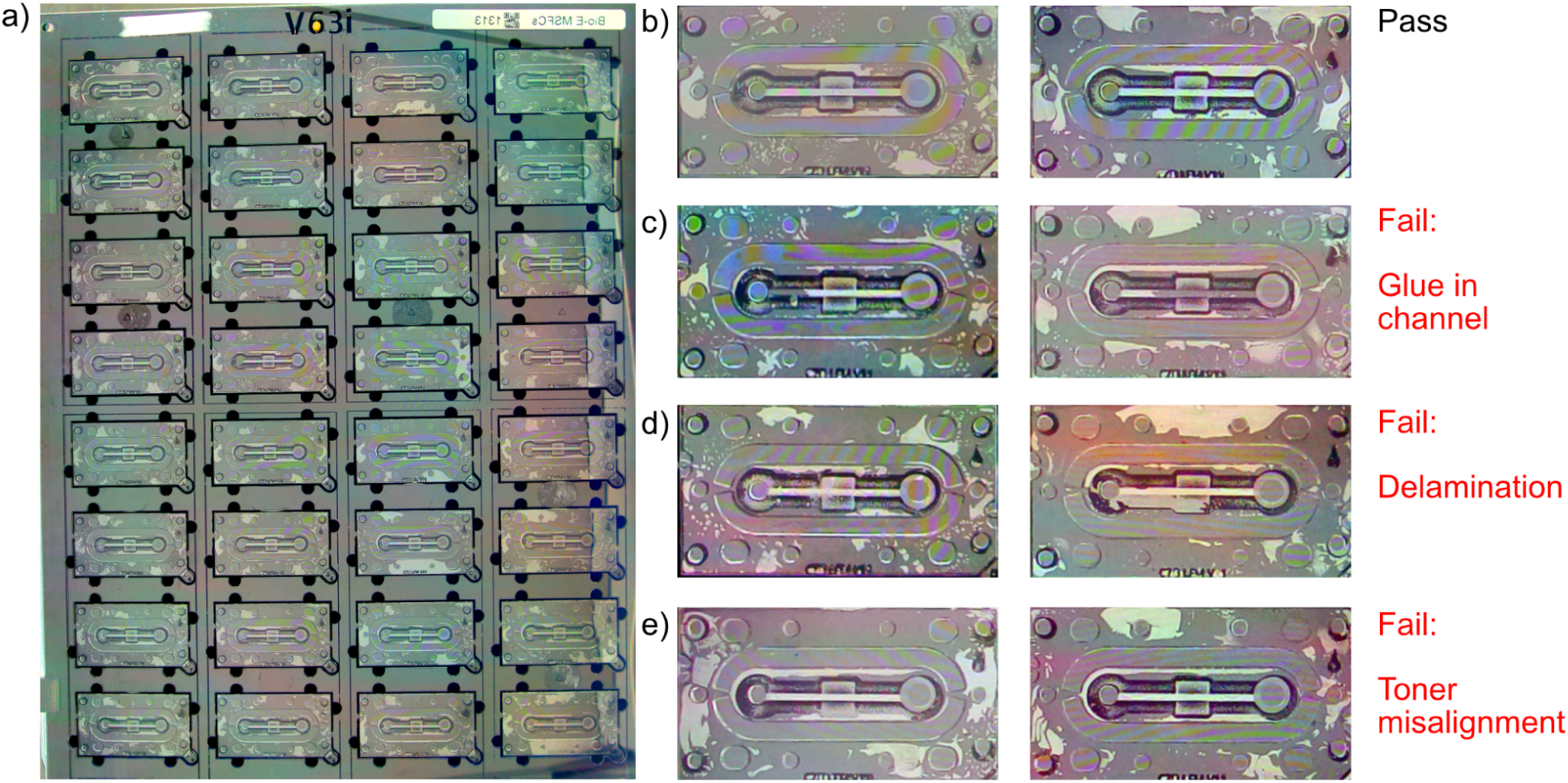
Example images from the interference fringe imager. a) Whole sheet image from the interference fringe imager. b) Example flow cartridges passing quality control. c) Example flow cartridges failing by presence of glue in the channel. d) Example flow cartridges failing by delamination. e) Example flow cartridges failing by misalignment of the toner pattern.

**Supplementary Fig. 14.**
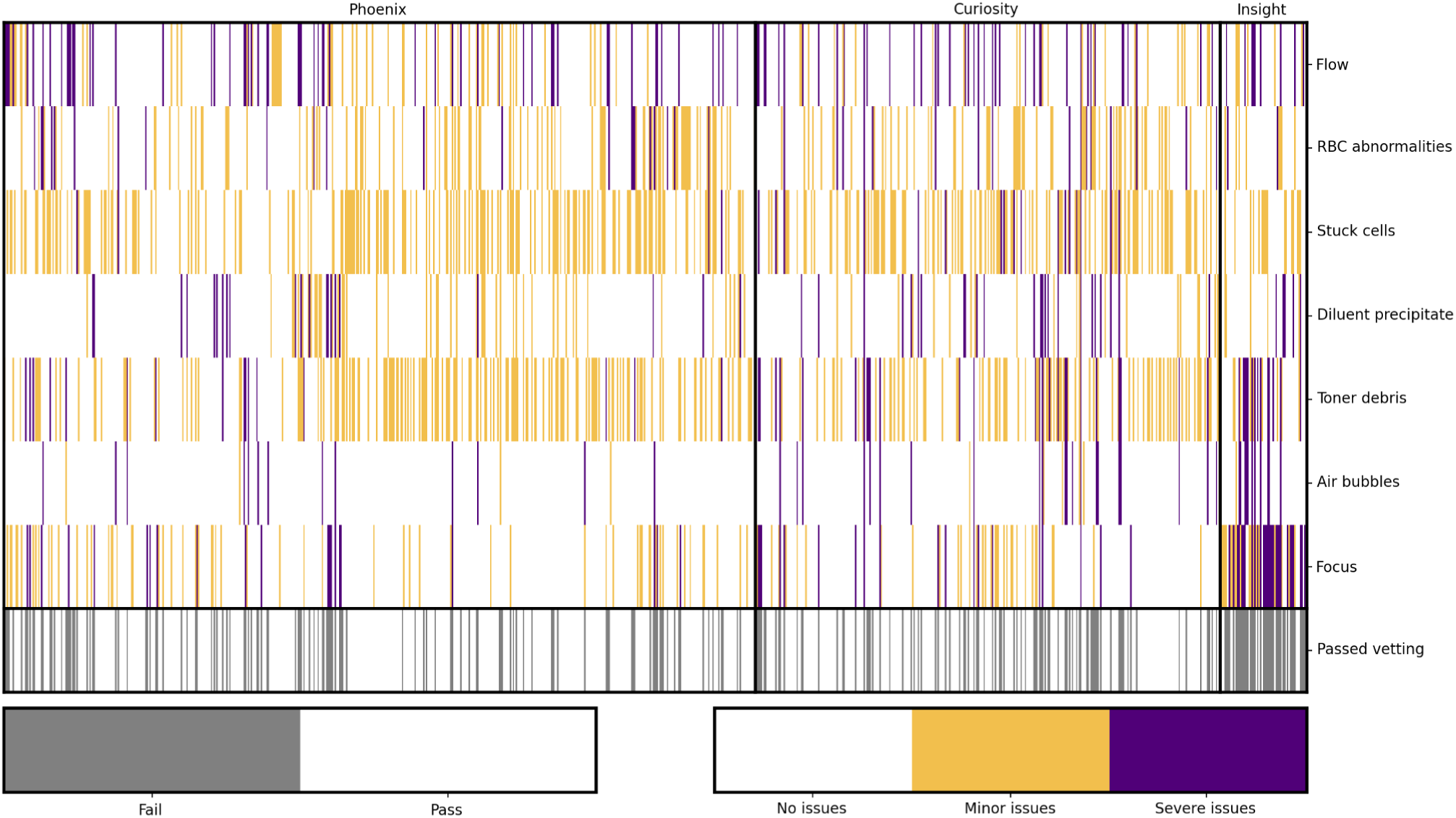
Manual vetting criteria for quality control of clinical runs. Videos exported from individual experiments in the PRISM border cohort were viewed and scored by the following criteria. Each criteria was subjected to a score of ‘No issues’, ‘Moderate issues’, or ‘Severe issues’. An experiment was rejected if it suffered from one or more severe issues, or if it suffered from four or more moderate issues:

- Flow: Stability of the flow rate.
- Condition of the RBCs: if the majority of RBCs were damaged/lysed the sample was rejected from analysis.
- Stuck cells: if more than ⅓ of the video had ⅓ of the field of view obscured it was considered ‘moderate’. If more than half the field of view was obscured for at least half the video it was considered ‘severe’.
- Diluent precipitate: If reconstituted diluent formed a precipitate that was visible and blocking ⅓ of the field of view it was considered moderate. More than ½ the field of view was considered severe.
- Toner debris: if stray droplets of printer toner (ink) covered the field of view, it was considered either moderate or severe, subject to the reviewer’s discretion.
- Air bubbles: if air bubbles obstructed the field of view, it was considered either moderate or severe, subject to the reviewer’s discretion.
- Focus: Single Shot Auto-Focus (SSAF) functioned the majority of the time, but some videos were degraded by poor focus. Reviewer’s discretion was used to score severity.

**Supplementary Fig. 15.**
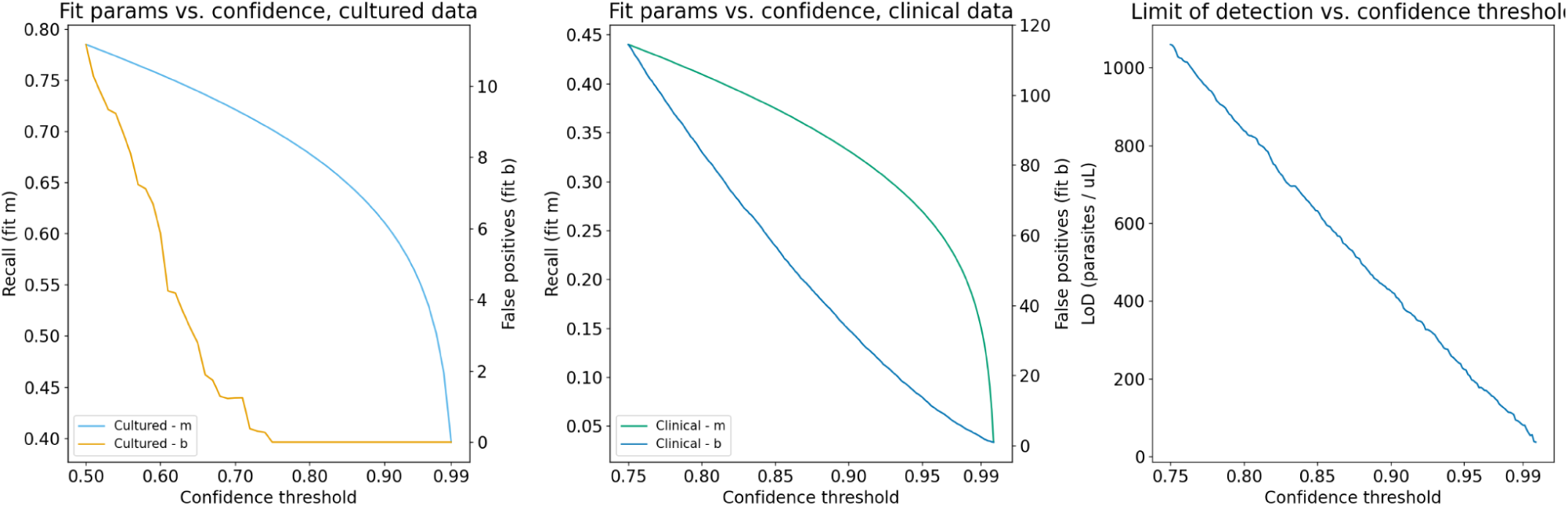
Transformation parameters and limit of detection as a function of model confidence. Linear transformation of the data was performed using fit parameters that were confidence threshold-dependent. With increasing confidence threshold, recall (m) and false positive rate (b) for overall parasitemia both decrease.

**Supplementary Fig. 16.**
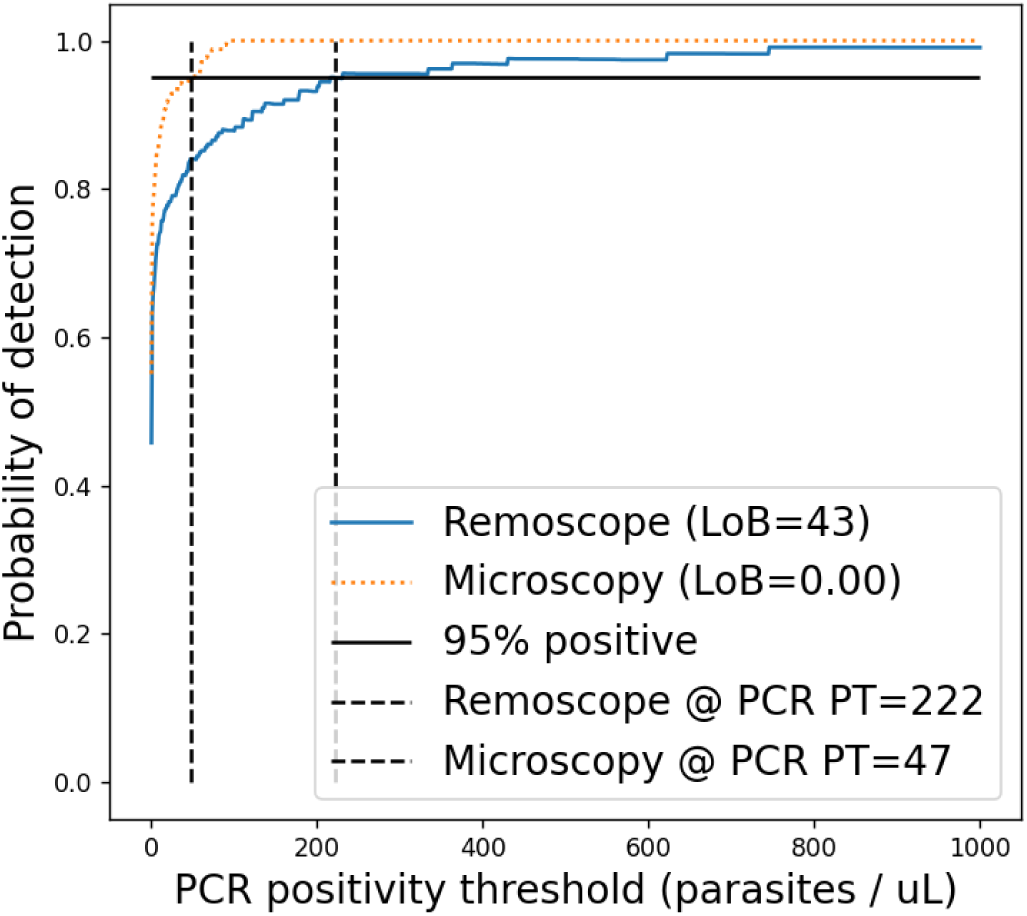
Probability of detection by Remoscope and microscopy. For each method, the fraction of points scoring above the Limit of Blank (LoB) as a function of parasitemia level. The point at which each method is able to detect 95% of points as positive is highlighted. The LoB for both methods was computed as the mean plus 1.645 standard deviations of all qPCR negative samples.

**Supplementary Fig. 17.**
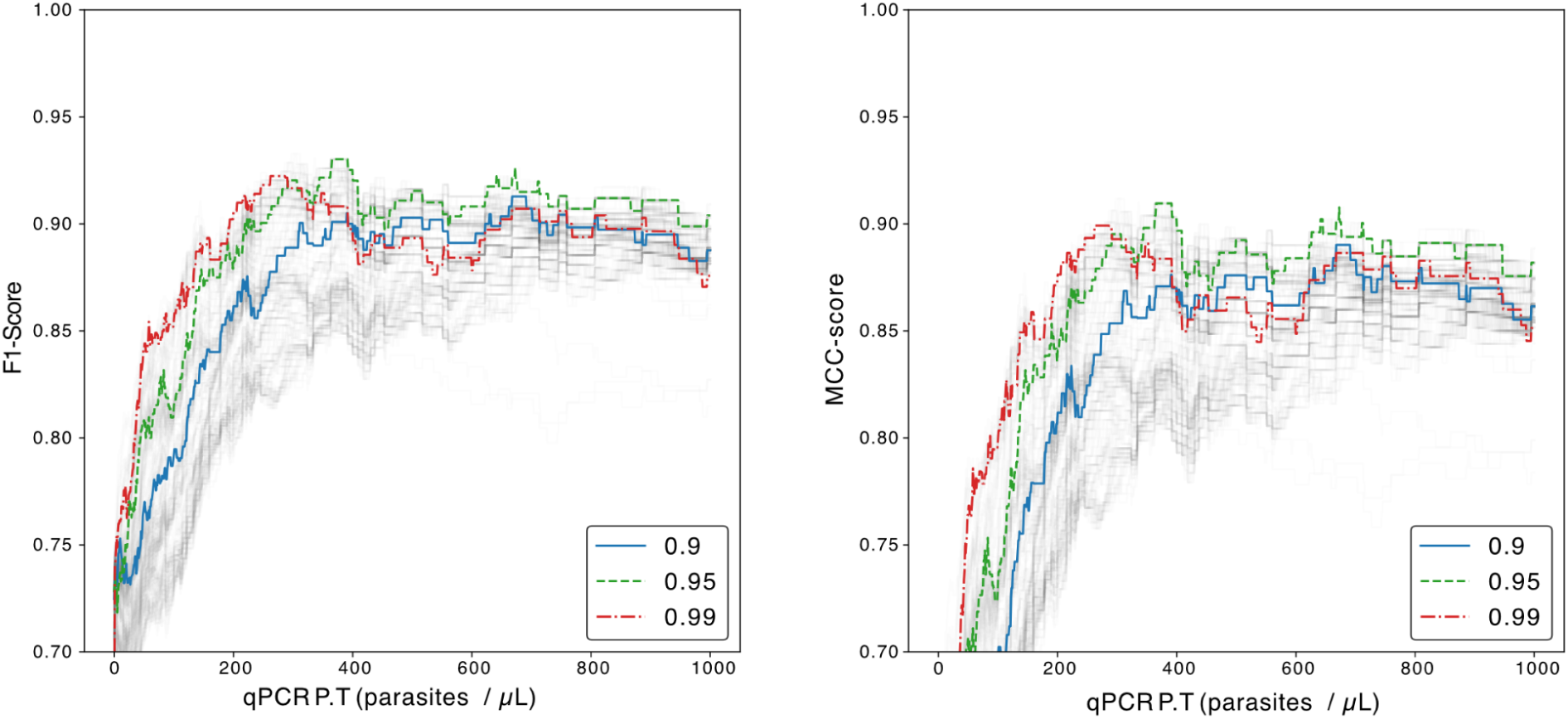
Remoscope F1 and MCC scores from the clinical cohort diluted blood assay presented as a line chart. Line plots are shown for confidence thresholds between 0.5 and 0.99, with key values highlighted.

**Supplementary Fig. 18.**
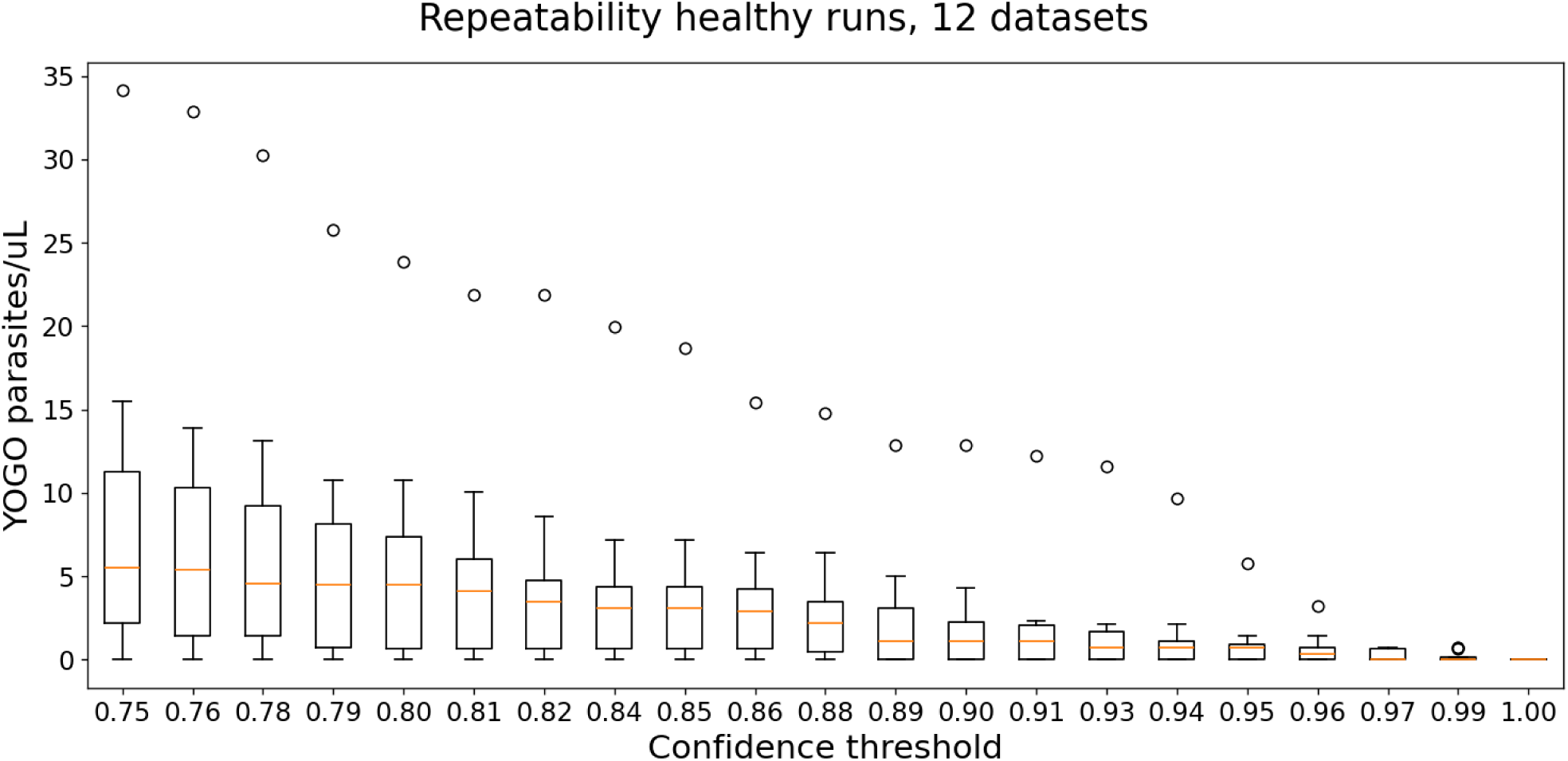
Box plot analysis of false positives as a function of confidence threshold for N=12 experiments from non-endemic donors, using the undiluted blood assay. The box widths represent the interquartile range (IQR), the yellow bands represent the median of the distribution, whiskers extend to the farthest datapoint lying within 1.5 times the IQR, and the individual data markers represent points outside the whiskers.

**Supplementary Fig. 19.**
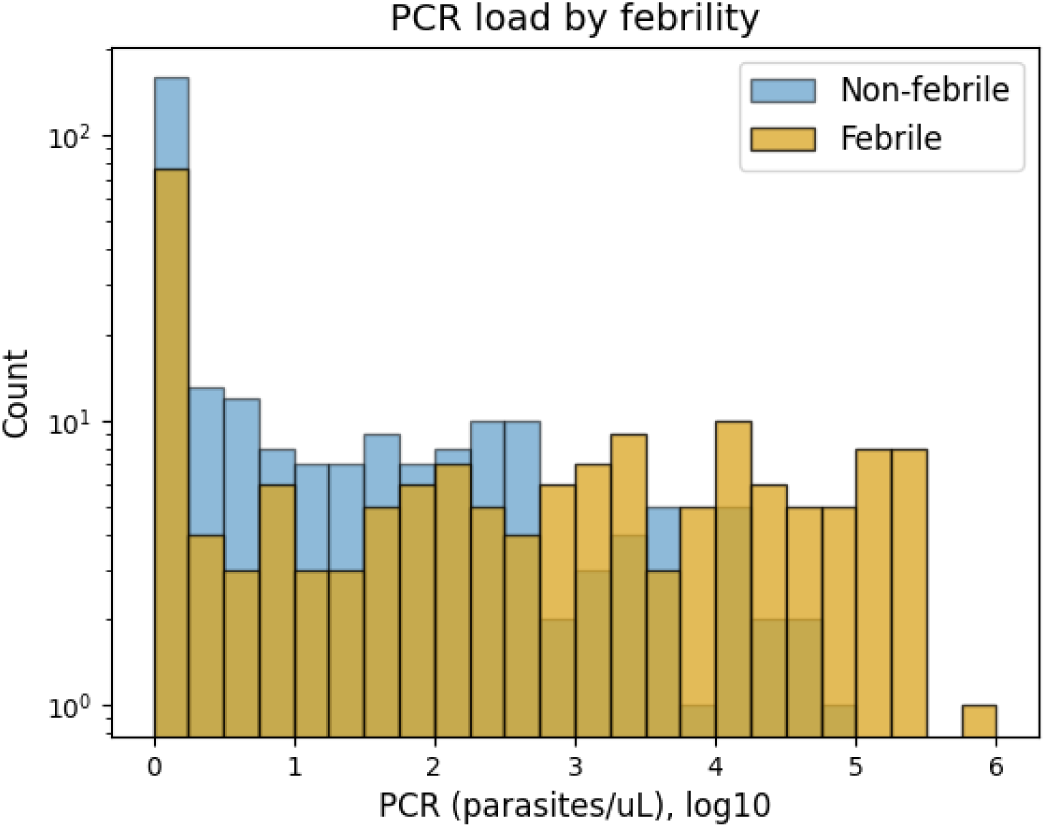
Histogram of clinical parasitemias by PCR stratified by febrility status. The correlation with febrility increased with high parasitemia levels above (> 1,000 / µL).

**Supplementary Fig. 20.**
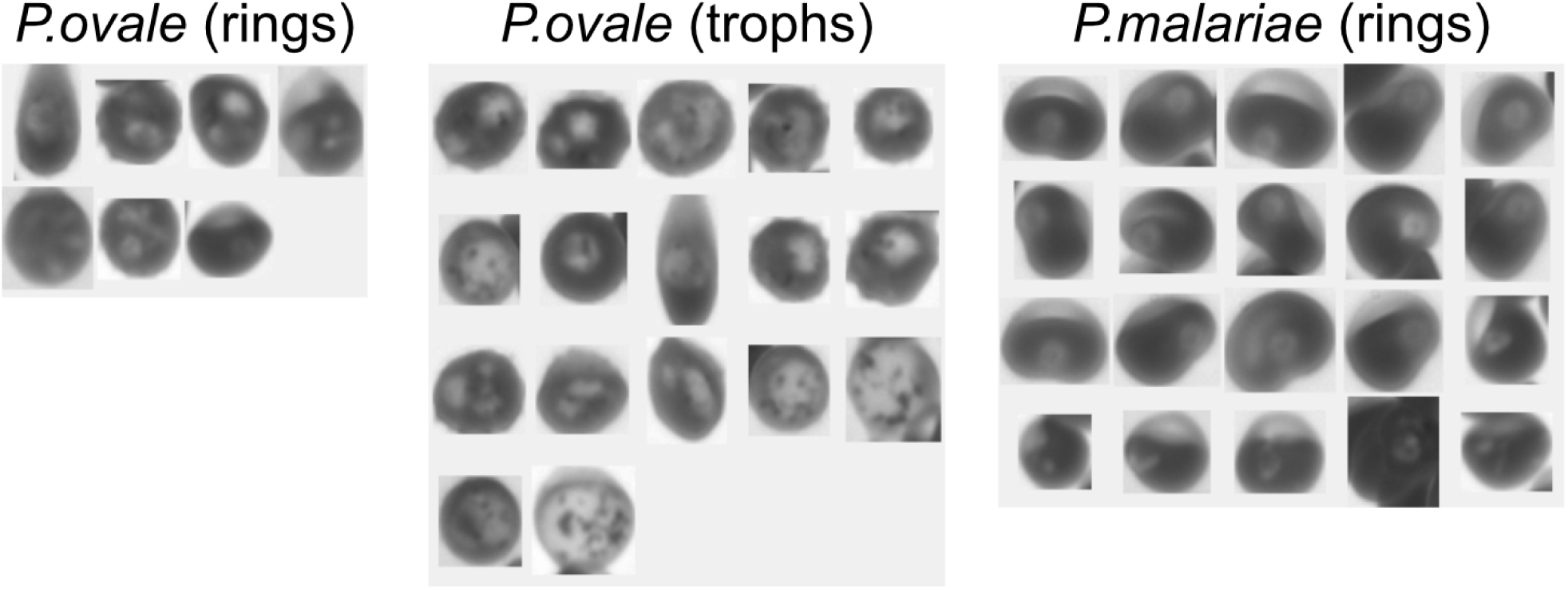
Putative *Plasmodium ovale* and *Plasmodium malariae*, found by YOGO in the clinical cohort. The indicated species was determined by a non-*falciparum* PCR panel in participants who tested positive by microscopy but negative by *Pf* qPCR reaction. YOGO was not trained to detect these species of parasites.

**Supplementary Fig. 21.**
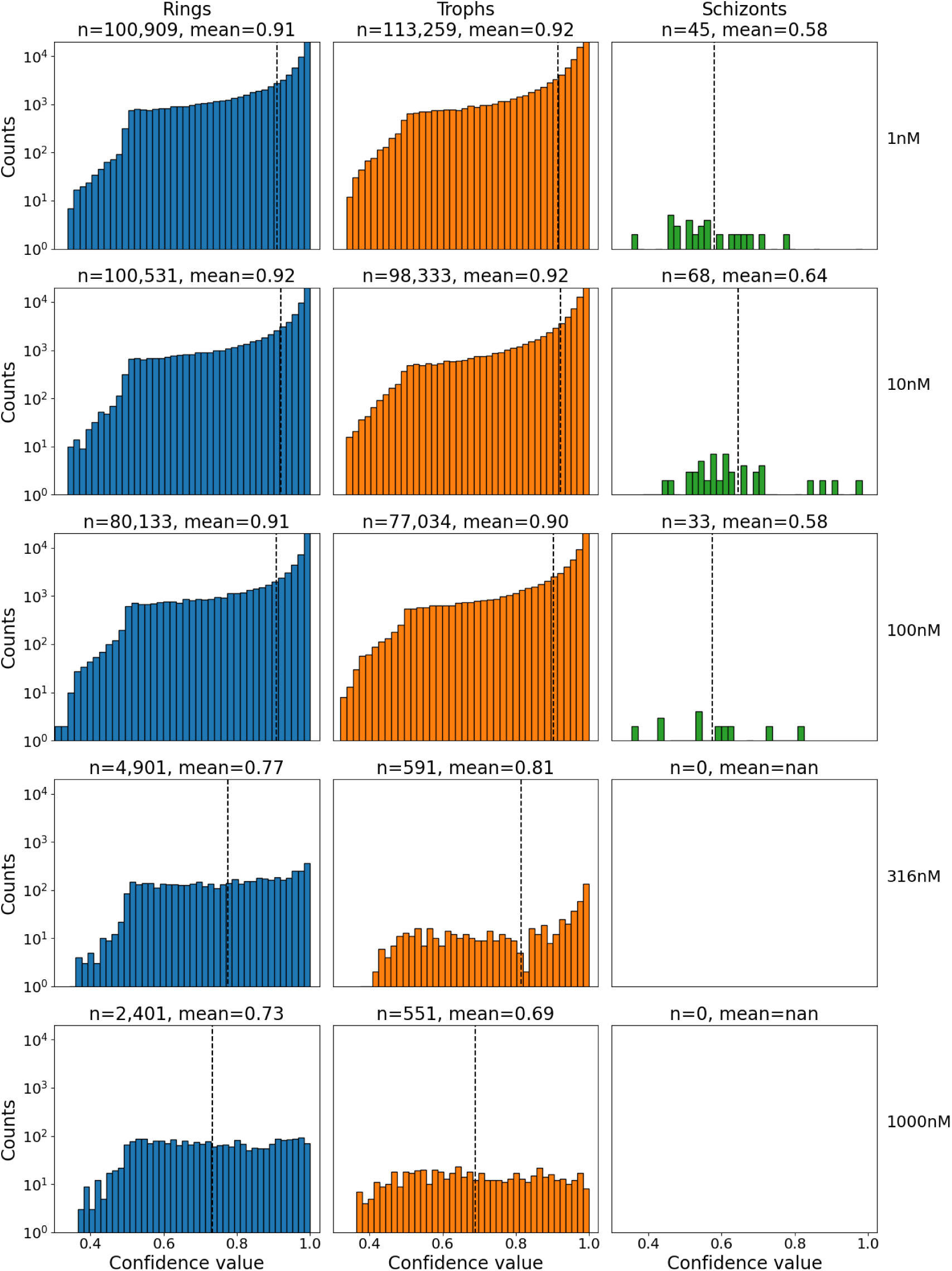
Model confidence vs drug concentration. Histograms showing YOGO confidence distributions as a function of CQ concentration, for *Pf* rings, trophozoites, and schizonts. YOGO was not trained on drugged parasites, and shows a decrease in mean confidence score in addition to a reduction in total number of detected parasites, consistent with drug-induced parasite morphological changes and death.

**Supplementary Fig. 22.**
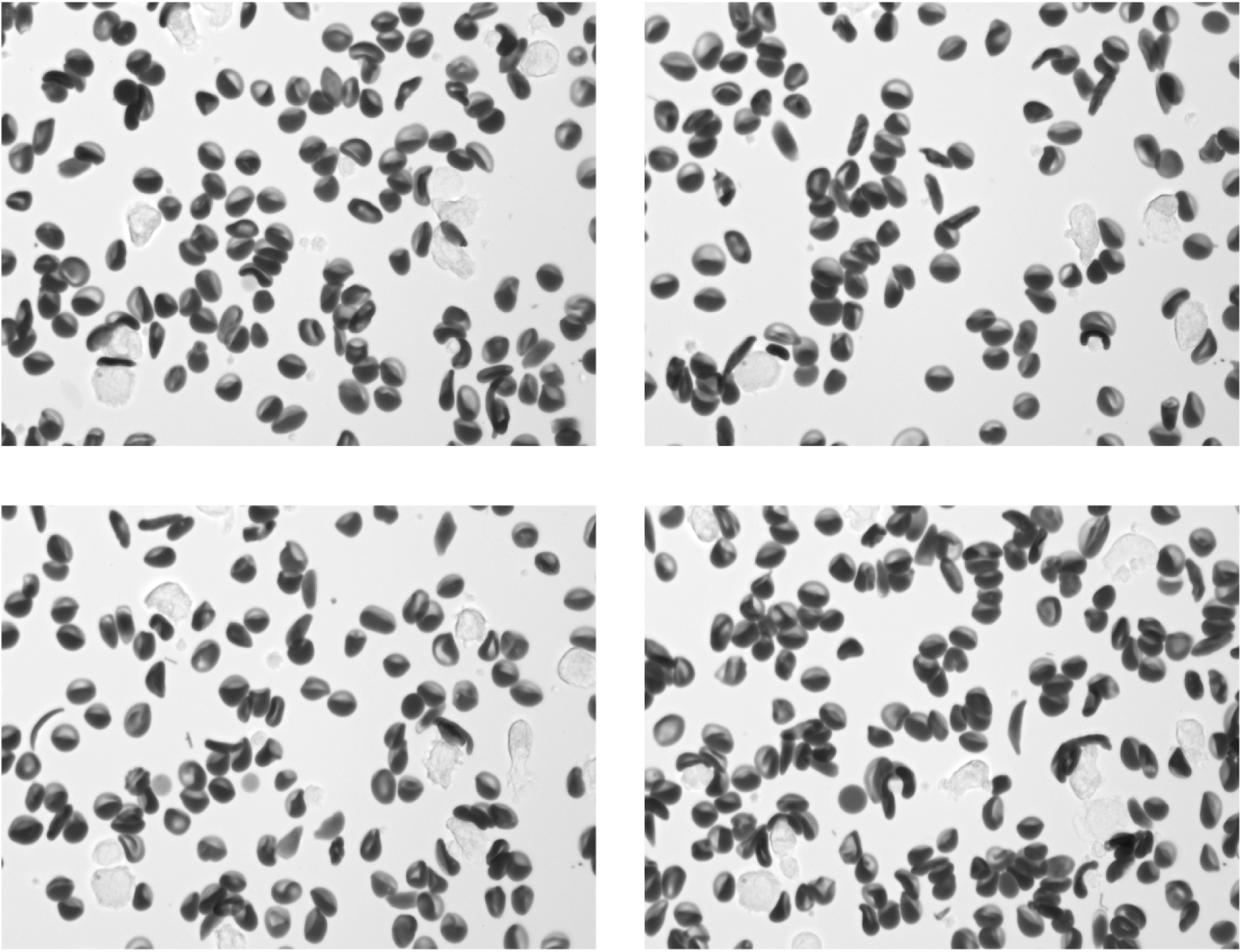
Example images from a participant with Sickle Cell Disease (SCD). While some cells exhibit well-known sickling behavior, the majority of RBCs exhibit a diversity of abnormal morphologies.

**Supplementary Fig. 23.**
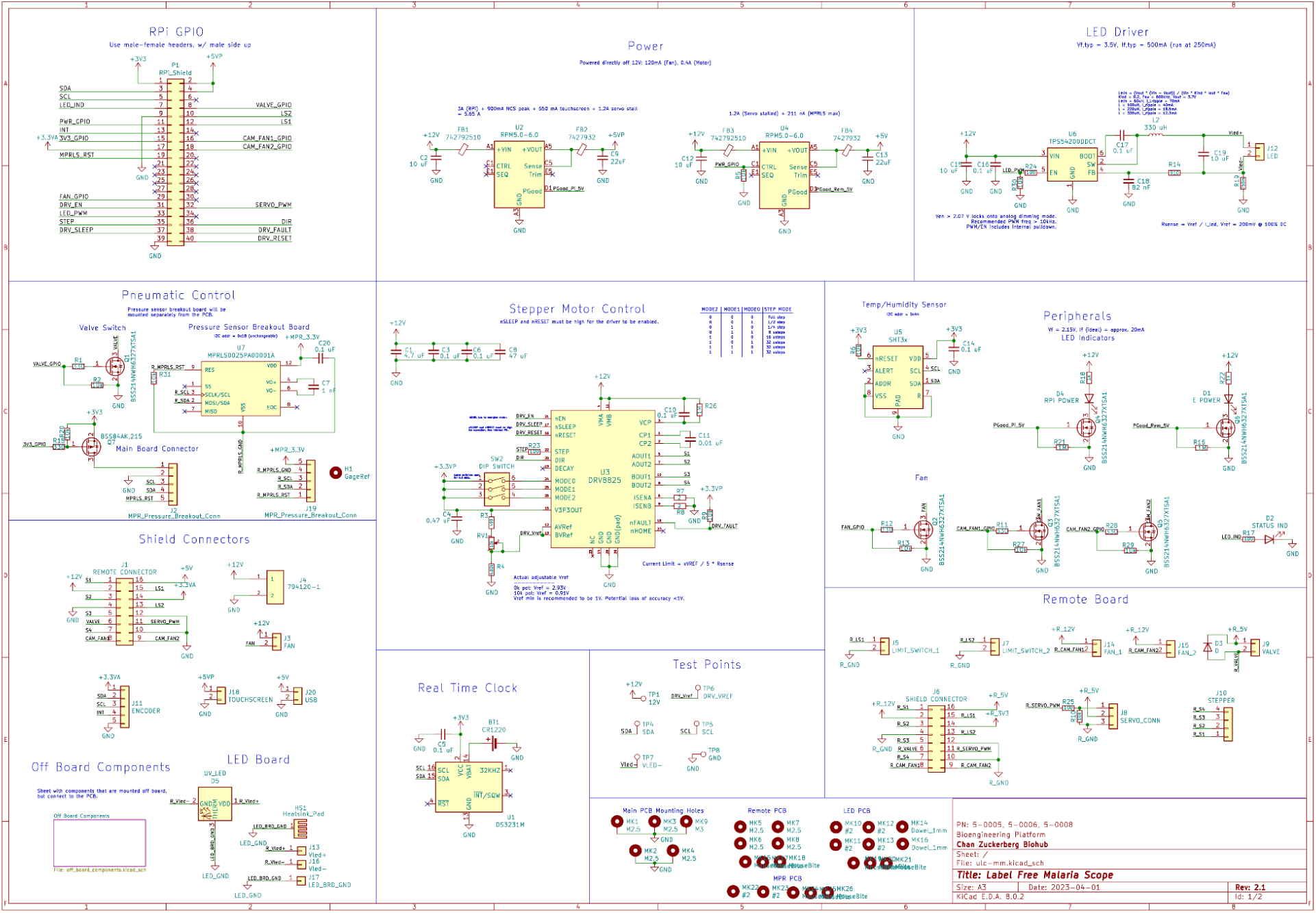
Remoscope main and daughter board PCB design schematic. This diagram contains all components present on the main board, daughter board, LED board, and pressure sensor board. See https://github.com/czbiohub-sf/remo-pcbs for full design details including PCB layout.

**Supplementary Fig. 24.**
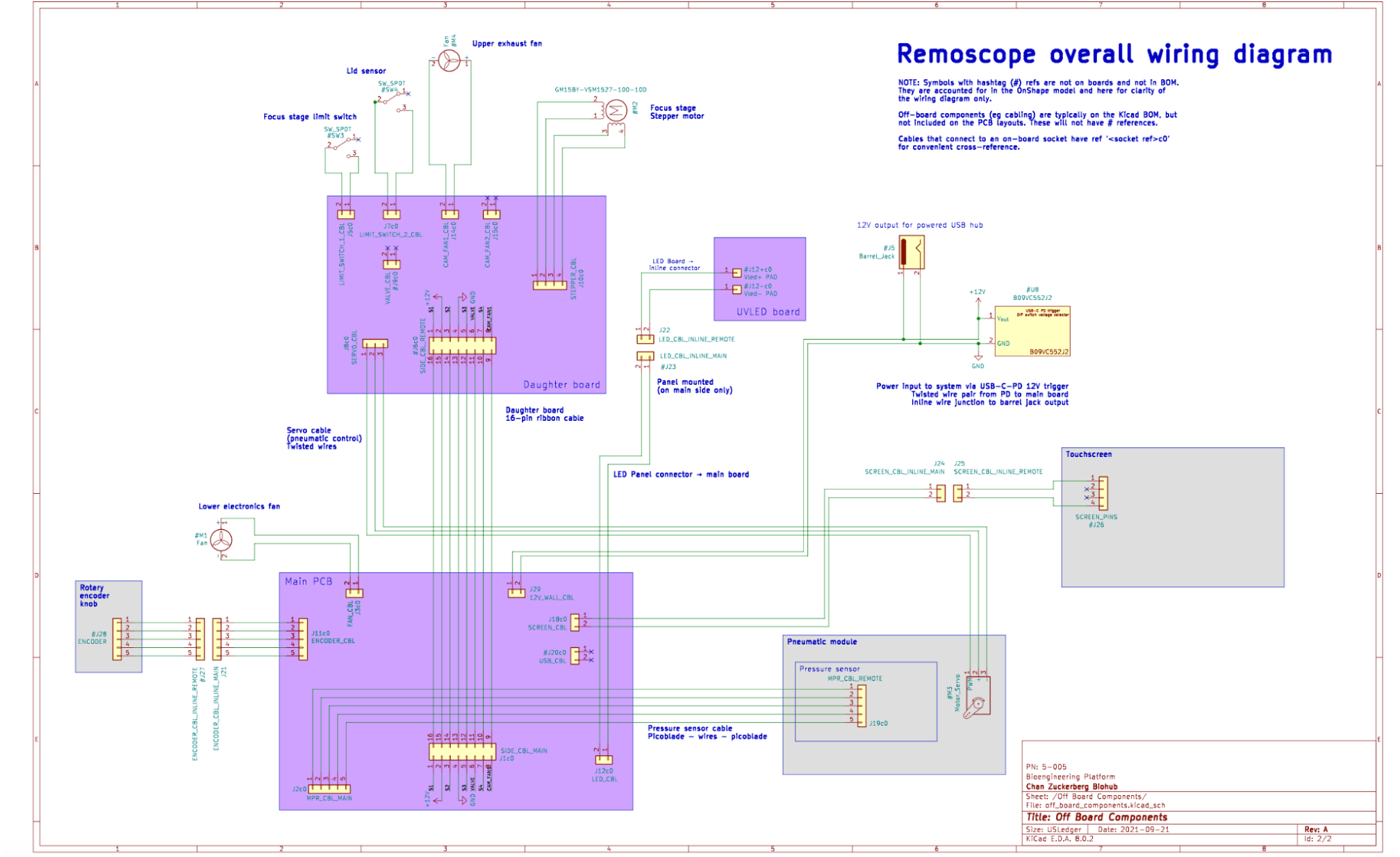
Remoscope overall wiring diagram. Shaded purple boxes represent custom printed circuit boards, and shaded beige boxes represent other modules within the system, and groups of wires represent custom cabling solutions.

