## Supplementary Note 1 for "Remoscope: a label-free imaging cytometer for malaria diagnostics"

### Supplementary note 1:
### Remoscope annotation guide

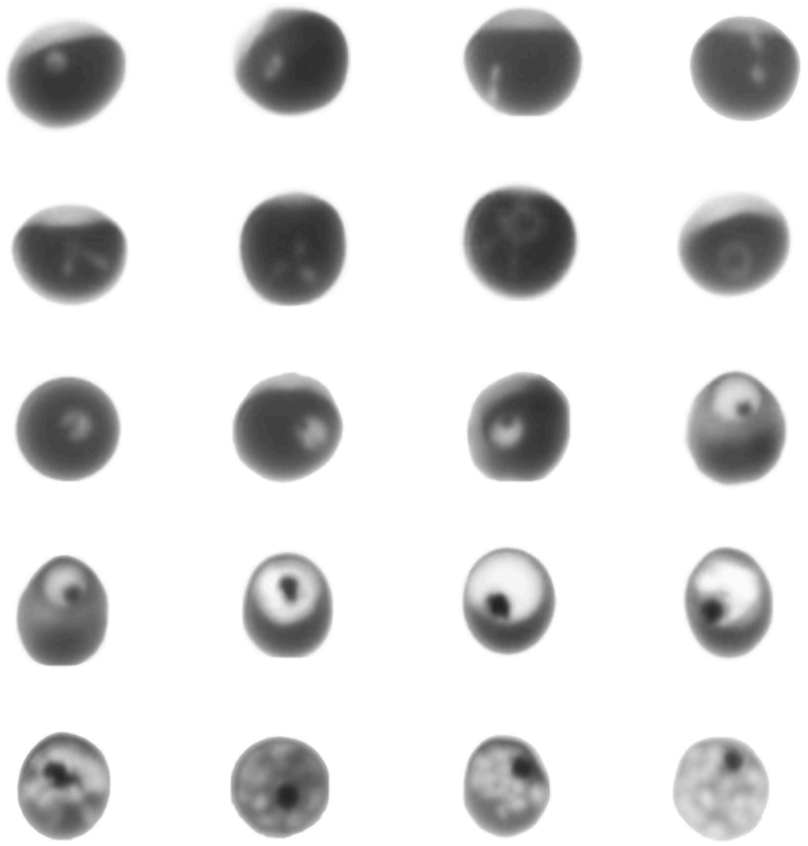

### Table of Contents


**Table of Contents** 2
**Annotation philosophy** 3
**Rings** 3
- Early Rings 3
  - Dendritic Rings 3
  - Amoeboid rings 4
- Canonical Rings 4
  - “Slit” rings 4
  - Multiple ring invasions 4

**Trophozoites** 5
- Early trophozoites 5
- Mature Trophozoites 5
    - Multiple invasions 5

**Schizonts** 6
- Partially-segmented 6
- Fully-segmented 6

**Gametocytes** 7
**Healthy** 8
- Canonical healthy cells 8
    - Debris-affected cells 10
  - Non-parasitic RBC pathologies 11
    - Damaged cells due to flow shear / bad consumables 11
    - Microscopic debris on camera sensor 11
  - Abnormal RBC pathologies 11
    - Echinocytes 11
    - Edge projections / horns 11
    - Dark spots on cells (Howell-Jolly bodies(4)?) 11
    - Stomatocytes 12
    - Reticulocytes 12
    - Acanthocytes 12
    - Basophilic stippling 12
  - Ambiguous RBC appearances 12
    - “Round white dot” cells 12
    - “Two small dots” cells 12
    - Long, narrow line 12
    - Lysed Trophozoites are Misc 13
    - Ghosts 13
- Appendix A: Old UV Scope Images 14
  - Rings 14
  - Trophozoites 15
  - Schizonts 15

### Introduction, disclaimer, and general guidelines

This guide was generated based on the empirically-learned experiences of the Remoscope team at Chan Zuckerberg Biohub SF during the development of the project. The guidelines presented are the result of studying a large number of healthy and Plasmodium falciparum-infected blood samples, iteratively annotating and training models, and evaluating model performance against in vitro lab titrations and in vivo clinical data where PCR data was available.

The specific guidelines were developed based on a combination of what morphologies were seen in healthy vs. infected samples, as well as the existing literature on parasite morphologies seen in peripheral blood smears (1, 2). The appearance of parasites in Remoscope images is different from those seen in peripheral blood smears because the label-free modality exhibits a different contrast mechanism, in addition to the blood being unfixed, unstained, and subjected to flow forces. The guidelines are therefore highly specific to training models on Remoscope data.

- Use a high-contrast display such as a macbook pro or other high quality display to view the fiimages. Older desktop monitors have poor dynamic range that will inhibit proper annotation of the parasites. Turn the brightness of the display all the way up.
- Quality over quantity. Miscategorizations are a net detriment to model training, so always ensure that annotations fit the criteria defined below.
- Quantity is only important if every single annotation is self-consistent with the criteria we outline.
- Consistency is fundamentally important to training a successful model.
- Apply a general rule: if you are not >95% sure it's a parasite, label it healthy. This is because:
    - If you have no other information about the cell or the sample, odds are it's healthy
- When a RBC has more than one parasite and they are of different stages, the cell should be annotated by the latest stage present.
- Gametocytes should only be called at stages 4 or 5, once they are sufficiently distinguished from trophozoites. See below for detailed guidelines.
- Lysed RBCs lacking hemoglobin, ie. "Ghosts" should be labeled "misc" even if there is a parasite inside (the parasite will not survive)
- Regarding platelets: their size is variable and this iteration of YOGO will not directly track them. However, they should be labeled as "misc" if their diameter is greater than ¼ the size of a RBC. This is to effectively track them as debris. Small platelets not meeting this criteria should be left unlabeled.
- In reality, life stage transitions are continuous, not discrete. There are therefore many parasites that will appear to straddle the boundary between categories (ring→ trophozoite in particular). The best we can do is attempt to abide by consistent criteria, but there will be unavoidable mixing between life stage categories. Ultimately, a small amount of mixing between stages is not damaging to a clinical result because the overall parasitemia is unaffected.
- The "misc" category is a catch-all for debris, toner, ghosts, and small platelets. The purpose of labeling debris is to allow tracking of non-cellular objects that could affect parasite calls.

### Rings

Rings are visible due to their lack of Hb pigment, presenting as a white pattern against the dark background of the RBC cytoplasm. They are a challenging category because they can take on a wide variety of appearances including dendritic, amoeboid, canonical and more. A key challenge is distinguishing ring stage parasites from the widely varying uninfected RBC population. The subtypes below are only used for internal understanding, and are not sub-categorized in the annotation process at this time. Imperfect focus and motion blur can also complicate annotation of rings. In these situations, the >95% confidence guideline suggests defaulting to the "healthy" label in the event the image is not clear.

#### Canonical Rings

Canonical rings take on a characteristic annular form. Their contrast is higher than earlier forms such as amoeboid and dendritic rings. Their size and shape are relatively consistent, but their location within the RBC can vary.

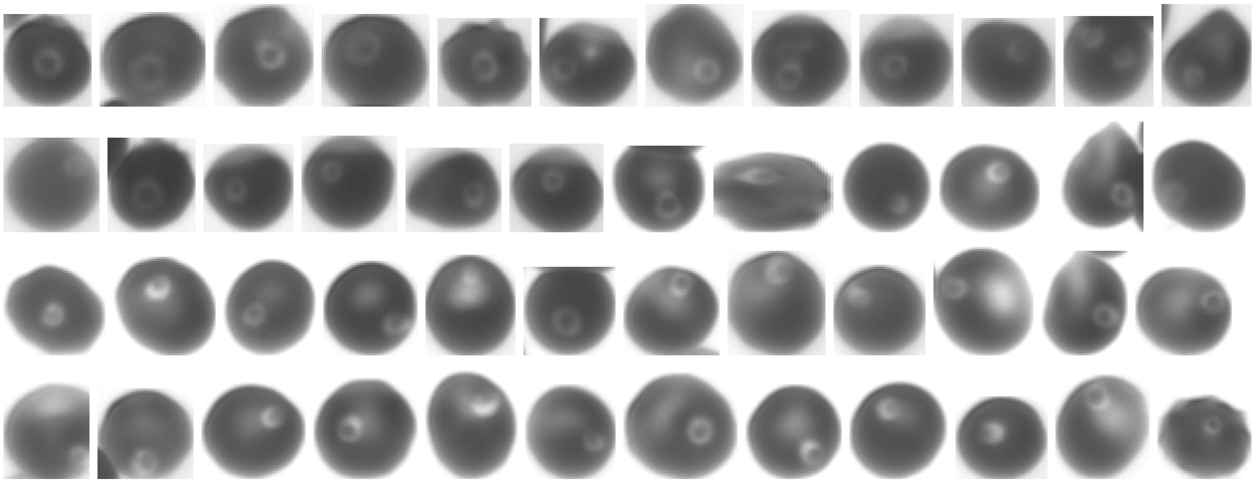

#### Early Rings

Early rings are a difficult category because they take on a very wide variety of amoeboid and dendritic shapes, sometimes making it difficult to define their appearance. They have not yet developed any hemozoin crystals and typically exhibit low imaging contrast. This may be due to a lower total cytoplasmic volume compared to mature rings, and/or a low thickness along the optical axis. Due to these challenges, annotation of early rings is generally lower in confidence compared with canonical rings or late stages. Past work has shown that parasites can even interconvert dynamically between amoeboid and dendritic forms (3, 4). We suspect that our recall is poor for this sub-category.

##### Dendritic Rings

Dendritic rings have multiple projections/lobes. These lobes can change shape over time and vary widely. Note that we score dendritic rings with three or more lobes.

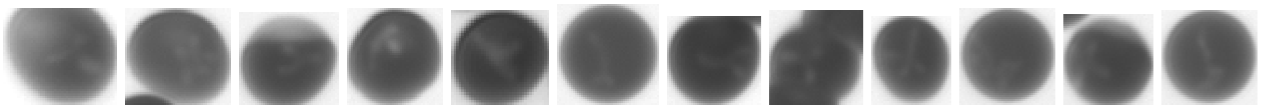

#### Amoeboid rings

Amoeboid rings have no well-defined structure, are blob-like, and range from nearly circular and pale to having mild projections. There is a continuum of morphologies between amoeboid, dendritic, and canonical rings, where the amoeboid blob-like shape variations can grow and extend outwards into dendritic form or contain a central rarefaction similar to a canonical ring. The distinction is only important insofar as recognizing geometries that we score as rings rather than healthy.

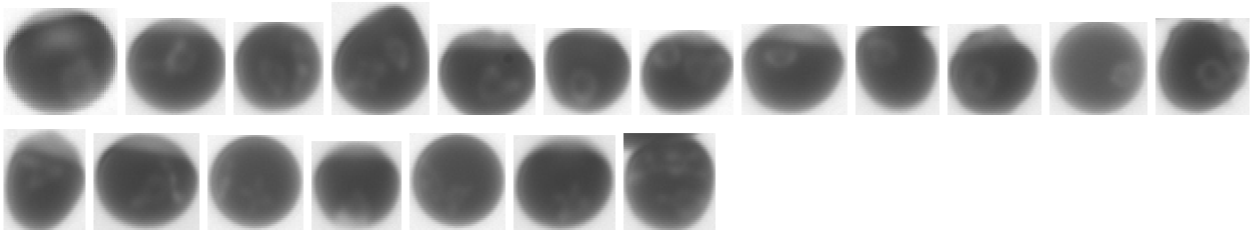

#### Narrow line rings

This is another recurring pattern not seen in uninfected populations. Note that the line does not extend across the full diameter of the RBC. A fully-spanning line is more likely to be a crease in the RBC due to flow shear. These rings are depicted in both the WHO microscopy guide (1) as well as in (2).

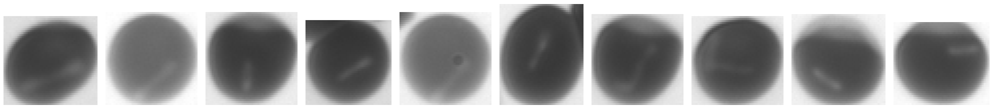

#### “Slit” rings

Rings can sometimes present off-axis and appear to have a slit down the middle:

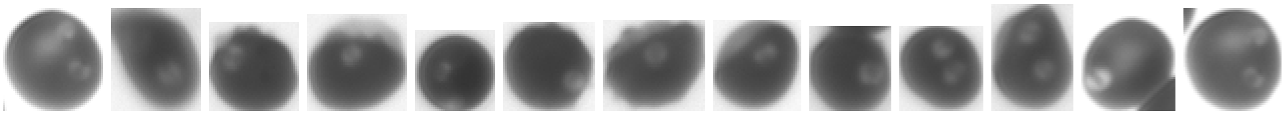

#### Multiple ring invasions

Multiple invasions are more common in cultured than in clinical samples, presumably due to the high parasitemia and lack of circulatory mixing in the stationary flasks commonly used in cultures, creating a high local concentration of merozoites after schizont ruptures.

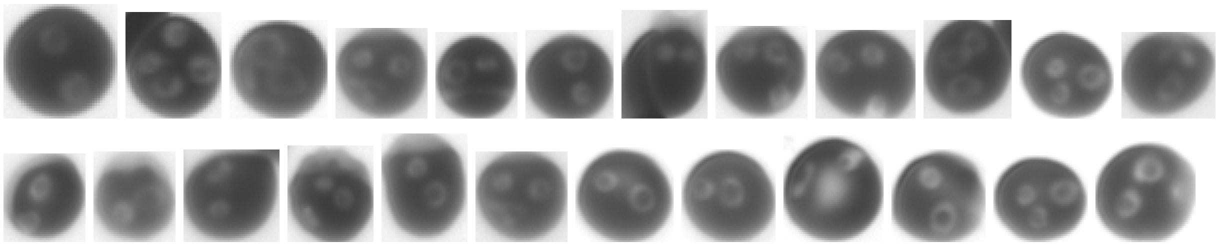

### Trophozoites

Trophozoites are more mature than rings. They are larger in size, have consumed hemoglobin from the RBC and have formed distinctive, dark hemozoin crystals. There is sometimes only one crystal or sometimes multiple distinct crystals.

**Defining characteristics:**

- Any parasite that has developed a hemozoin crystal, but has not yet segmented is a trophozoite.
- Size can vary a lot and range from the same as a ring to nearly the same as a schizont. This is the broadest category in terms of size.

#### Early trophozoites

Early trophozoites are often not distinct from rings aside from the nucleation of small hemozoin puncta. The transition to trophozoite is marked by:

- Increased solidity of shape (not dendritic - the shape is closer to circular and lacks projections)
- Small hemozoin puncta (a single dot is really hard to assess, especially if it's in the center. But two distinct, off-center dots like this example are a more sure sign)

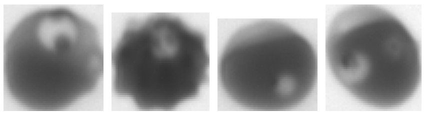

#### Mature Trophozoites

As trophozoites mature their size and contrast increases, making them more easily identifiable against the RBC cytoplasm background. Additionally the hemozoin crystals become larger and more distinct.

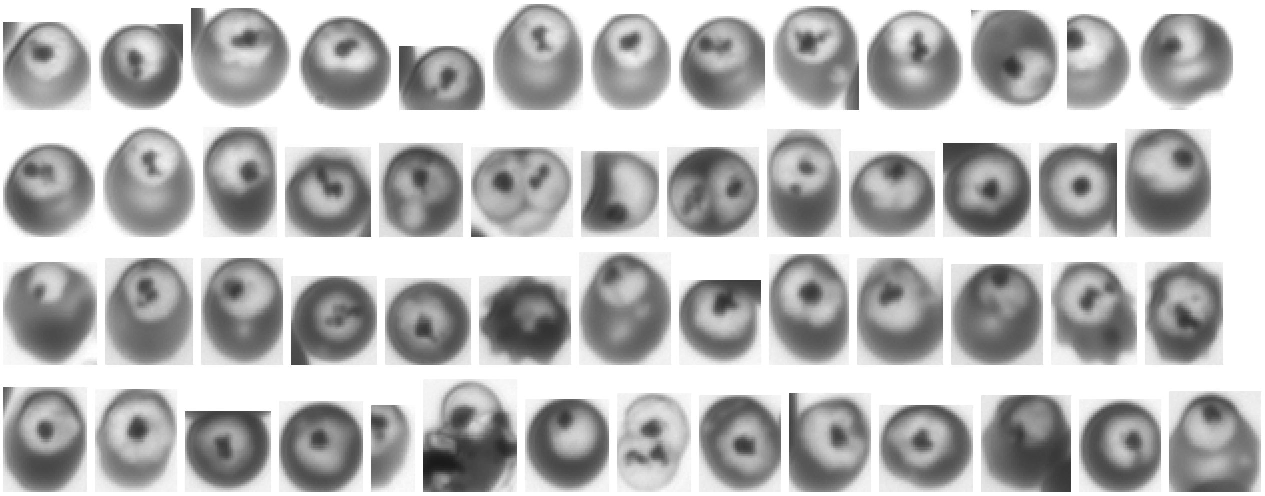

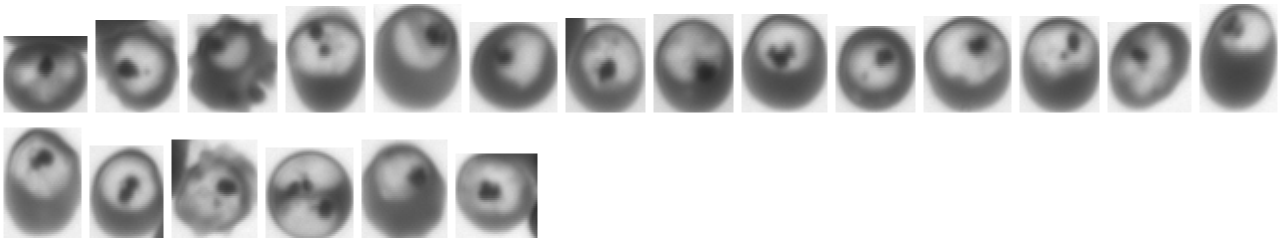

Multiple invasions

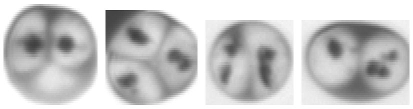

Not trophozoites:

The following

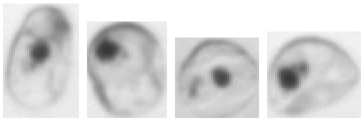

### Schizonts

Schizonts are the final stage before bursting the RBC with many merozoites, and resuming the reinfection cycle. Schizonts have begun segmenting internally into smaller ~~merozo~~ites. You can see the large heme crystal and segmented body. The RBC itself can look very pale or anemic due to its Hb being nearly gone, but not always.

**Defining characteristics:**

- Take up more than half the area of the cell
- Have a large, round, singular Hz crystal, usually centralized (but not always)
- Become segmented into merozoites: at first segmentation is subtle, but then grows into fully-separated white merozoites.
    - Segmentation can be partial (still anchored centrally), or complete (dissociated from each other)
- Hb depletion of the RBC can either be partial or complete

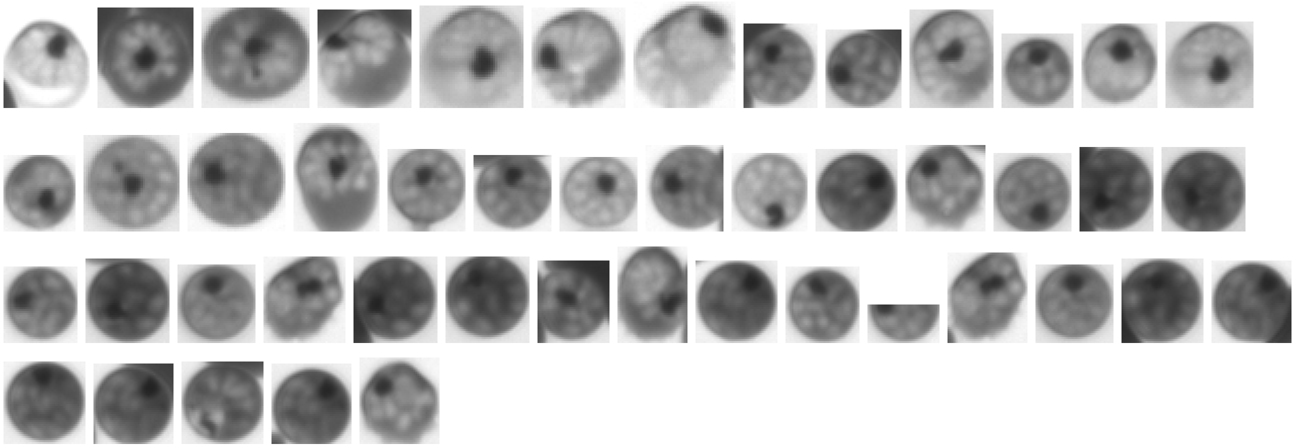

We also include cells that have near total Hb depletion and large Hz crystals, even if the segmentation is not resolved. This can be due to poor focus or other optical issues.

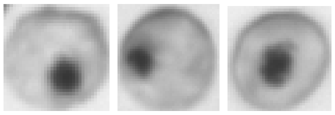

Schizont with co-inhabiting trophozoite

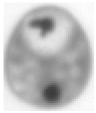

### Gametocytes

Gametocytes are the sexual form of the plasmodium parasite. During development, they begin to diverge from the appearance of the trophozoite form. Aditi Saxena maybe you can add like 1-2 sentences about this?

Although gametocytogenesis occurs over five stages (5), the earlier stages (I-III) are not reliably distinguished from trophozoites on Remoscope. We therefore annotate according to the following stage-specific criteria. We do not make the distinction between stages: all gametocytes are lumped into a single category.

**Defining characteristics:**

- Stage 1: do not call as gametocyte, call as troph instead
- Stage 2: do not call as gametocyte, call as troph instead
- Stage 3: do not call as gametocyte, call as troph instead
- Stage 4: Obvious distention of the host red blood cell, with parasite extrema visibly protruding along the long axis:

- 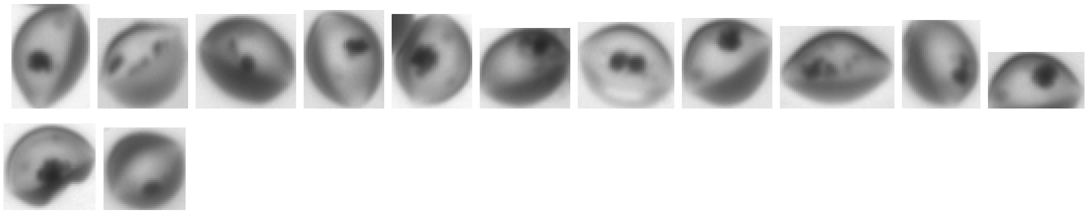
- Not included as gametocytes:
  - 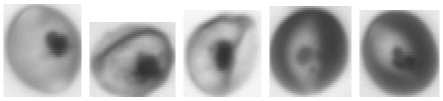
  - These would be classified as trophozoites. The parasite does not extend beyond the boundaries of the red blood cell at this point, and are not classified as gametocytes for this reason.

- Stage 5: RBC cytoplasm exists only as a thin layer on the periphery of the parasite, with an overall elongated shape either almond-like or banana-like:
  - 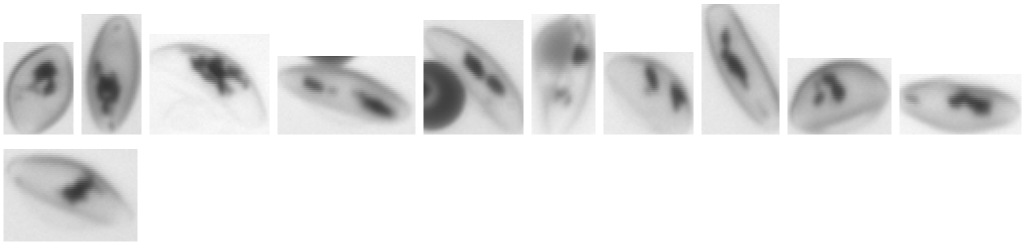

Collection of annotated gametocytes (mixed stages):

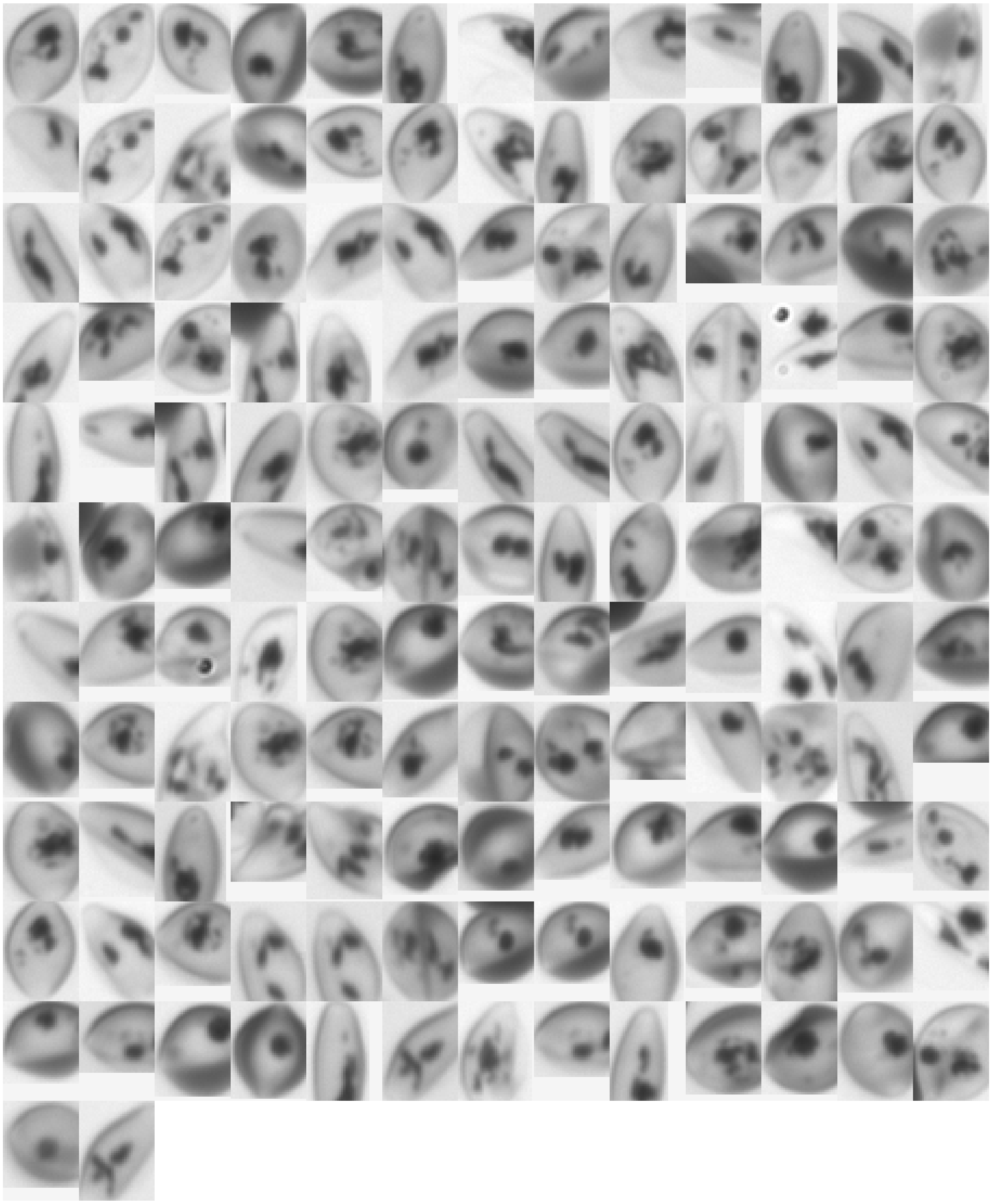

### Healthy

Healthy RBCs are nominally uniform in appearance and exhibit a canonical discocyte morphology (6). In reality, there are a large number of perturbations affecting the appearance of RBCs in Remoscope flow cells. The most dominant effects are the combination of flow shear, confinement, and collisions with other blood cells. These are physical interactions exerting mechanical stresses on the cells under flow, under which even a healthy discocyte may develop abnormal morphological features. Additionally, other RBC pathologies and/or imaging artifacts may affect their appearance, including but not limited to:

- Flow-induced shear forces
- Confinement artifacts
- Packing artifacts / collisions
- Hemoglobin abnormalities:
    - Sickle cell disease
    - Hemoglobin C disease
    - Beta-thalassemia
    - Various anemias
- Central pallor appearing as a parasite
- Debris
- Platelets overlapping with RBCs

Since the large majority of cells are healthy, it is the default category when we are not >95% sure that a cell is parasitized. Due to this class imbalance multiplier, it is generally better to lose a small amount of recall (fraction of parasites labeled as such) than to increase the false positive rate (calling healthy cells as sick).

Therefore, many abnormal-appearing RBCs get labeled as healthy – even cells that we are 80% sure are rings, for example.

#### Canonical healthy cells

Even healthy discocytes have various appearances under flow. In the best case, without other effects, the cells appear uniformly pigmented across the leading half of the cell, with a stretched, pale region at the trailing edge of the cell. The transition from symmetrical discocyte to this characteristic shape occurs as a function of flow rate.

**Healthy, high hematocrit:**

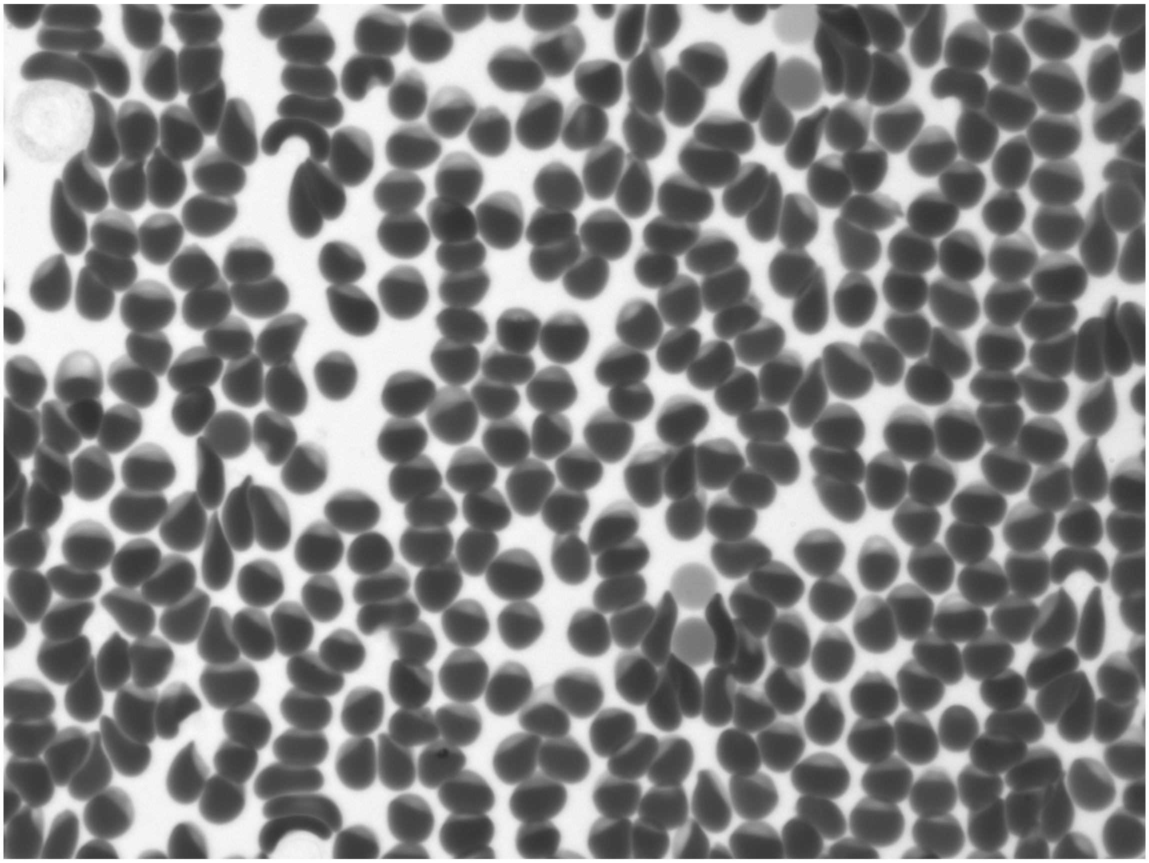

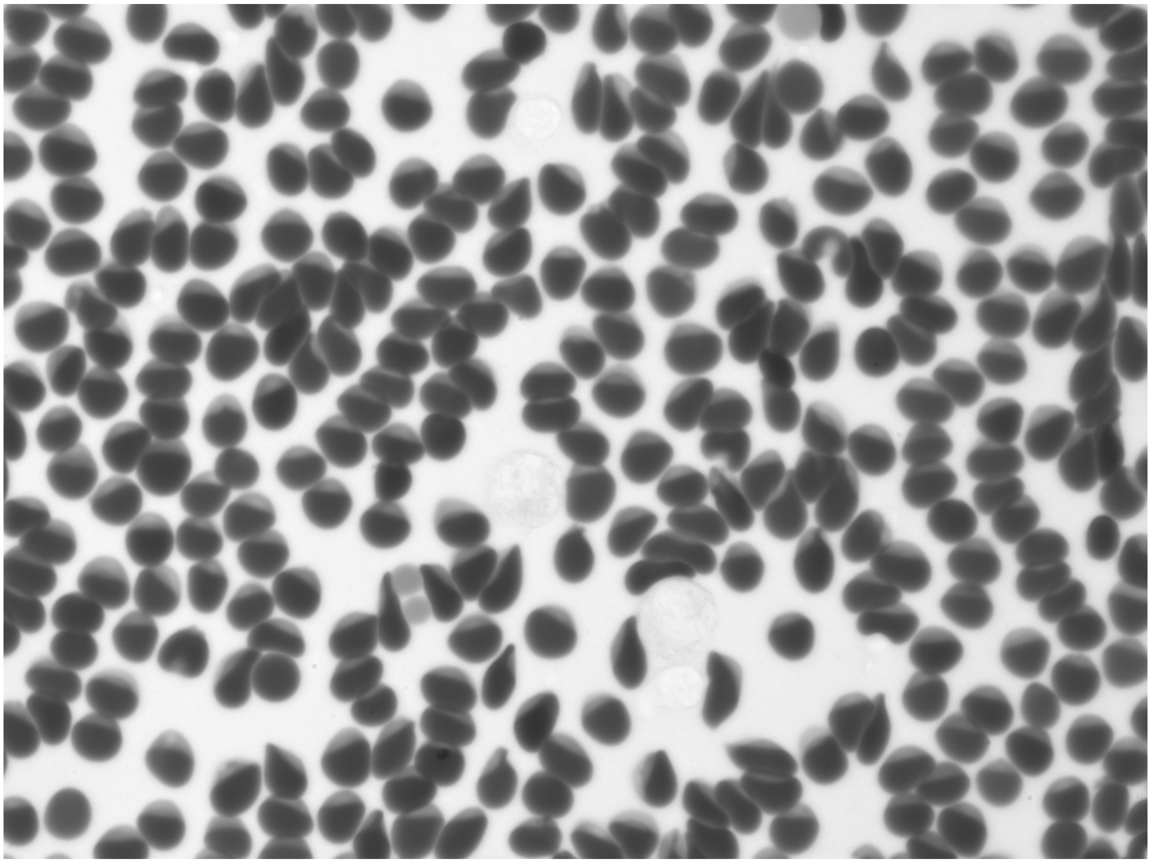

Healthy, low hematocrit, under high flow shear:

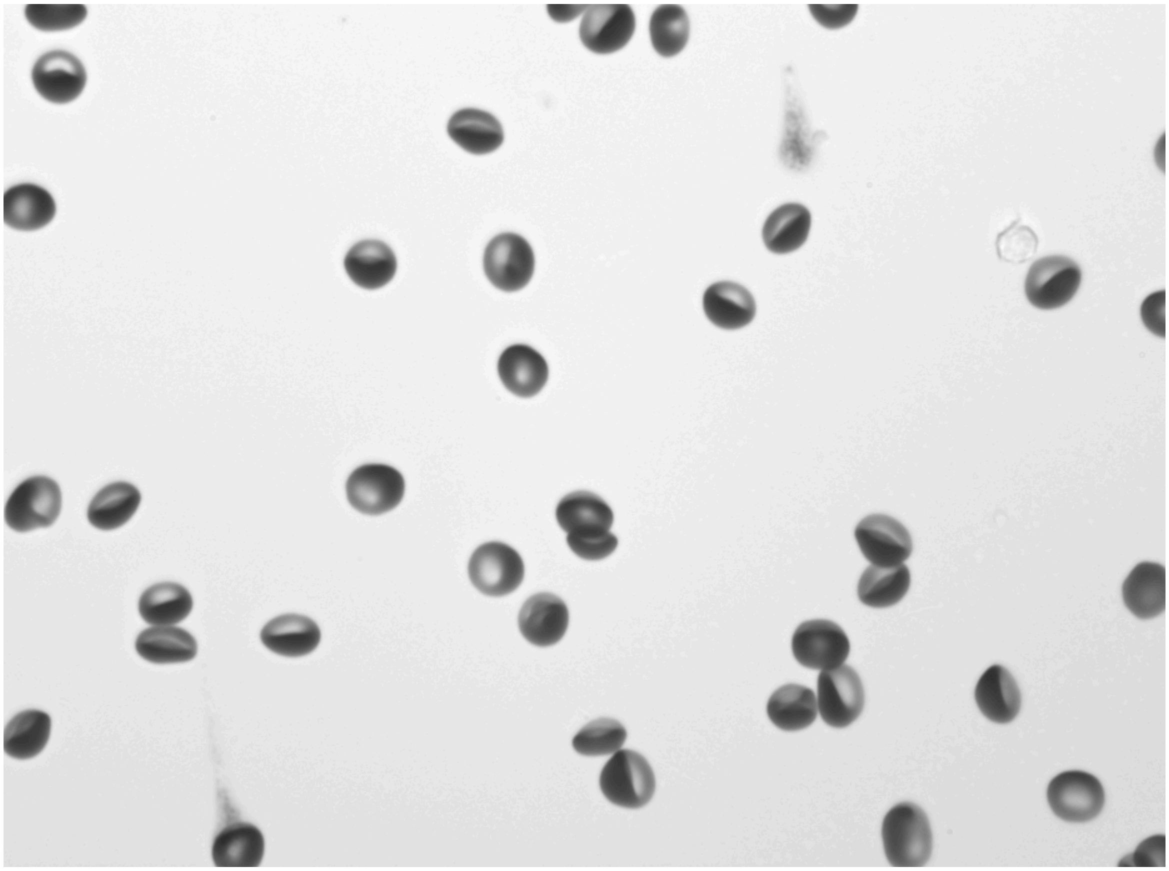

Low hematocrit, slow flow: at reduced flow rate, the RBCs recover their discocyte morphology. There is no shear distortion and the central pallor is visible.

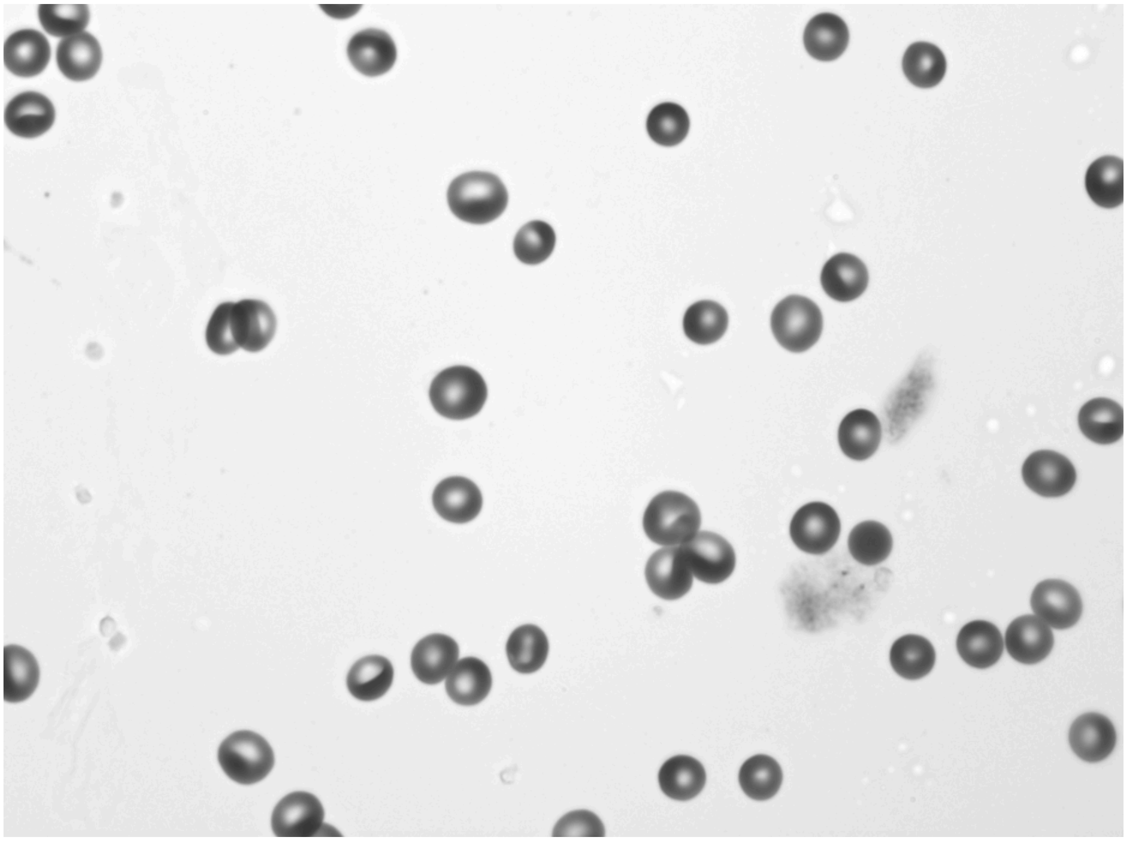

##### Debris-affected cells

The cell can pass over different types of debris, changing their appearance in ways that make them look like parasites.

- Platelets
- Stuck extracellular Hz crystals (or other dark spots)
- Toner

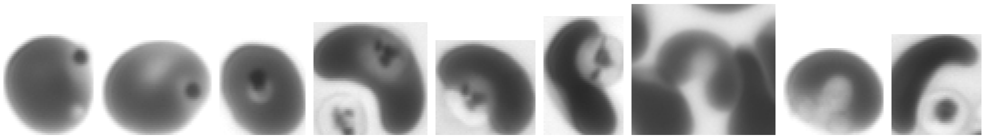

#### Non-parasitic, irregular RBC morphologies

Here is a collection of abnormal cell appearances that we will regularly encounter, but are not parasites. For more information, please see the following book chapter (6).

##### RBC lysing

Occasionally, flow chambers induce cell lysis, causing soluble hemoglobin to enter the bulk flow. RBC appearance turns to thin annuli, sometimes folding under shear. The bulk background darkens as hemoglobin absorption influences overall light transmission. This condition is rejected in the data vetting process (supplementary figure X).

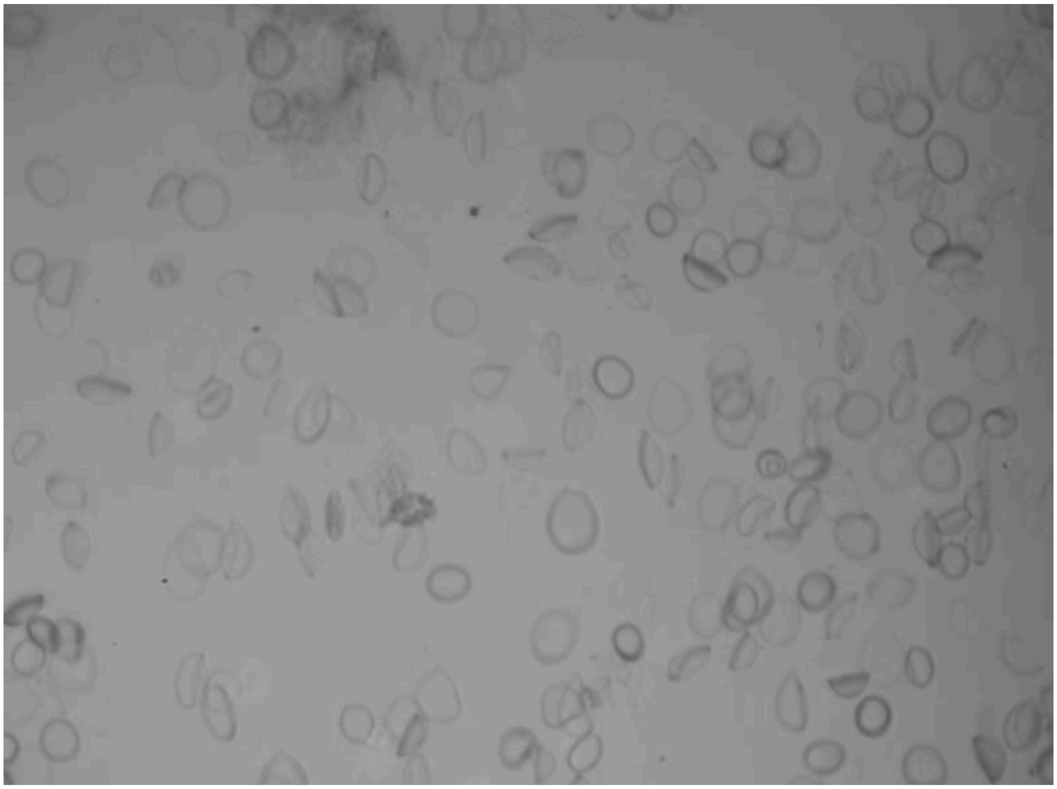

##### Microscopic debris on camera sensor

Micron-sized particles on the sensor can sometimes have tiny diffraction rings that look like miniature ring parasites. These are smaller than actual parasites and are highly symmetrical.

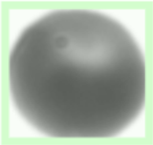

#### Abnormal RBC pathologies

##### Echinocytes

A raw image containing a majority echinocyte population, platelets, and toner debris:

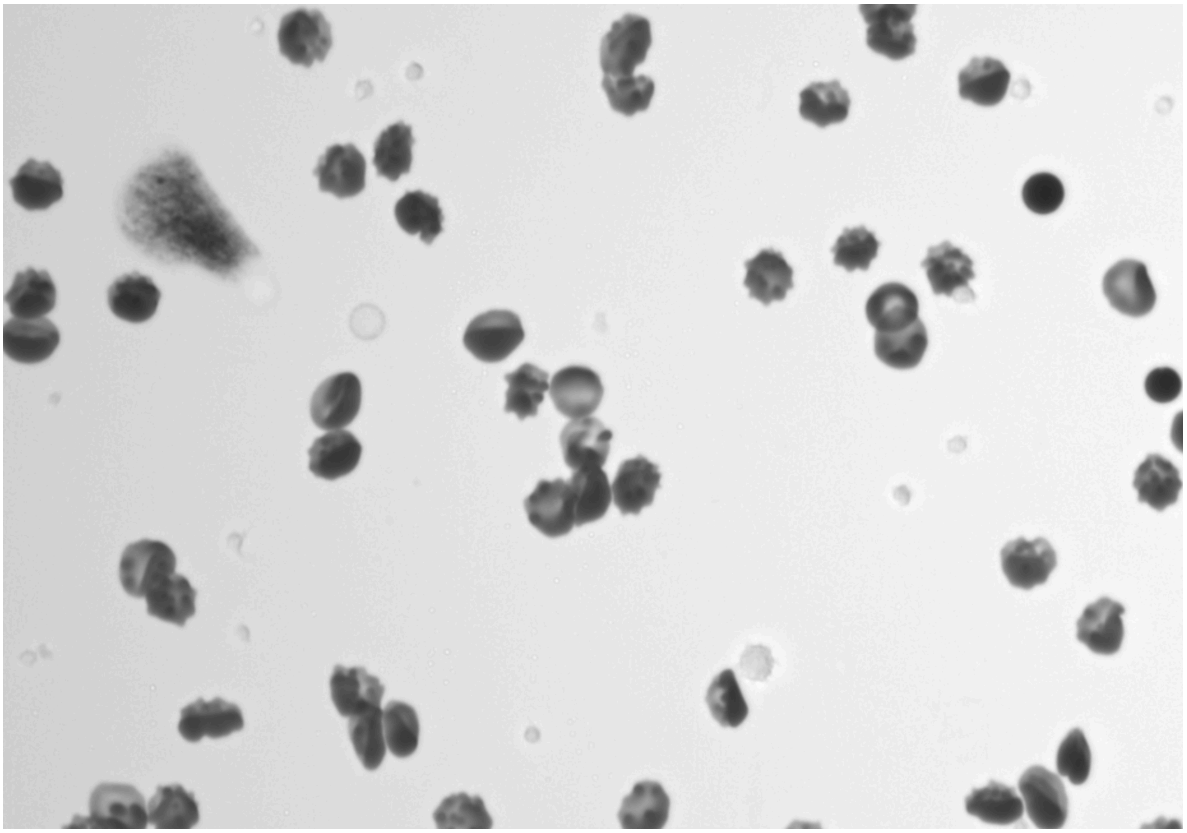

Individual echinocytes:

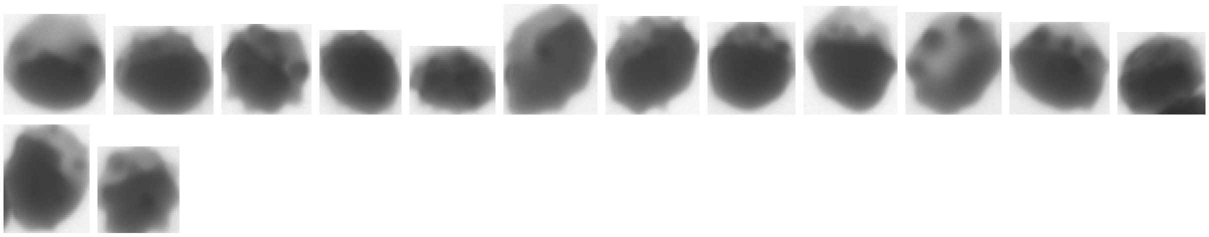

Target cells

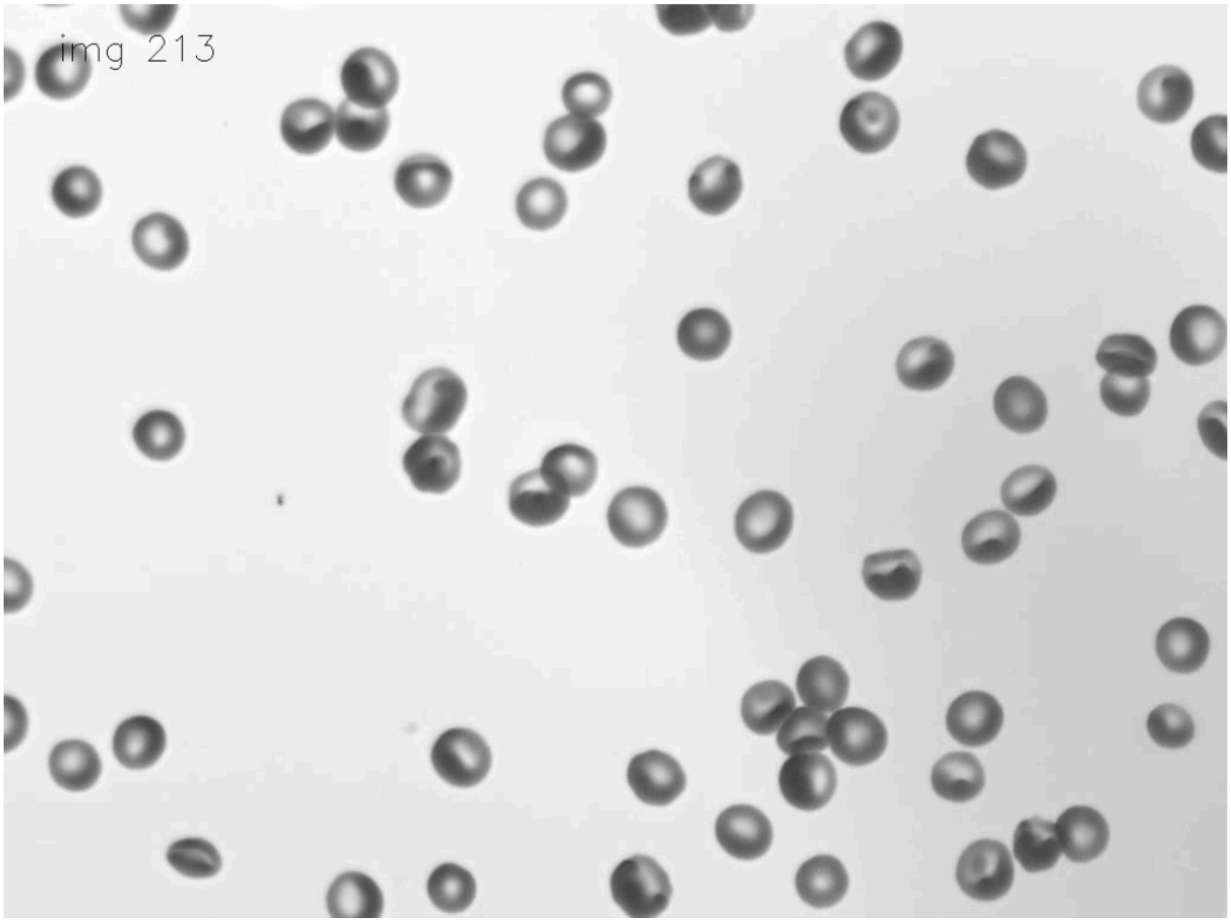

Sickle cell disease (SCD)

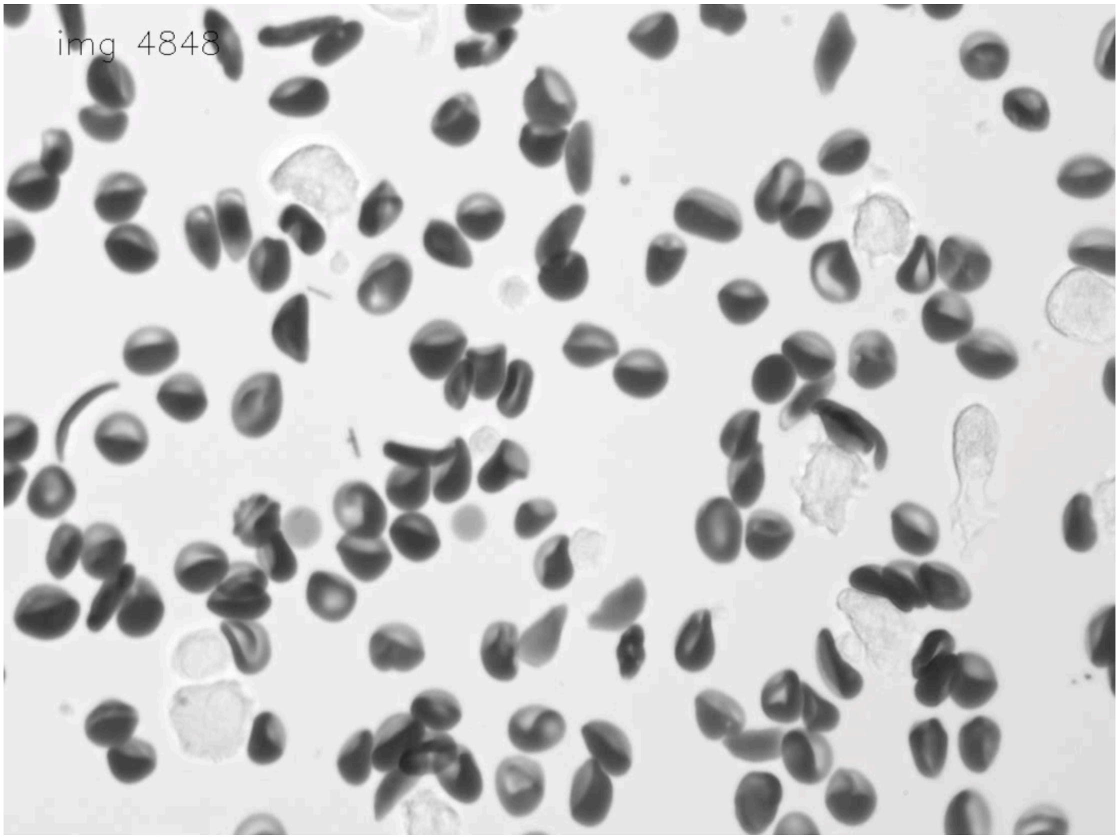


Dark spots on cells (Howell-Jolly bodies(6)?)

The presence of a dark impurity on the cell does not indicate the presence of a parasite. In this example, there is also a diffuse white region to add extra confusion. But the white region is too diffuse to be a parasite and the dark spot, possibly confused with a hemozoin crystal, is incongruous with what would otherwise be thought to be an early ring - it is too “perfect” of a circle.

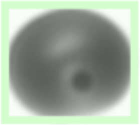

Basophilic stippling?

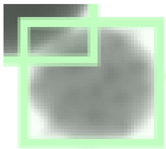

##### Stomatocytes

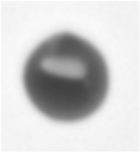

#### Parasite-like RBC appearances

Many of these closely resemble ring stage parasites, but are not distinct enough and can sometimes be observed in healthy (non-parasitic) cell populations. Therefore, we classify cells in this section as healthy despite there being some possibility we are letting a fraction of parasites go undetected.

##### “Round white dot” cells

We decided to call these healthy for now because it is a possible hemolysis pattern, or they might be dead parasites. We are not sure yet, so we have to call them healthy. These don’t often appear in healthy cultures, and other publications detailing the parasite life cycle do not show anything like this. They are characterized by a high level of symmetry and high contrast. Note: drug-inhibited trophozoites exhibit a similar appearance due to hemozoin polymerase inhibition.

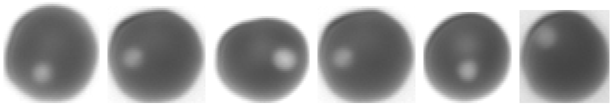

##### “Two or more small dots” cells

This is a recurring pattern that may be early rings but are not common or distinct enough to be called rings.

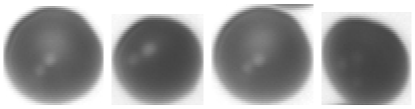

### Miscellaneous

YOGO has a limited number of classes it can  since it runs in real-time on embedded hardware. Therefore, we need to lump any remaining objects in the field of view into a ‘misc’ category. Despite not explicitly needing to count them, we need to detect them in order to understand if and when other objects could be occluding or otherwise interfering with actual RBCs. Here is an incomplete list of objects we want to include in this category:

- Toner blobs

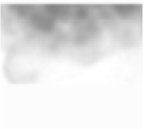

- Other physical debris

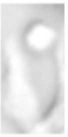

  - Sometimes cells overlap with debris. Only label as ‘misc’ if there’s enough debris to obscure the cell:

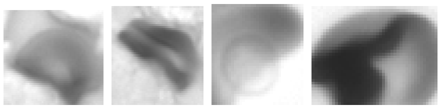

  - Example of ‘healthy’, NOT ‘misc’:

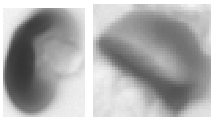

- Slivers of cells (optional)
  - Most of these will be rejected by area filtering anyways. Only classify as ‘misc’ if it’s unclear what is in the thumbnail

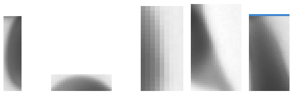

  - Example of ‘healthy’, NOT ‘misc’ (deformed cells or partially cut off thumbnails):

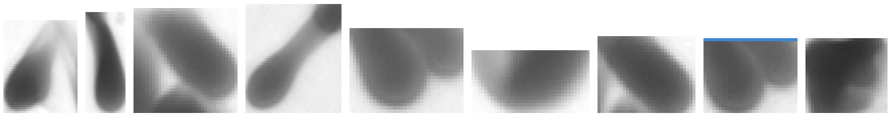

  - RBC fragments look like RBCs but are much smaller than typical RBCs do not need to be moved during thumbnail sorting, as they will also be rejected by area filtering

- Thumbnails that are centered between two cells

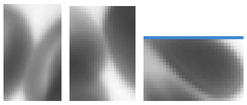

- Ghost RBCS (RBCs without hemoglobin)
- Extracellular parasites and parasite debris

##### Lysed Trophozoites are labeled as Misc

If the RBC does not have any hemoglobin remaining, but the parasite has not matured to a schizont, then the parasite is likely dead and should not be counted.

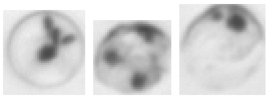

##### Ghosts

These are RBCs that have been ruptured and lost their Hb. We are not counting these as cells. Since they are so pale I'm not sure it really matters, but we can call them 'misc' if they are present.

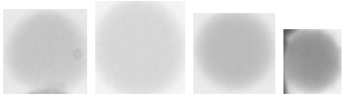

The example below would be classified as 'healthy' as it has enough hemoglobin.

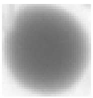
