## Supplementary Note 2 for "Remoscope: a label-free imaging cytometer for malaria diagnostics"

### Supplementary note 2: Statistical analysis and error estimation

Our statistical methods for quantifying estimation uncertainty capture both Poisson sampling error as well as uncertainty propagated from imperfect knowledge of average classification error rates, as expressed in the confusion matrix. Final uncertainty estimates are expressed in units of parasitemia – the relative fraction of parasitized red blood cells in the sample.

**Glossary of vector symbols.** The following symbols refer to vectors used throughout this supplementary note.

The $i$-th element of a vector $x$ is referenced with the notation $[x]_i$. Note that each value in the vector corresponds to a prediction class.

The following are all row vectors, in units of absolute class counts:

| | |
|---|---|
| $n_{actual}$ | True class counts |
| $n$ | Raw YOGO class count |
| $\hat{n}$ | Unbiased (compensated) YOGO class count |
| $\sigma_{\hat{n}}$ | Total class count uncertainty |

**Glossary of matrix symbols.** The following symbols refer to unitless matrices used throughout this supplementary note, where the element in row i and column j of matrix $X$ is referenced with the notation $[X]_{ij}$:

| | |
|---|---|
| $M$ | Confusion matrix |
| $M^{-1}$ | Inverse confusion matrix |
| $\sigma_M$ | Uncertainty of each confusion matrix value |
| $\sigma_{M^{-1}}$ | Uncertainty of each inverse confusion matrix value |

**Data compensation.** The neural network has average rates of misclassification with respect to human annotation, defined by the confusion matrix $M$. Here we define the rows of the confusion matrix to represent ground truth and the columns to represent model-predicted classes.

When evaluating the predicted counts for each class, the raw count $n$ is biased by the average rates defined in $M$:

(CMC-1) $$n_{actual} M = n$$

We can generate unbiased estimates of the true class counts by solving for $n_{actual}$. Given a row vector of predicted class counts $n$, the unbiased estimates $\hat{n}$ are given by

(CMC-2) $$\hat{n} = nM^{-1}$$

**YOGO confusion matrix.** We have two modalities for data compensation. The first directly uses the 7 x 7 confusion matrix $M$ that includes all YOGO prediction classes, such that the *i*-th element of a vector or matrix corresponds with

(YC-1) $i \in \{healthy,\ ring,\ trophozoite,\ schizont, gametocyte,\ WBC,\ miscellaneous\}$

To ensure $M$ is representative of the average misclassification rate, we used k-fold validation such that $M$ is the mean of every confusion matrix generated by $k = 5$ partitions of YOGO's test dataset.

Since $M$ is generated by averaging $k = 5$ confusion matrices element-wise, we generate each element's standard deviation $[\sigma_{M^{-1}}]_{i,j}$ by individually inverting each $k = 5$ confusion matrices and taking the standard deviation across all inverses.

**Fitted aggregate confusion matrix.** We can also use an aggregated confusion matrix that is based on fitting our clinical Uganda data to corresponding PCR results. We only consider RBCs in this case, such that

(FAC-1) $i \in \{\ healthy\ (h),\ parasites\ (p)\ \}$

where parasites include all asexual parasite stages (ring, trophozoite, and schizont). The resulting confusion matrix is 2 x 2 and can be understood as

(FAC-2) $M = [\ [TN,\ FP]\ ,\ [FN,\ TP]\ ]$

where $TN$, $FP$, $FN$, and $TP$ are *true negative*, *false positive*, *false negative*, and *true positive* respectively.

We compare Remoscope's parasitemia output $P_{Remoscope}$ to the PCR value $P_{PCR}$ to solve for $m$ and $b$ using a weighted least squares regression:

(FAC-3) $P_{Remoscope} = m * P_{PCR} + b$

Applying the inverse of this fit to estimate compensated parasitemia $\widehat{P}$ from Remoscope's output yields

(FAC-4) $\widehat{P} = (P_{Remoscope} - b)\ /\ m$

We express this linear operation as a matrix multiplication by conserving the total RBC count $N$ according to

(FAC-5) $N = \widehat{[n]}_h + \widehat{[n]}_p = [n]_h + [n]_p$

and linearly expanding equation (CMC-2) as

(FAC-6) $\widehat{[n]}_h = [n]_h\,[M^{-1}]_{11} + [n]_p\,[M^{-1}]_{21}$

(FAC-7) $\widehat{[n]}_p = [n]_h\,[M^{-1}]_{12} + [n]_p\,[M^{-1}]_{22}$

Substituting the definition of parasitemia

(FAC-8) $P = [n]_p\ /\ N$

into equation (FAC-4), results in the equality

(FAC-9) $\widehat{[n]}_p\ /\ N = [n]_p\ /\ (N * m) - b$

Multiplying equation (FAC-9) by $N$ and substituting in equations (FAC-7) and (FAC-5) for $\widehat{[n]}_p$ and $N$ yields

(FAC-10) $[n]_h\,[M^{-1}]_{12} + [n]_p\,[M^{-1}]_{22} = [n]_p\ /\ m - \left([n]_h + [n]_p\right) * b$

We match terms to determine two matrix elements

(FAC-11) $[M^{-1}]_{12} = \ - b$

(FAC-12) $[M^{-1}]_{22} = 1/m - b$

and the other matrix elements are constrained by normalization to satisfy equation (FAC-5):

(FAC-13) $[M^{-1}]_{11} = 1 - [M^{-1}]_{12} = 1 + b$

(FAC-14) $[M^{-1}]_{21} = 1 - [M^{-1}]_{22} = 1 - 1/m + b$

The standard deviation of each matrix element can be computed by error propagation

(FAC-15) $[\sigma_{M^{-1}}] = \left[ \left[ \sigma_b \, , \, \sigma_b \right], \left[ \sqrt{\sigma_b^{\ 2} + \sigma_m^{\ 2}} \, , \sqrt{\sigma_b^{\ 2} + \sigma_m^{\ 2}} \right] \right]$

where $\sigma_b$ and $\sigma_m$ can be derived from the covariance matrix output by the linear fitting algorithm.

**Class count error estimation methods.**

We estimate the error $\sigma_{\hat{n}}$ in the unbiased estimates $\hat{n}$ by linearly expanding equation (CMC-2) as

(CCU-1) $[\hat{n}]_j = \sum_i [n]_i \, [M^{-1}]_{ij}$

and applying general error propagation:

(CCU-2) $[\sigma_{\hat{n}}]_j^{\ 2} = \sum_i \left( \frac{\partial [\hat{n}]_j}{\partial [M^{-1}]_{ij}} \right)^2 [\sigma_{M^{-1}}]_{ij}^{\ 2} + \sum_i [\sigma_n]_i^{\ 2} \left( \frac{\partial [\hat{n}]_j}{\partial [n]_i} \right)^2$

The first error term $[\sigma_{M^{-1}}]_{ij}$ is the uncertainty of each element in the inverse confusion matrix. Since $M$ is generated by averaging $k = 5$ confusion matrices element-wise, we generate $\sigma_{M^{-1}}$ by inverting each individual confusion matrix and taking the standard deviation across all $k = 5$ inverses.

The second error term $[\sigma_n]_j$ comes from the fundamental random sampling uncertainty of the raw class counts, which can be computed using the equality of mean and variance in the Poisson distribution:

(CCU-3) $[\sigma_n]_j = \sqrt{[n]_j}$

Substituting equations (CCU-1) and (CCU-3) into equation (CCU-2) yields

(CCU-4) $[\sigma_{\hat{n}}]_j^{\ 2} = \sum_i \left( [n]_i^{\ 2} \, [\sigma_{M^{-1}}]_{ij}^{\ 2} + [n]_i \, [M^{-1}]_{ij}^{\ 2} \right)$

**Parasitemia error estimation methods.**

We provide a clinically relevant output by computing the parasitemia from the class counts $\hat{n}$. Parasitemia is computed from the ratio of asexual parasitized red blood cells to all red blood cells. We include the following classes in the parasite and red blood cell counts, respectively:

(PEM-1) $\mathbb{P} = \{ring, \ trophozoite, \ schizont\}$

(PEM-2) $\mathbb{R} = \{healthy, \ ring, \ trophozoite, \ schizont\}$

The total parasite count $\hat{n}_{\mathbb{P}}$ and red blood cell count $\hat{n}_{\mathbb{R}}$ can thus be defined as

(PEM-3) $\hat{n}_{\mathbb{P}} = \sum_{j \in \mathbb{P}} [\hat{n}]_j$

(PEM-4) $\hat{n}_{\mathbb{R}} = \sum_{j \in \mathbb{R}} [\hat{n}]_j$

and parasitemia $P$ can be computed as

(PEM-5) $P = \hat{n}_{\mathbb{P}} / \hat{n}_{\mathbb{R}}$

The relative parasitemia uncertainty $\delta_{\mathbb{P}}$ can be derived from the absolute parasite count uncertainty $\sigma_{\hat{n}_{\mathbb{P}}}$ by dividing by the total RBC count and total parasitemia $P$

(PEM-6) $\delta_{\mathbb{P}} = \sigma_{\hat{n}_{\mathbb{P}}} / (\hat{n}_{\mathbb{R}} \times P)$

Substituting equation (PEM-5) into equation (PEM-6) yields the simplified form

(PEM-7) $\delta_{\mathbb{P}} = \sigma_{\hat{n}_{\mathbb{P}}} / \hat{n}_{\mathbb{P}}$

where $\sigma_{\hat{n}_{\mathbb{P}}}$ can be computed from the individual class uncertainties $[\sigma_{\hat{n}}]_j$ in equation (CCU-4) as

(PEM-8) $\sigma_{\hat{n}_{\mathbb{P}}} = \sqrt{\sum_{j \in \mathbb{P}} [\sigma_{\hat{n}}]_j^2}$

Note that for negative samples, $\hat{n}_{\mathbb{P}} = 0$ so equation (PEM-7) is undefined. In this case, we use the rule of three to compute $\delta_{\mathbb{P}}$:

(PEM-9) $\delta_{\mathbb{P}} = \hat{n}_{healthy} / 3$

Given $\delta_{\mathbb{P}}$, we use the known area of a normal distribution, to compute the 95% confidence bound as

(PEM-10) $bounds = [\, max(0,\ P - 1.96 * \delta_{\mathbb{P}}),\ min(P + 1.96 * \delta_{\mathbb{P}},\ 1)\,]$

**Early termination condition.** We optionally terminate the experiment once the parasite count uncertainty falls below 5%. Since the parasitemia uncertainty $\delta_{\mathbb{P}}$ is computed according to equation (PEM-8), we terminate the experiment once the following condition is met:

(ETC-1) $\delta_{\mathbb{P}} < 5\%$

**Computing compensation parameters.** Parasitemia can be compensated for the known recall and false positive rate by $Y_c = (Y_{raw} - FPR) / recall$. Compensation factors were computed by weighted linear regression using the inverse of the PCR value as weights. PCR-0 values were clipped to 1 to prevent exploding weights. We find that fit values are assay-dependent: recall was higher and FPR was lower for undiluted blood than for diluted blood. This is hypothesized to be due to an increase in RBC morphological defects, caused by diluent reconstitution issues.
